# Long-Run Public Health Impact of Doxycycline Post-Exposure Prophylaxis and Behavioural Factors on Syphilis Transmission: A Modelling Study in Singapore and England

**DOI:** 10.1101/2025.07.28.25332286

**Authors:** Zihao Wang, Dariya Nikitin, Borame L. Dickens, Liang En Wee, Martin T.W. Chio, Rayner Kay Jin Tan, Keisuke Ejima, Yi Wang, David N. Fisman, Lilith K. Whittles, Jue Tao Lim

## Abstract

Syphilis remains a significant global public health challenge, particularly among men who have sex with men (MSM). Although penicillin is highly effective for treatment, primary prevention strategies are limited. Recent trials indicate that doxycycline post-exposure prophylaxis (doxy-PEP) has high efficacy in reducing syphilis incidence among MSM; however, its long-run population-level impact, effects on transmission dynamics, and optimal prescribing strategies remain unclear, especially when accounting for real-world behavioural factors such as screening frequency, uptake, adherence, and discontinuation. To address this gap, we developed a behavioural transmission-dynamic model calibrated with Bayesian methods using epidemiological and sexual behavioural data from Singapore and England to characterize transmission dynamics in MSM, quantify the potential long-run public health impact, efficiency, and robustness of alternative doxy-PEP prescribing strategies across different settings (e.g., schools, clinics, age, and risk groups) under varying behavioural patterns and epidemiological settings. Over a 15-year horizon, targeting high-risk MSM at diagnosis emerged as the most efficient approach, averting an estimated 2.50 (0.68 - 5.94) cases per prescription (10000 [95% Credible Interval 1100 - 53200] total cases averted) in Singapore and 4.60 (2.12 - 7.79) cases per prescription (165000 [49300 - 503700]) in England. In contrast, broader strategies such as offering doxy-PEP to all MSM attending sexual health clinics could achieve greater overall reductions (up to 24700 [6800 - 93100] cases in Singapore and 279800 [109200 - 724000] in England), but with substantially lower efficiency, averting as few as 0.02 (0.00 - 0.23) and 0.04 (0.01 - 0.46) cases per prescription, respectively. These findings suggest that untargeted strategies could substantially reduce syphilis incidence but would do so at the cost of over-prescription, increased resource burden, and unnecessary antibiotic exposure. More importantly, our findings remain robust despite variations in behavioural factors and future scenarios. In summary, our results underscore that doxy-PEP prescribing approaches aligned with behavioural risk factors can maximise population-level impact and implementation efficiency, supporting more sustainable syphilis prevention among MSM.

## 1. Introduction

Sexually transmitted infections (STIs) are among the most prevalent communicable diseases worldwide, posing a significant burden on global health and well-being. Among these, syphilis (*Treponema pallidum*) remains a major public health concern. and is a chronic, systemic STI. Globally, around 8 million individuals acquire disease yearly, with an increasing incidence in the past 2 decades [1]. In England, male syphilis incidence has risen sharply, increasing from 10.1 to 29.0 cases per 100,000 population between 2011 and 2024 [2], [3]. Whereas in Singapore, the incidence of syphilis among men has remained persistently high, fluctuating between 32.5 and 38.1 cases per 100,000 population from 2004 to 2018 [4], [5]. Even though syphilis diagnoses showed a decreasing trend during the COVID-19 pandemic, likely due to reduced sexual activity during lockdowns and decreased testing availability [6], evidence suggests that transmission rates may now be exceeding pre-pandemic levels as non-pharmaceutical interventions have been lifted [7].

Syphilis transmission is influenced by a complex interplay of behavioural, biological, and social factors. High partner turnover rate, inconsistent condom use, anonymous sexual encounters, and the use of geospatial dating applications facilitate frequent partner change within dense sexual networks, increasing opportunities for exposure and reinfection [8]. Anal intercourse, which has a higher transmission efficiency, further contributes to elevated risk [9]. Social and structural barriers - including stigma, discrimination, and limited access to culturally competent healthcare - can delay diagnosis and treatment, especially in key populations [10]. Gay, bisexual, and other men who have sex with men (MSM) as well as transgender women (TGW) are disproportionately affected by syphilis and other STIs [11]. MSM are estimated to comprise between 0.03% and 6.5% of the male population across various populations [12], yet account for a large proportion of syphilis cases in many high-income countries (in the European Union and European Economic Area, MSM accounted for 74% of syphilis cases with known transmission category in 2019) [13].

Despite the widespread incidence of syphilis, there are currently no vaccines and limited chemoprophylaxis options available for preventing infections [11]. To address these growing concerns, doxycycline post-exposure prophylaxis (doxy-PEP) has recently gained attention as a promising pharmaceutical intervention for STI prevention [14], [15], with the British Association for Sexual Health and HIV (BASHH) [16] and the US Centers for Disease Control and Prevention (CDC) [11], each providing their own recommendations and guidance. Doxy- PEP involves providing a 200 mg dose of immediate-release doxycycline in advance, which an individual can then take within 72 hours after condomless sex to reduce the risk of bacterial STIs, including syphilis. While still in the early stages of clinical adoption globally, this approach is being integrated into healthcare practices in countries such as the US [17].

Doxy-PEP efficacy and effectiveness have been demonstrated across multiple clinical trials and implementation settings, with ongoing studies in Canada and Australia aiming to further evaluate its effectiveness and broader implications [18], [19]. Clinical evidence from five open-label randomized trials in France [14], [15], the US [20], [21], and Kenya [22] has demonstrated high efficacy of doxy-PEP in lowering the incidence of syphilis among MSM and cisgender women. Additionally, the implementation of doxy-PEP guidelines in San Francisco, California, was linked to a decline in reported cases of early syphilis among MSM and transgender women [23]. A retrospective cohort study at Kaiser Permanente Northern California further found that doxy-PEP use was linked to significant reductions in syphilis incidence among HIV pre-exposure prophylaxis (PrEP) users [24]. Another study demonstrated that doxy-PEP was particularly effective in reducing syphilis incidence among HIV PrEP users [25], while further evidence found that prescribing strategies based on individuals’ STI history were more effective than those based solely on HIV status or PrEP use [26], highlighting the importance of risk-based targeting in implementation strategies. Furthermore, an analysis of electronic health records in Boston, Massachusetts, estimated that implementing doxy-PEP for individuals with bacterial STIs, especially those with concurrent or recurrent infections, could substantially lower overall STI rates among gay, bisexual, and other MSM, transgender women, and nonbinary individuals assigned male at birth who accessed STI testing at a community health centre [26].

Given that the acceptability of antibiotic chemoprophylaxis such as doxy-PEP is high (e.g., about 75.8% among Australian MSM) [27], [28], it remains crucial to understand how its use might influence long-run, population-level syphilis trends - especially when accounting for real-world behavioural dynamics such as uptake, adherence, and discontinuation. Emphasizing behavioural factors is essential because these directly impact intervention effectiveness; variations in how individuals initiate, continue, or cease prophylaxis can significantly alter transmission dynamics and the overall success of the programme. To date, no study has quantified the potential long-run public health impact and efficiency of doxy-PEP under different prescribing strategies, nor examined how targeting high-risk groups may influence overall transmission dynamics, reduce the total number of infections, or optimise the number of prescriptions needed. It is also uncertain which implementation strategies - such as prescribing doxy-PEP broadly versus focusing on subgroups with the highest behavioural risk - can achieve the greatest reduction in syphilis incidence while minimising unnecessary antibiotic exposure.

Thus, the goal of this study is to evaluate how doxy-PEP can be optimally deployed to maximize public health benefits. Specifically, we aim to ensure that individuals who are most likely to benefit from doxy-PEP have access to it, while reducing the total number of prescriptions to limit potential adverse effects. More importantly, to date, no comprehensive population-level modelling study has explored the long-run transmission dynamics of syphilis in the context of doxy-PEP as a preventive intervention.

This study addresses this gap by developing a behavioural transmission-dynamic model of doxy-PEP for syphilis, calibrated to epidemiological and sexual behavioural data streams from both Singapore and England. These two countries with stabilising and increasing incidence of syphilis were used to compare the performance of different population-health strategies to prescribe doxy-PEP in changing epidemiological contexts. To examine long-run impacts, the model projects syphilis trends over 2026 to 2040 and assess the comparative public health impact, efficiency, and robustness of various doxy-PEP prescribing strategies under different behavioural scenarios. Prescribing strategies were modelled such that they could be practically implemented in healthcare settings, and we would examine the individual and combined public health impacts of these strategies when prescription is delivered across different settings, such as schools, clinics, by age and risk groups. Importantly, our framework also integrates individual-level behavioural heterogeneities - such as STI screening frequency, uptake of doxy- PEP, adherence rates, discontinuation patterns, partner change rates, and the degree of assortative sexual mixing - into a population-level framework to comprehensively evaluate the implementation of doxy-PEP, and determine the potential impact and efficiency of each strategy. In addition, we explore how key behavioural factors - such as asymptomatic screening rates (in this work, we define the asymptomatic screening rate as the rate at which both susceptible individuals and asymptomatic infections are detected through routine diagnostic testing), uptake, adherence, and discontinuation - influence the relative impact and efficiency of each strategy. This analysis provides a robust framework for optimizing doxy-PEP deployment in diverse epidemic settings, balancing population health impact and efficiency.

## 2. Methodology

### a. Data

We used syphilis incidence data from Singapore and England to calibrate the model. We consider the sexually-active population aged as 15-65. For Singapore, annual male syphilis incidence rates from 2004 to 2018 were obtained from the Communicable Diseases Surveillance annual reports published by the Ministry of Health [6]. Age- and gender-specific population data were sourced from the Singapore Department of Statistics. As the total number of MSM is unknown in Singapore, we estimated the annual number of MSM syphilis diagnoses using three scenarios, adapted from our previous work [29]: (A) main scenario: male syphilis incidence minus female incidence, assuming all excess male syphilis diagnoses represent infections in MSM; (B) upper bound scenario: total male syphilis incidence, assuming all reported male syphilis cases are among MSM; and (C) lower bound scenario: male syphilis incidence scaled by the proportion of MSM in the male population (about 6.09%) [30], assuming equal infection risk across all men. For England, annual numbers of MSM syphilis diagnoses from 2011 to 2024 were retrieved from the Genitourinary Medicine Clinic Activity Dataset [3]. Due to data quality concerns related to COVID-19 disruptions, data from 2021 and 2022 were excluded from our analysis. The detailed data sources and estimates are provided in Supplementary Information Appendix B.

### b. Doxy-PEP model for syphilis

To evaluate and compare the impact and efficiency of various doxy-PEP strategies for syphilis, we developed a novel behavioural transmission-dynamic model (see Fig. 1 for a schematic overview and parameter descriptions). The model stratifies the MSM population into two risk groups based on sexual behaviour: high-risk individuals with five or more sexual partners per year, and low-risk individuals with fewer than five. This stratification captures behavioural heterogeneity in transmission dynamics and intervention uptake. Model calibration was performed using annual syphilis incidence data through a Bayesian framework. Specifically, we employed Hamiltonian Monte Carlo (HMC) via the RStan package (version 2.32.7) in R (version 4.5.0), running six chains of 2,000 iterations each, with the first 1,000 discarded as burn-in. Convergence diagnostics - including traceplots, marginal posterior distributions, effective sample size (ESS), and the Gelman-Rubin (GR) statistic - were used to ensure robust parameter estimation. Posterior estimates of the transition parameters were used to simulate disease dynamics - with and without intervention strategies - using the deSolve package (version 1.40). Simulations were conducted over time from 2026 to 2040 for MSM populations in Singapore and England, based on a sample of 1,000 parameter sets drawn from the joint posterior distribution. Two alternative future scenarios were considered: (A) a stabilized behaviour scenario, in which the time-varying behavioural parameters inferred from historical data (i.e., force of infection 𝜆_j_(𝑡) and asymptomatic screening rate 𝜂_j_(𝑡)) remain constant beyond the final year where data was available; and (B) a continued trend scenario, where these behavioural trends continue to evolve through to 2040 based on past incidence. A detailed introduction to syphilis and doxy-PEP is provided in Supplementary Information Appendix A.

**Fig. 1:**
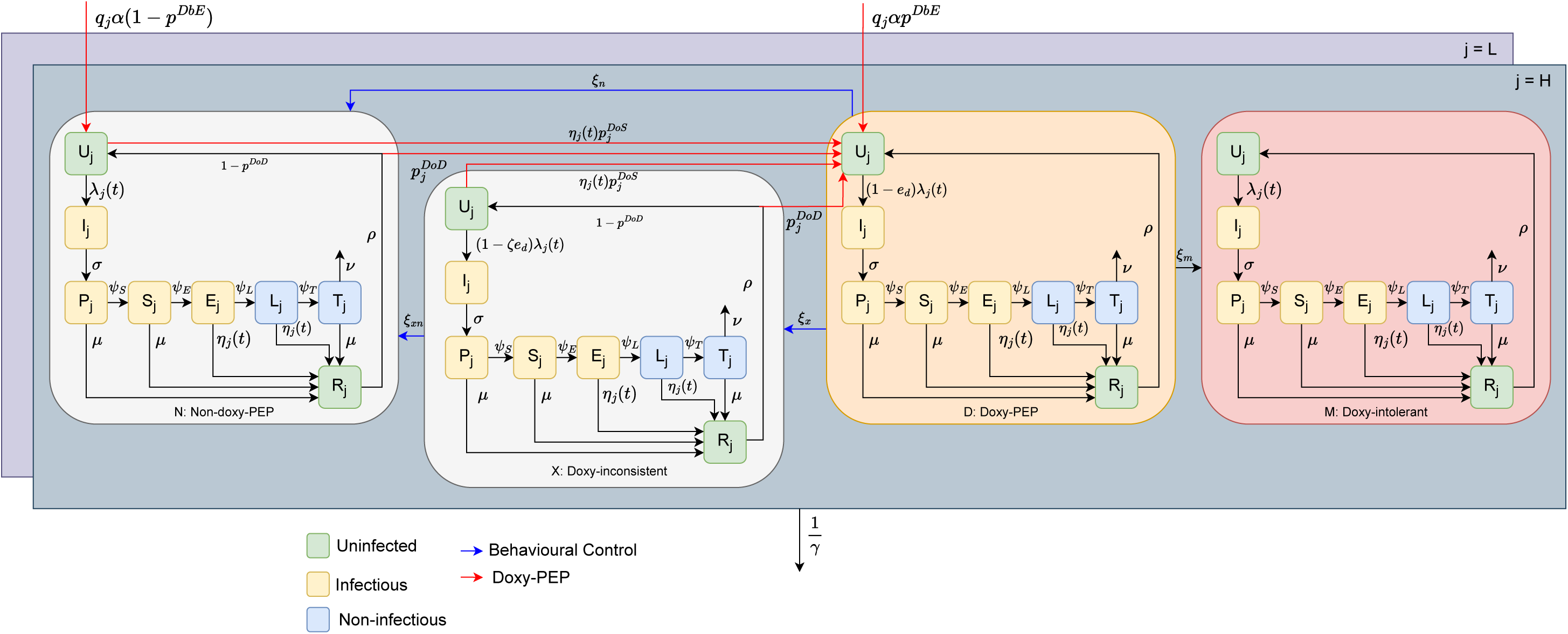
Model architecture of doxy-PEP for syphilis. For each doxy-PEP stratum 𝑖 ∈ {𝑁, 𝑋, 𝐷, 𝑀}, the population is subdivided into compartments representing different stages of syphilis infection. The model distinguishes between individuals with low and high sexual activity levels (𝑗 ∈ {𝐿, 𝐻}), illustrated as the purple (low-activity) and grey (high-activity) layers. Both groups follow the same compartmental structure but may differ in transition rates. For clarity, only the transitions in and out of the high-activity group (upper layer) are shown. For syphilis transmission (refer to stratum 𝑁), individuals enter the sexually active population as uninfected (𝑈_j_). Upon infection, driven by the time-varying force of infection 𝜆_j_(𝑡), they progress through an incubation phase (𝐼_j_) before advancing to successive stages of syphilis infection: primary (𝑃_j_) at a rate 𝜎, secondary (𝑆_j_) at a rate 𝜓_𝑆_, early latent (𝐸_j_) at a rate 𝜓_𝐸_, late latent (𝐿_j_) at a rate 𝜓_𝐿_, and tertiary (𝑇_j_) stages at a rate 𝜓_𝑇_. The primary, secondary, and tertiary stages are symptomatic, prompting individuals to seek treatment and transition to the recovered state (𝑅_j_) at a rate 𝜇. In contrast, early latent and late latent stages are asymptomatic, with infections typically detected through screening, leading to treatment (𝑅_j_) at a screening-dependent rate 𝜂_j_(𝑡). Individuals in the tertiary stage face a risk of mortality, permanently exiting the model at a rate 𝜈. Notably, only the incubation, primary, secondary, and early latent stages are infectious, while the late latent and tertiary stages are non-infectious. Treated individuals are assumed to be cured and return to the uninfected compartment at a rate 𝜌. Individuals may exit the sexually active population due to aging at any stage at a rate 1/𝛾. For the doxy-PEP model, individuals α enter the sexually active population at age 15. With probability 𝑝^𝐷𝑏𝐸^, they initiate doxy-PEP upon entry and enter stratum 𝐷; otherwise, they enter stratum 𝑁. Individuals at stratum 𝐷 may discontinue doxy-PEP and move to stratum 𝑁 at a rate 𝜉_𝑛_; become suboptimally adherent and transition to stratum 𝑋 at a rate 𝜉_𝑥_; or develop intolerance and transition to stratum 𝑀 at a rate 𝜉_𝑚_. Individuals at stratum 𝑋 may also discontinue doxy-PEP and return to stratum 𝑁 at a rate 𝜉_𝑥𝑛_. Individuals at strata 𝑁 and 𝑋 may initiate or reinitiate doxy-PEP following a sexual health clinic visit - either through screening or diagnosis - with probabilities 𝑝^𝐷𝑜𝑆^ and 𝑝^𝐷𝑜𝐷^, respectively, and transition into stratum 𝐷.

Model parameters - both fixed and fitted - were informed by relevant literature or calibrated using behavioural survey data. For instance, bounds on stage-specific syphilis transmission rates, as well as adherence and discontinuation rates, were obtained from existing studies. The estimation of time-varying behavioural parameters - specifically, the force of infection 𝜆_j_(𝑡) and the asymptomatic screening rate 𝜂_j_(𝑡) - was guided by the approach described in [31]. Parameters such as the annual rate of partner change 𝑐_j_and the distribution of MSM across high- and low-risk groups 𝑞_j_ were estimated from survey data. For the survey conducted in Singapore, responses were collected using multiple recruitment strategies to ensure representation across diverse demographic groups. To reach younger participants, the survey was disseminated through popular social media platforms such as Facebook and Telegram. To increase participation among individuals with diverse sexual orientations, the survey was promoted in collaboration with Pink Dot SG - a non-profit movement supporting the LGBTQ community - via their Instagram account and at the offline Pink Dot 16 event. To engage older residents, additional responses were gathered through the survey company IPSOS. Moreover, we characterized doxy-PEP adherence using studies on HIV PrEP, which share similar behavioural challenges related to adherence and discontinuation - this data can be used as a reasonable analogue for informing model assumptions. Key doxy-PEP parameters used in scenario analysis are presented in Table 1. Full details on fixed parameters, prior distributions, posterior summaries, convergence diagnostics, and the system of ordinary differential equations are provided in Supplementary Information Appendices B and C, and source code is available in https://github.com/killingbear999/doxypep_syphilis.

**Table 1:**
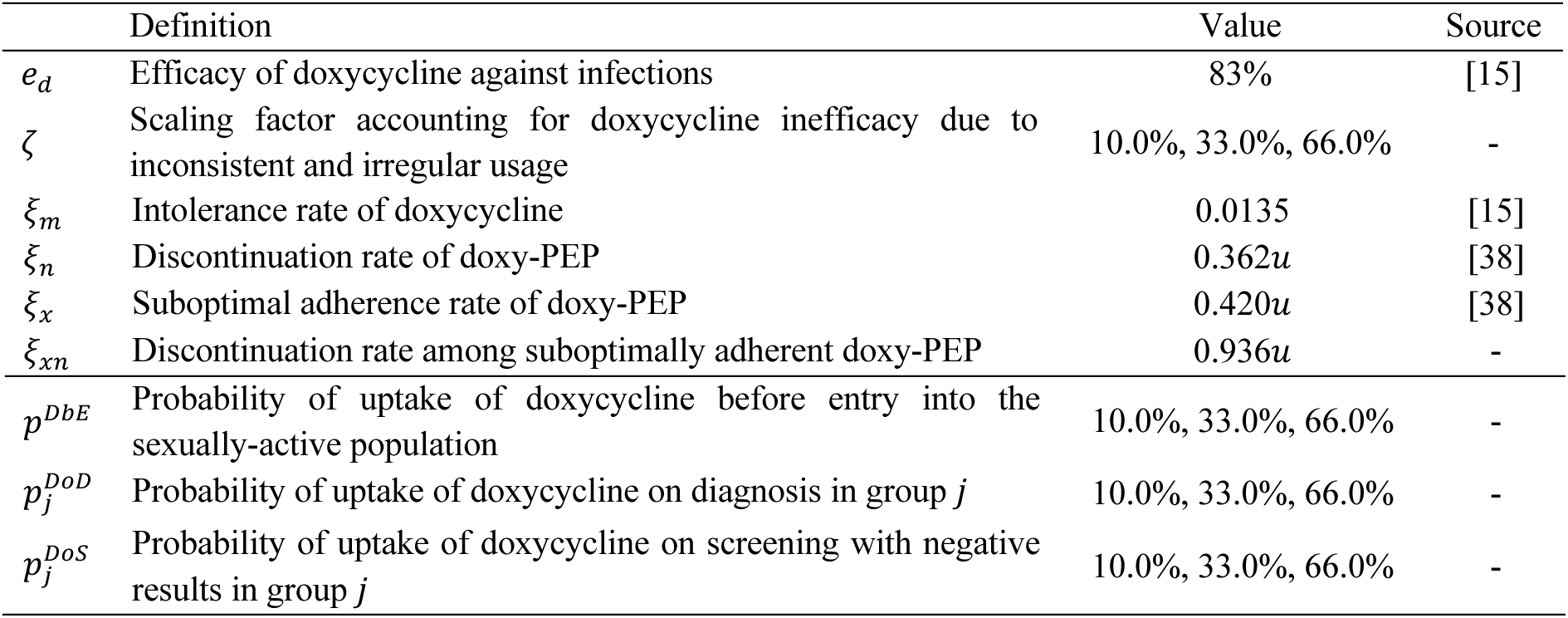
Doxy-PEP parameters used in scenario analysis. We examined three uptake levels: low (10.0%), moderate (33.0%), and high (66.0%), along with three adherence behavioural pattern: low (𝑢 = 2), normal (𝑢 = 1), and high (𝑢 = 0.5). A high adherence behavioural pattern corresponds to low rates of both discontinuation and suboptimal adherence, whereas a low adherence behavioural pattern reflects high rates of both. The different adherence levels are defined relative to HIV PrEP, with ‘normal’ corresponding to typical adherence observed in HIV PrEP studies.

|  | Definition | Value | Source |
| --- | --- | --- | --- |
| $e_d$ | Efficacy of doxycycline against infections | 83% | [15] |
| $\zeta$ | Scaling factor accounting for doxycycline inefficacy due to inconsistent and irregular usage | 10.0%, 33.0%, 66.0% | - |
| $\xi_m$ | Intolerance rate of doxycycline | 0.0135 | [15] |
| $\xi_n$ | Discontinuation rate of doxy-PEP | 0.362u | [38] |
| $\xi_x$ | Suboptimal adherence rate of doxy-PEP | 0.420u | [38] |
| $\xi_{xn}$ | Discontinuation rate among suboptimally adherent doxy-PEP | 0.936u | - |
| $p^{DbE}$ | Probability of uptake of doxycycline before entry into the sexually-active population | 10.0%, 33.0%, 66.0% | - |
| $p_j^{DoD}$ | Probability of uptake of doxycycline on diagnosis in group $j$ | 10.0%, 33.0%, 66.0% | - |
| $p_j^{DoS}$ | Probability of uptake of doxycycline on screening with negative results in group $j$ | 10.0%, 33.0%, 66.0% | - |

Our model comprises four strata based on doxy-PEP usage patterns: (1) Stratum 𝑁 (non-doxy-PEP users): MSM who do not take doxycycline after condomless sex, including those who rely on alternative STI prevention methods such as condom use, as well as individuals with contraindications [32]; (2) Stratum 𝑋 (doxy-inconsistent users): MSM who take doxycycline after condomless sex but experience reduced efficacy due to factors such as alcohol consumption and inconsistent and irregular use (e.g., skipping doses, delayed intake); (3) Stratum 𝐷 (doxy-PEP user): MSM who take doxycycline after condomless sex; (4) Stratum 𝑀 (doxy-intolerant users): MSM who cannot take doxycycline due to medical conditions, including but not limited to a history of allergic reactions to doxycycline or related medications, kidney or liver dysfunction [33], as well as those who initially take doxycycline but discontinue use due to side effects (e.g., gastrointestinal discomfort, photosensitivity, oesophageal irritation) [34]. We incorporate two key assumptions: (1) doxycycline acquired from non-STI-related sources (e.g., primary care, informal sharing, or for unrelated infections like acne or malaria prophylaxis) contributes negligibly to STI prevention [35]; and (2) individuals in strata 𝑁 and 𝑋 only initiate or reinitiate doxy- PEP after attending a sexual health clinic. It is important to note that *T. pallidum* is genetically highly conserved, and despite more than five decades of doxycycline use, no resistance has been reported [36], [37]. Consequently, for syphilis, strain diversity is more relevant for molecular epidemiology than for treatment or prophylaxis planning. Therefore, we assume no evolution of antimicrobial resistance during the projection period.

Doxy-PEP uptake is modelled to occur at five critical decision points: (1) prior to entering the sexually active population (𝑝^𝐷𝑏𝐸^); (2) upon syphilis diagnosis for the high-risk and/or (3) low-risk group (𝑝_j_^𝐷𝑜𝐷^); and (4) after testing negative during routine screening visits for the high-risk and/or (5) low-risk group (𝑝_j_^𝐷𝑜𝑆^), under the *DoS* strategy. By enabling doxy-PEP uptake through these pathways individually or in combination, we evaluate six implementation strategies: (1) Doxy-PEP on Attendance (DoA: 𝑝_j_^𝐷𝑜𝐷^ & 𝑝_j_^𝐷𝑜𝑆^, 𝑗 ∈ {𝐿, 𝐻}): provide doxy-PEP to MSM attending sexual health clinics for STI testing and screening, regardless of their diagnosis; (2) Doxy-PEP on Diagnosis (DoD: 𝑝_j_^𝐷𝑜𝐷^, 𝑗 ∈ {𝐿, 𝐻}): offer doxy-PEP only to MSM diagnosed with syphilis at current visit; (3) Doxy-PEP According to Risk (DaR: 𝑝_j_^𝐷𝑜𝐷^ & 𝑝_𝐻_^𝐷𝑜𝑆^, 𝑗 ∈ {𝐿, 𝐻}): target MSM engaging in high-risk behaviours by offering doxy-PEP to all MSM diagnosed with syphilis and additionally to those with more than 5 partners per year upon attendance at a sexual health clinic regardless of their diagnosis; (4) Doxy-PEP Before Entry (DbE: 𝑝^𝐷𝑏𝐸^): offer doxy-PEP to MSM prior to entering sexually-active age group; (5) Doxy-PEP on Attendance - High-Risk Group (DoA(H): 𝑝_𝐻_^𝐷𝑜𝐷^ & 𝑝_𝐻_^𝐷𝑜𝑆^): provide doxy-PEP to high-risk MSM attending sexual health clinics only; (6) Doxy-PEP on Diagnosis - High-Risk Group (DoD(H): 𝑝_𝐻_^𝐷𝑜𝐷^): provide doxy-PEP to high-risk MSM diagnosed with syphilis only. This framework, combined with sensitivity analyses on key parameters, enables assessment of each strategy’s impact under varying behavioural patterns, including screening frequency, uptake rate, adherence level, and discontinuation rate, offering critical insights for designing efficient and equitable intervention policies.

### c. Performance metrics

To compare the impact and efficiency of different strategies under various scenario settings, we define and quantify key outcomes as follows: (1) Impact: the total number of syphilis cases averted over a 15-year projection period; and (2) Efficiency: the total number of cases averted per doxy-PEP prescription over the same period. To gain deeper insights into transmission dynamics and to inform targeted detection and intervention strategies, we also: (3) estimate the total number of averted syphilis cases stratified by stage - primary, secondary, other (i.e., early latent, late latent, and tertiary), and diagnosed; (4) track the annual number of syphilis cases; (5) monitor the annual number of susceptible MSM in the high-risk group; and (6) infer the force of infection across different countries and epidemiological settings. A comprehensive list of formulas used to compute each metric is provided in Supplementary Information Appendices C and D.

## 3. Results

In this section, we present the results for Singapore’s main scenario (i.e., assuming all excess male syphilis diagnoses represent infections in MSM) and for England, under the assumption that inferred time-varying behavioural trends stabilize. The conclusions drawn from these results are robust to all scenario settings we consider, including Singapore’s lower and upper bound incidence scenarios, and the scenario where behavioural trends are assumed to continue at their inferred level through to 2040, based on historical incidence. Full results for all scenarios and the sensitivity analyses are provided in Supplementary Information Appendix D.

### a. Model calibration and estimated force of infection

Fig. 2 shows that the observed annual numbers of diagnosed syphilis cases predominantly lie within the interquartile range of the posterior predictions from our calibrated model for both Singapore and England. Moreover, convergence diagnostics (see Supplementary Information Appendix B) - including traceplots, marginal posterior distributions, ESS, and the GR statistic - confirm that samples which form the posterior distribution have converged. This indicates that the calibration process successfully captures the slightly increasing trend in Singapore and the steeper upward trend observed in England.

**Fig. 2:**
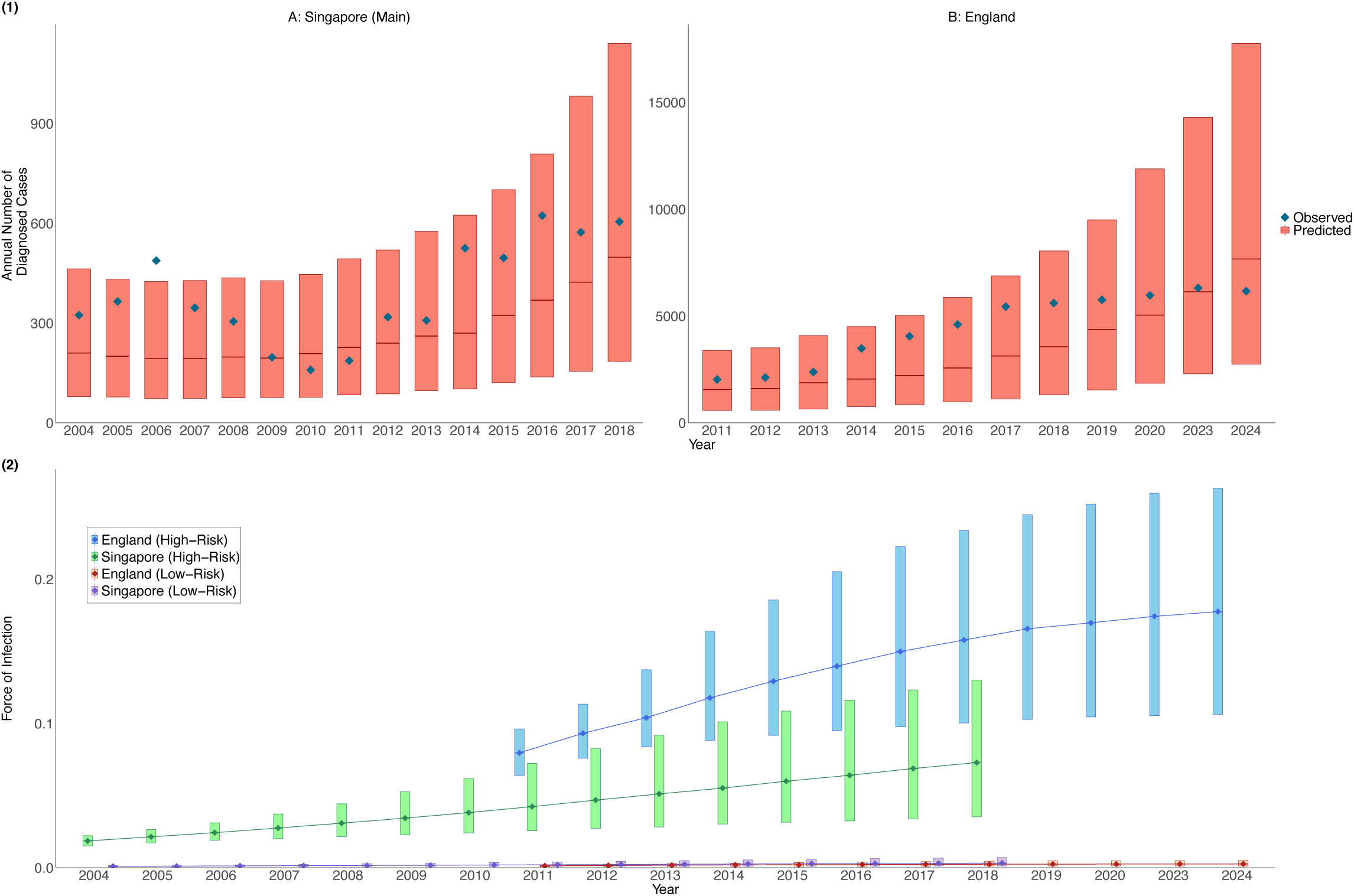
Panel 1 shows simulated epidemic trajectories and observed syphilis incidence among MSM in England and Singapore (assuming all excess male syphilis diagnoses represent infections in MSM). Each box represents the interquartile range of the posterior distribution, with the median indicated by a horizontal line. Observed annual syphilis incidence data are shown as diamonds. Panel 2 shows force of infection estimates based on Bayesian calibration for England and Singapore, stratified by high-risk and low- risk groups. For each box, the median is represented by a diamond, while the interquartile range is indicated by the upper and lower edges of the box.

The model also inferred an increasing force of infection in both settings, consistent with the rising number of diagnosed cases. Notably, the estimated force of infection is substantially higher for the high-risk MSM group compared to the low-risk group in both countries, highlighting the disproportionate contribution of high-risk individuals to overall transmission dynamics (see Fig. 2). Additionally, the force of infection for high-risk MSM in England is consistently higher than that in Singapore across the calibration period, whereas for low-risk MSM, Singapore’s force of infection is consistently higher than England’s. It is important to note that the force of infection and the annual incidence rate, while related, are not equivalent measures. Force of infection describes the per-susceptible hazard of acquiring infection within a given time period, whereas the incidence rate represents the number of new detected cases relative to the total population. Mathematically, incidence is the product of force of infection and the proportion of the population that remains susceptible. Therefore, settings with high testing coverage or a greater proportion of susceptible individuals may exhibit a higher observed incidence rate even if the underlying force of infection is lower. For example, Singapore’s syphilis incidence per 100,000 population is higher than England’s, yet our model estimates a lower force of infection in high-risk MSM in Singapore, likely reflecting differences in population structure, routine screening coverage, and the proportion of infections detected through intensive case finding.

### b. Potential impact and efficiency of doxy-PEP

Fig. 3 illustrates the projected long-run impact of alternative doxy-PEP strategies on syphilis incidence among MSM in Singapore and England. Across both settings, all evaluated strategies are projected to reduce syphilis incidence over time, with greater reductions achieved the longer doxy-PEP is implemented. Strategies targeting high-risk MSM at diagnosis (DoD(H)) demonstrate a moderate but sustained impact, closely mirroring the results of DoD overall. In contrast, broader strategies such as DoA, DoA(H), and DaR yield the greatest total reductions in incidence over the 15-year horizon. Notably, DbE - which targets MSM before they enter the sexually active population - produces the smallest overall reduction in incidence across years.

**Fig. 3:**
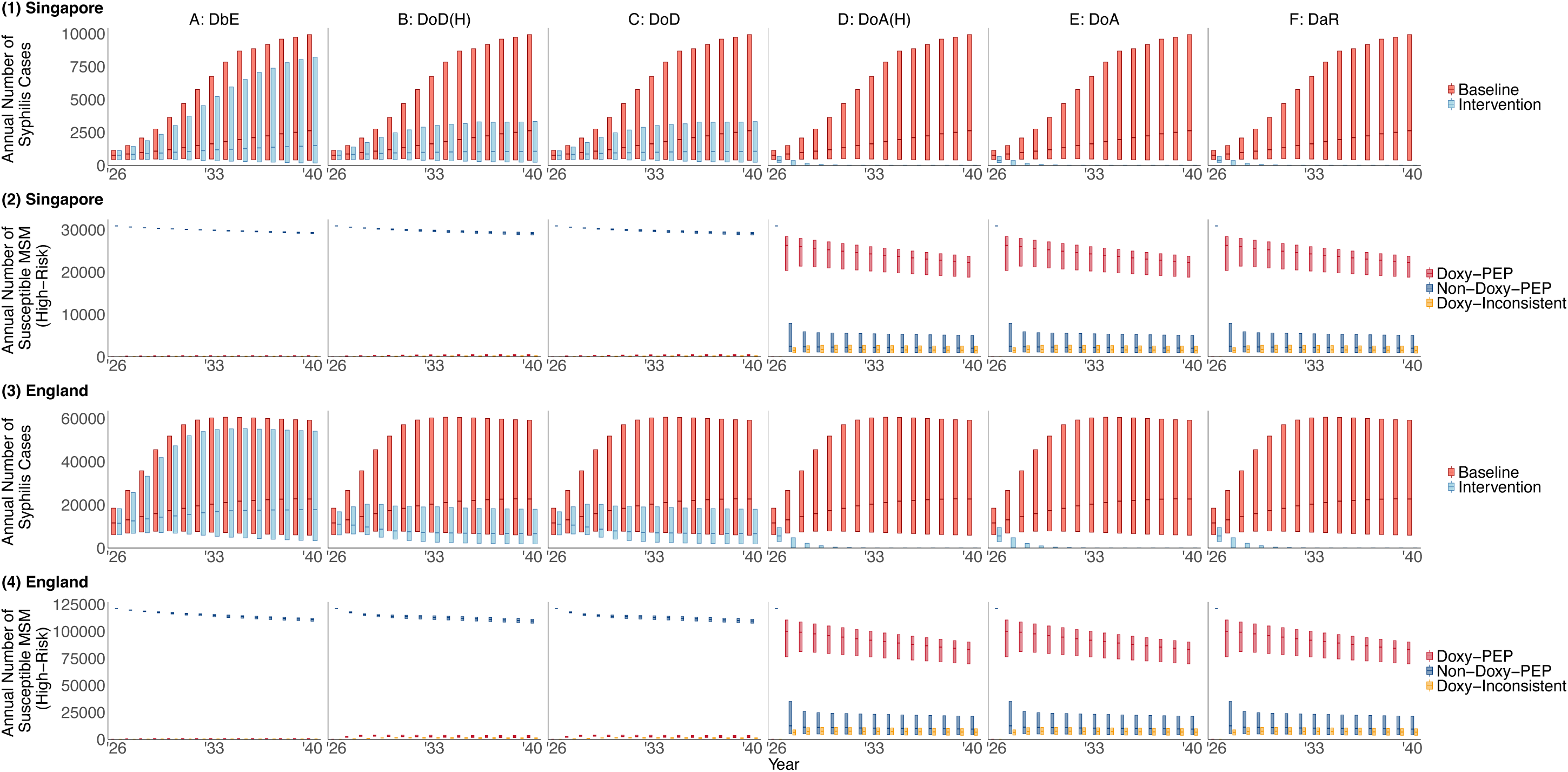
From left to right, the doxy-PEP strategies are: doxy-PEP on entry (DbE), doxy-PEP on diagnosis for the high-risk group (DoD(H)), doxy-PEP on diagnosis (DoD), doxy-PEP on attendance to the high-risk group (DoA(H)), doxy-PEP on attendance (DoA), and doxy-PEP according to risk (DaR). The presented Singapore scenario assumes stabilization of inferred time-varying behavioural trends, with the MSM incidence of syphilis assumed to be male syphilis cases minus the female syphilis cases. Panels (1) and (3) show the annual number of syphilis cases under intervention (uptake rate of 33.0%) and baseline scenarios for Singapore and England, respectively, where protection is reduced to 33.0% of the baseline efficacy for suboptimal adherence strata (i.e., ζ = 33.0%). Each boxplot shows the median (central line) and the 95% credible interval (box bounds). Panels (2) and (4) present the annual number of susceptible MSM at high-risk group for Singapore and England, respectively. Each boxplot shows the median (central line) and the interquartile range (box bounds).

Table 2 and Table 3 (all results are reduced to the nearest 100 to avoid spurious accuracy) summarize the estimated population-level impact and implementation efficiency of different doxy-PEP prescribing strategies among MSM in Singapore and England, assuming moderate uptake (33.0%), normal adherence patterns, and stabilization of inferred time-varying behavioural trends. Overall, our results show that any strategy including the low-risk group is substantially less efficient. For example, comparing DoD(H) with DoD demonstrate that adding the low-risk group yields negligible additional benefit in terms of total cases averted (Singapore: DoD(H) 10000 [95% CrI 1100 - 53200] vs. DoD 10100 [1200 - 53200]; England: both DoD(H) and DoD 165000 [49300 - 503700]), but markedly reduces efficiency per prescription (Singapore: DoD(H) 2.50 [0.68 - 5.94] vs DoD 2.08 [0.54, 4.88]; England: DoD(H) 4.60 [2.12 - 7.79] vs DoD 4.19 [1.92 - 7.01]). Similarly, the comparison among DoA(H), DoA, and DaR demonstrates that extending doxy-PEP to low-risk MSM after negative syphilis screening provides minimal additional population-level impact (Singapore: all three report 24700 [6800 - 93100]; England: all three report 279800 [109100 - 724000]), yet greatly increases the number of prescriptions required per case averted (Singapore: DoA(H) 0.08 [0.02 - 0.63], DoA 0.02 [0.00 - 0.23], DaR 0.08 [0.02 - 0.63]; England: DoA(H) 0.21 [0.07 - 1.38], DoA 0.04 [0.01 - 0.46], DaR 0.21 [0.07 - 1.38]). These findings reinforce that including low-risk MSM leads to inefficiency without meaningful additional population-health benefit. The negligible impact of targeting the low-risk group also explains why DaR and DoA(H) produce nearly identical outcomes, reflecting the very low number of diagnoses in this group - likely due to the low force of infection from calibration. This pattern remains consistent across different scenarios (see Supplementary Information Appendix D).

**Table 2:** Estimated impact and efficiency of doxy-PEP strategies among MSM (uptake rate of 33.0%) with a normal adherence behavioural pattern (i.e., u = 1), where protection is reduced to 33.0% of the baseline efficacy for suboptimal adherence strata (i.e., ζ = 33.0%), assuming stabilization of inferred time-varying behavioural trends. Results are presented as median (95% credible interval), assuming a high asymptomatic screening rate (taken from our calibration, aligned with CDC-recommended levels [39]).

|  | Singapore (main scenario) |  |  | England |  |  |
| --- | --- | --- | --- | --- | --- | --- |
|  | Total number of averted cases, thousands | Number of doxy-PEP prescriptions, thousands | Number of averted cases per prescription | Total number of averted cases, thousands | Number of doxy-PEP prescriptions, thousands | Number of averted cases per prescription |
| DbE | 7.3<br>(1.6, 19.2) | 12.5<br>(12.5, 12.5) | 0.59<br>(0.13, 1.53) | 44.5<br>(20.6, 74.6) | 59.4<br>(59.4, 59.4) | 0.75<br>(0.35, 1.26) |
| DoD(H) | 10.0<br>(1.1, 53.2) | 4.0<br>(1.6, 9.8) | 2.50<br>(0.68, 5.94) | 165<br>(49.3, 503.7) | 35.6<br>(17.8, 77.2) | 4.60<br>(2.12, 7.79) |
| DoD | 10.1<br>(1.2, 53.2) | 4.8<br>(2.0, 12.8) | 2.08<br>(0.54, 4.88) | 165<br>(49.3, 503.7) | 38.4<br>(19.9, 87.7) | 4.19<br>(1.92, 7.01) |
| DoA(H) | 24.7<br>(6.8, 93.1) | 358.7<br>(46.3, 659.4) | 0.08<br>(0.02, 0.63) | 279.8<br>(109.1, 724.0) | 1308.0<br>(215.8, 2336.9) | 0.21<br>(0.07, 1.38) |
| DoA | 24.7<br>(6.8, 93.1) | 1390.7<br>(130.5, 2390.9) | 0.02<br>(0.00, 0.23) | 279.8<br>(109.2, 724.0) | 6919.2<br>(724.1, 11785.3) | 0.04<br>(0.01, 0.46) |
| DaR | 24.7<br>(6.8, 93.1) | 358.7<br>(46.4, 659.4) | 0.08<br>(0.02, 0.63) | 279.8<br>(109.1, 724.0) | 1308.1<br>(216.5, 2337.1) | 0.21<br>(0.07, 1.38) |

**Table 3:** Estimated impact of doxy-PEP strategies (total number of averted cases by stage) among MSM (uptake rate of 33.0%) with a normal adherence behavioural pattern (i.e., 𝑢 = 1), where protection is reduced to 33.0% of the baseline efficacy for suboptimal adherence strata (i.e., 𝜁 = 33.0%), assuming stabilization of inferred time-varying behavioural trends. Results are presented as median (95% credible interval), assuming a high asymptomatic screening rate (taken from our calibration, aligned with CDC-recommended levels [39]).

|  | Singapore (main scenario) |  |  |  | England |  |  |  |
| --- | --- | --- | --- | --- | --- | --- | --- | --- |
|  | Primary stage,<br>thousands | Secondary stage,<br>thousands | Other stages | Diagnosed,<br>thousands | Primary stage,<br>thousands | Secondary stage,<br>thousands | Other stages | Diagnosed,<br>thousands |
| DbE | 7.4<br>(1.6, 17.2) | 0.3<br>(0.1, 0.8) | 3<br>(0, 42) | 7.3<br>(1.6, 19) | 44.4<br>(20.5, 74.6) | 2.3<br>(0.5, 7.8) | 25<br>(2, 260) | 44.0<br>(20.3, 74.2) |
| DoD(H) | 10.1<br>(1.1, 53.6) | 0.5<br>(0, 4.0) | 5<br>(0, 83) | 10.0<br>(1.1, 53.1) | 164.8<br>(48.6, 507.0) | 8.4<br>(1.3, 40.9) | 90<br>(7 - 1161) | 164.2<br>(48.4, 505.8) |
| DoD | 10.1<br>(1.2, 53.8) | 0.5<br>(0, 4.0) | 5<br>(0, 83) | 10.0<br>(1.1, 53.3) | 164.8<br>(48.6, 507.0) | 8.4<br>(1.3, 41.0) | 90<br>(7 - 1162) | 164.3<br>(48.4, 505.8) |
| DoA(H) | 24.8<br>(6.8, 94.2) | 1.2<br>(0.2, 6.7) | 12<br>(1, 167) | 24.7<br>(6.8, 93.6) | 276.7<br>(109.5, 724.2) | 14.2<br>(2.9, 64.0) | 155<br>(14 - 1815) | 275.9<br>(108.9, 722.3) |
| DoA | 24.8<br>(6.9, 94.2) | 1.2<br>(0.2, 6.7) | 12<br>(1, 167) | 24.7<br>(6.8, 93.6) | 276.7<br>(109.5, 724.2) | 14.2<br>(2.9, 64.0) | 155<br>(14 - 1815) | 276.0<br>(108.9, 722.3) |
| DaR | 24.8<br>(6.8, 94.2) | 1.1<br>(0.2, 6.6) | 12<br>(1, 167) | 24.7<br>(6.8, 93.6) | 276.7<br>(109.5, 724.2) | 14.2<br>(2.9, 64.0) | 155<br>(14 - 1815) | 275.9<br>(108.9, 722.3) |

Moreover, across settings, DoD(H) consistently achieved the best balance between impact and efficiency. In contrast, broader strategies such as DoA, DoA(H), and DaR achieved greater total reductions in syphilis incidence, but were much less efficient. By comparison, DbE shows a more moderate impact (averting 7300 [1600 - 19200] total cases in Singapore and 44500 [20600 - 74600] in England), and moderate efficiency (0.59 [0.13 - 1.53] cases averted per prescription in Singapore and 0.75 [0.35 - 1.26] in England). This indicates that DbE is neither as impactful as the broader coverage strategies (DoA, DoA(H), DaR) in reducing total incidence nor as efficient as more targeted approaches like DoD(H) and DoD. These findings reinforce the importance of targeting high-risk MSM at diagnosis to maximize impact and efficiency while minimizing unnecessary prescriptions.

Stage-specific results (Table 3) show that the majority of infections averted occur at the primary syphilis stage. For example, under DoD(H) in Singapore, most averted cases are in the primary stage (10100 [1100 - 53600], compared to 500 [0 - 4000] at the secondary stage and only 5 [0 - 83] at later stages (i.e., early and late latent, and tertiary stage)). Similar patterns hold in England (DoD(H) averts an estimated 164800 [48600 - 507000] primary stage cases, 8400 [1300 - 40900] secondary stage cases, and only 90 [7 - 1161] at later stages).

### c. Impact of uptake rate, screening frequency and adherence behavioural patterns on prescription strategies

Strategy selection should weigh both efficiency - defined as the number of averted cases per prescription - and overall population-level impact - defined as the total number of averted cases. Three key behavioural parameters strongly influence the impact and efficiency of doxy-PEP prescribing strategies: uptake rate, screening frequency, and adherence patterns (including the suboptimal adherence rate among fully adherent individuals, the discontinuation rate among fully adherent individuals, and the discontinuation rate among suboptimally adherent individuals). It is important to note that uptake rate and adherence behavioural patterns are specific to doxy-PEP implementation and therefore do not influence the no-intervention baseline. In contrast, screening frequency directly affects both the intervention and no-intervention scenarios. This is because screening determines how many infections are detected and treated through routine care, independently of whether doxy-PEP is offered. Higher asymptomatic screening rates can reduce underlying transmission by facilitating earlier diagnosis and treatment, thereby lowering the baseline incidence of syphilis. As a result, the incremental impact of adding doxy- PEP will differ depending on the background screening capacity. This highlights the need to consider interactions between behavioural interventions and existing public health infrastructure when evaluating the potential effectiveness and efficiency of doxy-PEP strategies. We begin by analysing how variations in uptake rate and screening frequency affect the impact and efficiency of doxy-PEP strategies, while keeping adherence behavioural patterns fixed at normal levels.

Fig. 4 illustrates the impact and efficiency of different doxy-PEP strategies in Singapore and England under varying uptake levels and asymptomatic screening rates. Results for both settings show broadly consistent patterns. When holding uptake rate constant and comparing screening scenarios, nearly all strategies exhibit higher estimated impact and efficiency under a low asymptomatic screening rate. This is because lower screening results in fewer diagnosed and promptly treated infections, resulting in more infectious individuals and higher ongoing transmission, which raises the baseline incidence and therefore the absolute number of cases that can be averted by doxy-PEP. This underscores the importance of maintaining high asymptomatic screening rates as a primary means of controlling syphilis transmission. Notably, even under low screening conditions (where baseline transmission is more severe), doxy-PEP demonstrates substantial potential to reduce incidence, highlighting its added value when routine screening coverage is suboptimal.

**Fig. 4:**
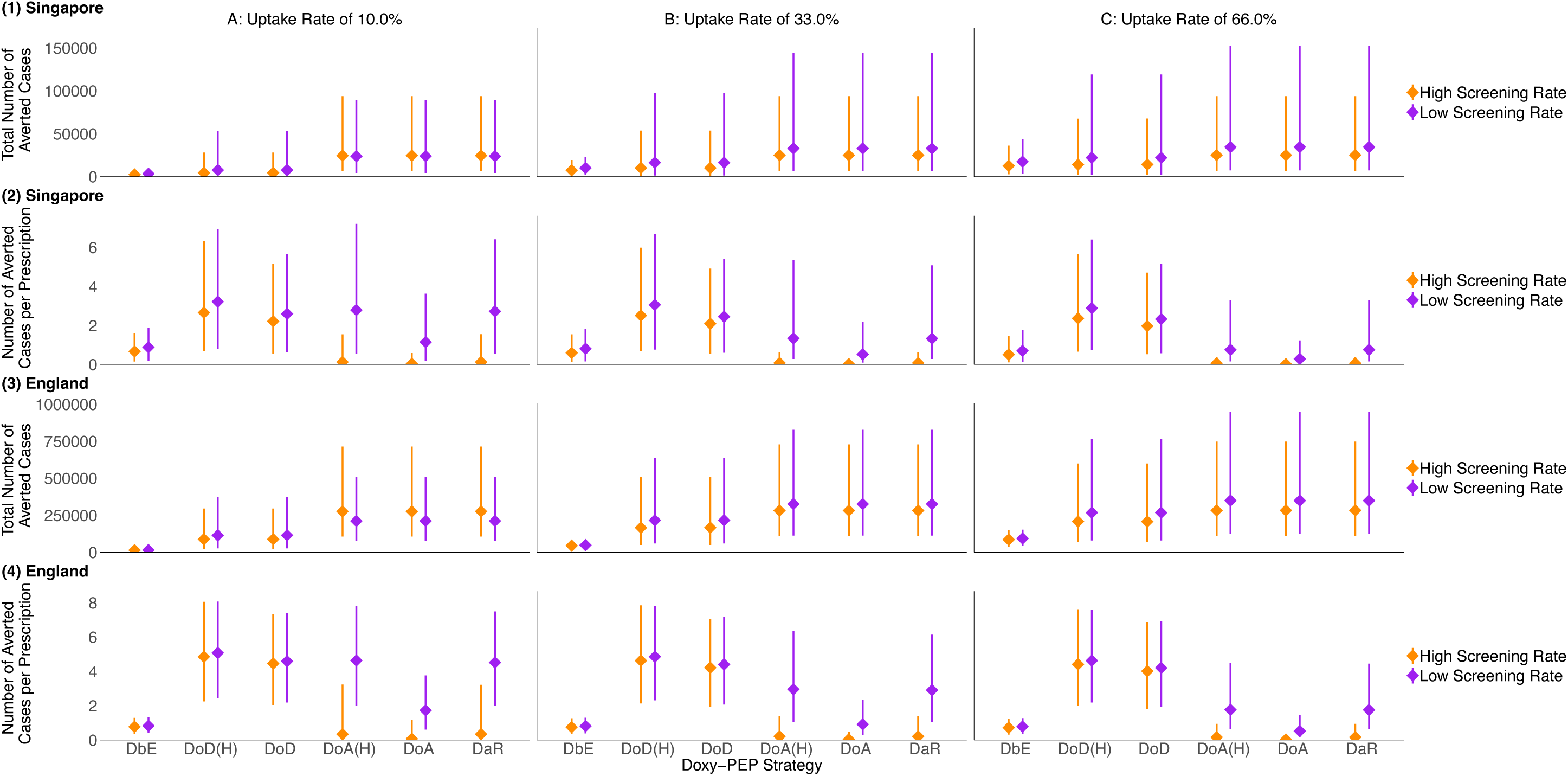
From left to right, the assumed uptake rates are 0.10, 0.33, and 0.66. Each point-range plot displays the median estimate (central diamond) and the 95% credible interval (lines), under the assumption of suboptimal adherence that reduces protection to 33.0% of the baseline efficacy (i.e., 𝜁 = 33.0%). The main scenario assumes stabilization of inferred time-varying behavioural trends, with all males not engaging in sex with females classified as MSM. Panels (1) and (3) show the total number of averted cases comparing low vs. high asymptomatic screening rates across doxy-PEP strategies for Singapore and England, respectively. Panels (2) and (4) show the number of averted cases per prescription, comparing low vs. high asymptomatic screening rates across doxy-PEP strategies for Singapore and England, respectively.

Under high screening conditions, across all uptake levels, DoD(H) consistently achieves the highest efficiency (yielding 2.50 [0.68 - 5.94] averted cases per prescription in Singapore and 4.60 [2.12 - 7.79] in England for an uptake rate of 0.33), while maintaining a substantial total number of averted cases (10000 [1100 - 53200] in Singapore and 165000 [49300 - 503700] in England). This makes DoD(H) the most favourable strategy in high screening settings. Although broader approaches such as DoA(H), DoA, and DaR generate larger absolute reductions (e.g., DoA(H) averts 24700 [6800 - 93100] cases in Singapore and 279800 [109100 - 724000] in England for an uptake rate of 0.33), these gains come at the cost of markedly lower efficiency. DoA(H), for instance, achieves only 0.08 (0.02 - 0.63) averted cases per prescription in Singapore and 0.21 (0.07 - 1.38) in England. Such low efficiency raises concerns about unnecessary prescribing, increased resource burden, and the heightened risk of antimicrobial resistance strain emergence. In contrast, under low screening conditions, DoA(H) strikes a better balance by achieving both relatively high efficiency and substantial impact (yielding 1.32 [0.28 - 5.33] averted cases per prescription and 32600 [6800 - 143000] total averted cases in Singapore, and 2.93 [1.04 - 6.32] cases per prescription and 323700 [111800 - 821500] total averted cases in England at an uptake rate of 0.33). In this setting, DoA(H) slightly outperforms DaR. While DoD(H) remains the most efficient per prescription under low screening (3.04 [0.76 - 6.63] in Singapore and 4.82 [2.30 - 7.76] in England), its lower overall impact (16200 [1400 - 96500] in Singapore and 214500 [58900 - 632900] in England) limits its standalone appeal. Therefore, when screening resources are constrained, DoA(H) may offer the most practical and favourable balance between impact and efficiency for real-world implementation.

Next, we examine how uptake rate and adherence behavioural patterns affect the impact and efficiency of doxy- PEP prescribing strategies, holding screening frequency constant at a CDC-recommended level (as calibrated in our model). Fig. 5 presents results for Singapore and England across varying uptake rates and adherence patterns. For strategies such as DbE, DoD(H), and DoD, higher adherence - defined by greater consistency and lower suboptimal adherent and discontinuation rates - consistently translates into greater impact and efficiency, regardless of uptake. For example, at an uptake rate of 0.33, DoD(H) in Singapore is projected to avert 6900 (700 - 40100) cases under a low adherence pattern versus 13100 (1700 - 64100) under a high adherence pattern. Correspondingly, efficiency increases from 1.43 (0.38 - 3.35) to 4.00 (1.09 - 9.90) averted cases per prescription. A similar trend holds in England, where DoD(H) averts 124800 (34300 - 398300) cases under low adherence and 199600 (63300 - 573800) under high adherence, with efficiency rising from 2.56 (1.20 - 4.26) to 7.77 (3.45 - 13.61) cases per prescription. This pattern highlights the critical role of sustained, proper use for realising the full benefit of more targeted prescribing approaches. In contrast, for strategies such as DoA(H), DoA, and DaR, the impact remains largely unchanged across different adherence scenarios - again demonstrating a saturation effect. For instance, with an uptake rate of 0.33, Singapore’s DoA(H) averts 24600 (6800 - 93100) cases under low adherence versus 24800 (6900 - 93100) under high adherence; similar stability is seen in England, where DoA(H) averts 278300 (107500 - 724000) under low adherence and 280000 (109600 - 738500) under high adherence. This occurs because, under these strategies, a large proportion of the target population enters the protected stratum even with moderate adherence, meaning that further improvements in adherence behaviour contribute little additional benefit. This finding underscores the risk of over-prescription and diminishing returns when low-risk groups are included. Notably, across all adherence and uptake scenarios, DoD(H) consistently achieves the highest efficiency while delivering a substantial total number of averted cases, making it the most robust strategy under varying behavioural conditions.

**Fig. 5:**
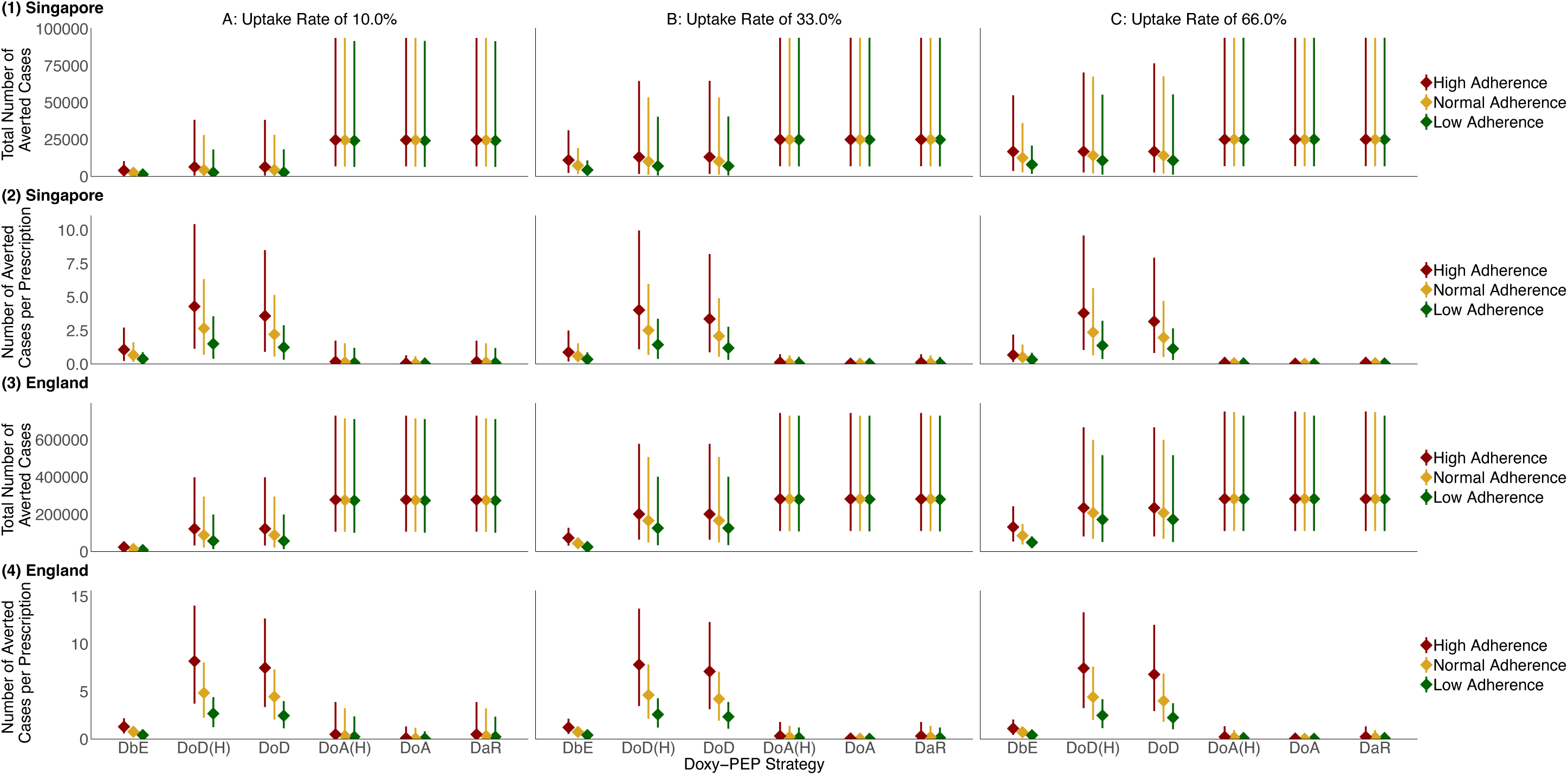
From left to right, the assumed uptake rates are 0.10, 0.33, and 0.66. Each point-range plot displays the median estimate (central diamond) and the 95% credible interval (lines), under the assumption of suboptimal adherence that reduces protection to 33.0% of the baseline efficacy (i.e., 𝜁 = 33.0%). The Singapore main scenario assumes stabilization of inferred time-varying behavioural trends, with all males not engaging in sex with females classified as MSM. Panels (1) and (3) show the total number of averted cases comparing low vs. normal vs. high adherence behavioural patterns across doxy-PEP strategies for Singapore and England, respectively. Panels (2) and (4) show the number of averted cases per prescription, comparing low vs. normal vs. high adherence behavioural patterns across doxy-PEP strategies for Singapore and England, respectively.

Lastly, regardless of uptake rate, screening frequency, or adherence behavioural patterns, our results in Fig. 4 and Fig. 5 consistently show that DoD(H) and DoD achieve nearly identical total impact but that DoD is notably less efficient per prescription. Similarly, DoA(H), DoA, and DaR yield comparable overall reductions in syphilis incidence, yet DoA is substantially less efficient. These consistent patterns further reinforce the conclusion that including low-risk MSM adds little additional benefit while significantly reducing implementation efficiency, highlighting the importance of targeted prescribing strategies.

### d. Model calibration and sensitivity analyses

As expected in Bayesian calibration of time-series models, the widening of the interquartile range toward the end of the calibration period reflects the increasing uncertainty from parameter estimates propagating through the dynamic system (see Fig. 2 and Supplementary Information Appendix B). This is typical for Bayesian inference of time-varying processes, as the model describes the observed data probabilistically rather than deterministically. Rather than forcing the fit to pass through each point exactly, the model represents a plausible generative process consistent with both the data and the underlying epidemiological mechanisms. Moreover, it is important to highlight that calibration to other Singapore-specific incidence scenarios (i.e., lower and upper bound estimates) and sensitivity analyses on key parameters (e.g., the scaling factor 𝜁 accounting for reduced doxycycline efficacy due to inconsistent or irregular use) produced results that were consistent with the final analyses and conclusions (see Supplementary Information Appendix D).

## 4. Discussion

Our study provides new evidence to guide the sustainable implementation of doxy-PEP for syphilis prevention among the MSM population. Results presented corroborate with prior evidence in open-label randomized trials [14], [15], [20], [21] and modelling studies which found that doxy-PEP can be efficacious and/or effective in preventing syphilis transmission [25], at both trial and population levels. Importantly, our study adds to the existing evidence base by calibrating a high-resolution, risk-stratified, behavioural transmission-dynamic framework to real-world epidemiological and behavioural data across Singapore and England. This is of importance given rising population-wide incidence of syphilis in both countries [2], [3], [40], doxy-PEP has potential to reduce incidence of syphilis at the population level [23].

Moreover, our work enables quantification of long-run population impact, efficiency, and transmission dynamics under real-world behavioural patterns. We quantify the efficiency and impact of different prescription strategies which can be realistically implemented at the population level, and examine how behavioural factors, such as adherence, screening and uptake will impact potential public health programmes across disparate epidemiological settings.

We found that strategies targeting high-risk MSM - particularly prescribing doxy-PEP at diagnosis (DoD(H)) - consistently deliver the best balance of substantial population-level impact and high efficiency for syphilis prevention. Our results underscore a critical trade-off: looking at total impact alone does not provide a sufficient basis for recommending an optimal strategy. While broad-targeting strategies can achieve greater absolute reductions, they do so at a greater cost, meaning substantially more prescriptions are required to avert each additional case. As the force of infection for low-risk MSM is historically far smaller versus the high-risk MSM group in both contexts (see Fig. 2), it implies that including the low-risk group in doxy-PEP prescribing strategies would have limited impact on population-level transmission reduction. More importantly, including low-risk MSM greatly increases the number of prescriptions needed, raising concerns about overuse of antibiotics and resource burden. This is relevant given co-transmission of syphilis with other STIs (e.g., gonorrhoea), limited effectiveness of doxy-PEP in preventing gonorrhoea, and emerging antibiotic resistance for gonorrhoeal infections globally [41]; this, in turn, raises the concern that doxy-PEP could accelerate emergence of antibiotic resistance in *N gonorrhoeae*, when the population perspective is considered [42]. As such, it is crucial for implementation strategies of doxy-PEP to strike the balance between impact/effectiveness and potential over-prescription.

Our findings were robust to variations in uptake and adherence behaviour, reinforcing that asymptomatic screening rate is the primary driver in selecting an optimal strategy. Where asymptomatic screening rates are expected to be high - as in Singapore [43] and England [44] - DoD(H) should be prioritised. In lower screening settings, broader approaches such as DoA(H) can achieve greater reductions in incidence with acceptable efficiency, highlighting the importance of tailoring strategies to local capacity and behavioural contexts.

A major strength of our study is the use of epidemiological and behavioural data streams to comprehensively calibrate our high-resolution modelling framework using Bayesian methods to account for uncertainty in behavioural and epidemiological parameters, including time-varying changes in sexual behaviour and testing patterns. Our approach directly integrates real-world surveillance data from Singapore and England, incorporates local sexual network characteristics, and explicitly considers adherence dynamics - an aspect especially critical for event-driven prevention strategies like doxy-PEP, which rely more heavily on consistent user behaviour than vaccines do. We modelled realistic implementation approaches for doxy-PEP which can be directly applied to most strata. At baseline, as the model incorporated behavioural parameters, we could also easily provide policymakers with a broad decision space for the optimal doxy-PEP programme, based on local contexts. Our study also incorporates two separate epidemiological contexts (Singapore, England), which reinforces the applicability of the prescription programmes over different trends in incidence.

However, several limitations should be acknowledged. First, estimates of adherence behaviour were informed by HIV PrEP studies, as doxy-PEP-specific adherence data are not yet available; however, these patterns may differ given the distinct dosing regimens (daily for PrEP versus event-driven for doxy-PEP). Second, syphilis incidence among MSM in Singapore was indirectly estimated due to the lack of disaggregated data, although England’s data were directly available. Third, we made simplifying assumptions regarding future trends in transmission probability and asymptomatic screening rates using linear functions. Additionally, we did not incorporate proximal risk factors such as detailed sexual history, nor did we account for network-level risk dynamics in this model. For example, a “low-risk” MSM - defined as having few partners - who contracts syphilis may still face elevated risk of reinfection if their partners belong to higher-risk networks. This nuanced transmission pattern, which could influence the overall force of infection, is not captured in our current framework. Superspreading dynamics, migration effects, and declining birth rates (since we assumed a constant population inflow at age 15, although the number reaching this age may decrease in future years) were also not explicitly modelled. However, these factors are unlikely to materially affect our outcomes, as their impact is expected to be minor relative to the large base population size. Furthermore, we assumed minimal impact from non-clinic doxycycline use and restricted re-initiation to sexual health clinic visits, as relevant real-world data for these pathways are not yet available.

Finally, our model infers that broad-coverage strategies - specifically DoA, DoA(H), and DaR - could virtually eliminate syphilis in the long term in both countries. These strategies achieve near-elimination by ensuring a large proportion of high-risk MSM consistently use doxy-PEP. Given doxy-PEP’s high efficacy (∼80% for fully adherent users), the force of infection declines substantially, interrupting transmission chains and allowing elimination thresholds to be met. Two factors may have driven this outcome. First, hesitancy is modelled as a probability of accepting doxy-PEP each time it is offered (e.g., a 33.0% chance per offer), rather than as a fixed trait of individuals (e.g., 33.0% always accept, 67.0% never do). This distinction matters in scenarios like DoA, DoA(H), and DaR, where high incidence and frequent asymptomatic screening mean individuals receive multiple offers, leading to high cumulative doxy-PEP coverage [45]. Second, our model relies on a relatively simple heterogeneity structure with only two risk groups. In reality, increasing the number of modelled risk strata which captures very high-activity individuals may raise disease transmissibility within the core group, and can render the intervention inefficacious in these strata [46].

Overall, our findings highlight the importance of aligning doxy-PEP prescribing with behavioural risk and screening capacity to maximise public health benefits while minimising unnecessary antibiotic exposure. Specifically, our results support prioritising targeted strategies such as prescribing doxy-PEP to high-risk MSM (five or more sexual partners per year) at syphilis diagnosis in settings with high screening coverage, like Singapore and England. Maintaining high screening coverage remains essential for sustainable syphilis control. Future work should continue to refine behavioural assumptions and incorporate additional empirical data on adherence patterns specific to doxy-PEP use, including discontinuation and irregular usage. It will also be important to evaluate equity considerations to ensure that implementation strategies are accessible and acceptable to those populations at highest risk, thereby maximising public health impact while minimising disparities. In addition, future studies should conduct robust cost-effectiveness and cost-utility analyses to quantify the economic value and broader health system implications of different prescribing strategies. By integrating these dimensions, health systems can design targeted and efficient prophylaxis programmes that sustainably reduce syphilis transmission and antimicrobial resistance risks in at-risk communities.

## Data Availability

All data produced in the present work are contained in the manuscript.

## Reference

[1] “Global Sexually Transmitted Infections Programme,” World Health Organization. Accessed: June 29, 2025. [Online]. Available: https://www.who.int/teams/global-hiv-hepatitis-and-stis-programmes/stis/strategic-information

[2] “[ARCHIVED CONTENT] UK Government Web Archive - The National Archives.” Accessed: June 25, 2025. [Online]. Available: https://webarchive.nationalarchives.gov.uk/ukgwa/20170517101539/https://www.gov.uk/government/statistics/sexually-transmitted-infections-stis-annual-data-tables

[3] “Sexually transmitted infections (STIs): annual data,” GOV.UK. Accessed: June 25, 2025. [Online]. Available: https://www.gov.uk/government/statistics/sexually-transmitted-infections-stis-annual-data-tables

[4] “The Communicable Disease Surveillance in Singapore 2004,” Ministry of Health. Accessed: Feb. 08, 2025. [Online]. Available: https://www.moh.gov.sg/others/resources-and-statistics/reports-communicable-disease-surveillance-in-singapore-2004/

[5] “The Communicable Diseases Surveillance in Singapore 2018,” Ministry of Health. Accessed: Feb. 08, 2025. [Online]. Available: https://www.moh.gov.sg/others/resources-and-statistics/the-communicable-diseases-surveillance-in-singapore-2018/

[6] “COMMUNICABLE DISEASES SURVEILLANCE IN SINGAPORE 2019-2020,” Ministry of Health. Accessed: Feb. 08, 2025. [Online]. Available: https://www.moh.gov.sg/others/resources-and-statistics/reports-communicable-diseases-surveillance-in-singapore-2019-2020/

[7] K. Sinka, “The global burden of sexually transmitted infections,” Clin Dermatol, vol. 42, no. 2, pp. 110–118, 2024, doi: 10.1016/j.clindermatol.2023.12.002.

[8] Y. Zheng et al., “Syphilis epidemic among men who have sex with men: A global systematic review and meta-analysis of prevalence, incidence, and associated factors,” J Glob Health, vol. 14, p. 04004, doi: 10.7189/jogh.14.04004.

[9] R. F. Baggaley et al., “Heterosexual Anal Intercourse: A Neglected Risk Factor for HIV?,” Am J Reprod Immunol, vol. 69, no. 0 1, pp. 95–105, Feb. 2013, doi: 10.1111/aji.12064.

[10] R. Parker and P. Aggleton, “HIV and AIDS-related stigma and discrimination: a conceptual framework and implications for action,” Social Science & Medicine, vol. 57, no. 1, pp. 13–24, July 2003, doi: 10.1016/S0277-9536(02)00304-0.

[11] L. H. Bachmann, “CDC Clinical Guidelines on the Use of Doxycycline Postexposure Prophylaxis for Bacterial Sexually Transmitted Infection Prevention, United States, 2024,” MMWR Recomm Rep, vol. 73, 2024, doi: 10.15585/mmwr.rr7302a1.

[12] D. E. Mauck et al., “Population-based methods for estimating the number of men who have sex with men: A systematic review,” Sex Health, vol. 16, no. 6, pp. 527–538, Nov. 2019, doi: 10.1071/SH18172.

[13] “Syphilis - Annual Epidemiological Report for 2019,” European Centre for Disease Prevention and Control. Accessed: July 18, 2025. [Online]. Available: https://www.ecdc.europa.eu/en/publications-data/syphilis-annual-epidemiological-report-2019

[14] J.-M. Molina et al., “Post-exposure prophylaxis with doxycycline to prevent sexually transmitted infections in men who have sex with men: an open-label randomised substudy of the ANRS IPERGAY trial,” Lancet Infect Dis, vol. 18, no. 3, pp. 308–317, Mar. 2018, doi: 10.1016/S1473-3099(17)30725-9.

[15] J.-M. Molina et al., “Doxycycline prophylaxis and meningococcal group B vaccine to prevent bacterial sexually transmitted infections in France (ANRS 174 DOXYVAC): a multicentre, open-label, randomised trial with a 2 × 2 factorial design,” The Lancet Infectious Diseases, vol. 24, no. 10, pp. 1093–1104, Oct. 2024, doi: 10.1016/S1473-3099(24)00236-6.

[16] J. Saunders, J. Deering, C. Dewsnap, R. Drayton, and J. Gilmore, “UK National Guideline for the Use of Doxycycline Post-Exposure Prophylaxis (DoxyPEP) for the Prevention of Syphilis”.

[17] J. C. Dombrowski et al., “Evidence-Informed Provision of Doxycycline Postexposure Prophylaxis for Prevention of Bacterial Sexually Transmitted Infections,” *Clinical Infectious Diseases*, p. ciae527, Oct. 2024, doi: 10.1093/cid/ciae527.

[18] J. S. Grant et al., “Doxycycline Prophylaxis for Bacterial Sexually Transmitted Infections,” Clinical Infectious Diseases, vol. 70, no. 6, pp. 1247–1253, Mar. 2020, doi: 10.1093/cid/ciz866.

[19] A. Hazra et al., “Filling in the Gaps: Updates on Doxycycline Prophylaxis for Bacterial Sexually Transmitted Infections,” *Clinical Infectious Diseases*, p. ciae062, Feb. 2024, doi: 10.1093/cid/ciae062.

[20] R. K. Bolan, M. R. Beymer, R. E. Weiss, R. P. Flynn, A. A. Leibowitz, and J. D. Klausner, “Doxycycline prophylaxis to reduce incident syphilis among HIV-infected men who have sex with men who continue to engage in high-risk sex: a randomized, controlled pilot study,” Sex Transm Dis, vol. 42, no. 2, pp. 98–103, Feb. 2015, doi: 10.1097/OLQ.0000000000000216.

[21] A. F. Luetkemeyer et al., “Postexposure Doxycycline to Prevent Bacterial Sexually Transmitted Infections,” N Engl J Med, vol. 388, no. 14, pp. 1296–1306, Apr. 2023, doi: 10.1056/NEJMoa2211934.

[22] J. Stewart et al., “Doxycycline Prophylaxis to Prevent Sexually Transmitted Infections in Women,” N Engl J Med, vol. 389, no. 25, pp. 2331–2340, Dec. 2023, doi: 10.1056/NEJMoa2304007.

[23] M. Sankaran et al., “Doxycycline Postexposure Prophylaxis and Sexually Transmitted Infection Trends,” JAMA Internal Medicine, Jan. 2025, doi: 10.1001/jamainternmed.2024.7178.

[24] M. W. Traeger et al., “Doxycycline Postexposure Prophylaxis and Bacterial Sexually Transmitted Infections Among Individuals Using HIV Preexposure Prophylaxis,” JAMA Internal Medicine, Jan. 2025, doi: 10.1001/jamainternmed.2024.7186.

[25] J. Zhu et al., “Impact of screening and doxycycline prevention on the syphilis epidemic among men who have sex with men in British Columbia: a mathematical modelling study,” The Lancet Regional Health - Americas, vol. 33, p. 100725, May 2024, doi: 10.1016/j.lana.2024.100725.

[26] M. W. Traeger, K. H. Mayer, D. S. Krakower, S. Gitin, S. M. Jenness, and J. L. Marcus, “Potential Impact of Doxycycline Post-exposure Prophylaxis Prescribing Strategies on Incidence of Bacterial Sexually Transmitted Infections,” *Clinical Infectious Diseases*, p. ciad488, Aug. 2023, doi: 10.1093/cid/ciad488.

[27] D. P. Wilson et al., “Chemoprophylaxis is likely to be acceptable and could mitigate syphilis epidemics among populations of gay men,” Sex Transm Dis, vol. 38, no. 7, pp. 573–579, July 2011, doi: 10.1097/OLQ.0b013e31820e64fd.

[28] M. Holt et al., “Acceptability of Doxycycline Prophylaxis, Prior Antibiotic Use, and Knowledge of Antimicrobial Resistance Among Australian Gay and Bisexual Men and Nonbinary People,” Sex Transm Dis, vol. 52, no. 2, pp. 73–80, Feb. 2025, doi: 10.1097/OLQ.0000000000002079.

[29] L. Geng et al., “Potential public health impacts of gonorrhea vaccination programmes under declining incidences: A modeling study,” PLoS Med, vol. 22, no. 2, p. e1004521, Feb. 2025, doi: 10.1371/journal.pmed.1004521.

[30] S. E. D. Quaye et al., “Application of the network scale-up method to estimate the sizes of key populations for HIV in Singapore using online surveys,” J Int AIDS Soc, vol. 26, no. 3, p. e25973, Mar. 2023, doi: 10.1002/jia2.25973.

[31] L. K. Whittles, X. Didelot, and P. J. White, “Public health impact and cost-effectiveness of gonorrhoea vaccination: an integrated transmission-dynamic health-economic modelling analysis,” The Lancet Infectious Diseases, vol. 22, no. 7, pp. 1030–1041, July 2022, doi: 10.1016/S1473-3099(21)00744-1.

[32] “Taking doxycycline with other medicines and herbal supplements,” National Health Service. Accessed: Feb. 10, 2025. [Online]. Available: https://www.nhs.uk/medicines/doxycycline/taking-doxycycline-with-other-medicines-and-herbal-supplements/

[33] “Who can and cannot take doxycycline,” National Health Service. Accessed: Feb. 10, 2025. [Online]. Available: https://www.nhs.uk/medicines/doxycycline/who-can-and-cannot-take-doxycycline/

[34] M. H. Junejo, J. L. Marcus, and K. A. Katz, “Doxycycline Postexposure Prophylaxis (DoxyPEP) for Bacterial STI Prevention,” JAMA Dermatology, vol. 161, no. 1, pp. 7–8, Jan. 2025, doi: 10.1001/jamadermatol.2024.4567.

[35] “About doxycycline,” National Health Service. Accessed: Feb. 20, 2025. [Online]. Available: https://www.nhs.uk/medicines/doxycycline/about-doxycycline/

[36] N. Borel, C. Leonard, J. Slade, and R. V. Schoborg, “Chlamydial Antibiotic Resistance and Treatment Failure in Veterinary and Human Medicine,” Curr Clin Microbiol Rep, vol. 3, pp. 10–18, 2016, doi: 10.1007/s40588-016-0028-4.

[37] A. Sanchez et al., “Surveillance of Antibiotic Resistance Genes in Treponema Pallidum Subspecies Pallidum from Patients with Early Syphilis in France,” Acta Derm Venereol, vol. 100, no. 14, p. adv00221, July 2020, doi: 10.2340/00015555-3589.

[38] J. Zhang et al., “Discontinuation, suboptimal adherence, and reinitiation of oral HIV pre-exposure prophylaxis: a global systematic review and meta-analysis,” The Lancet HIV, vol. 9, no. 4, pp. e254–e268, Apr. 2022, doi: 10.1016/S2352-3018(22)00030-3.

[39] “STI Screening Recommendations,” Centers for Disease Control and Prevention. Accessed: June 08, 2025. [Online]. Available: https://www.cdc.gov/std/treatment-guidelines/screening-recommendations.htm

[40] L. W. Ang, C. S. Wong, O. T. Ng, and Y. S. Leo, “Incidence of syphilis among HIV-infected men in Singapore, 2006-2017: temporal trends and associated risk factors,” Sex Transm Infect, vol. 96, no. 4, pp. 293–299, June 2020, doi: 10.1136/sextrans-2019-054163.

[41] H. S. Marshall et al., “Management and prevention of Neisseria meningitidis and Neisseria gonorrhoeae infections in the context of evolving antimicrobial resistance trends,” Eur J Clin Microbiol Infect Dis, vol. 44, no. 2, pp. 233–250, Feb. 2025, doi: 10.1007/s10096-024-04968-8.

[42] M. W. Traeger, J. E. Volk, and J. L. Marcus, “Doxycycline Postexposure Prophylaxis Could Accelerate the Emergence of Gonococcal Ceftriaxone Resistance—Reply,” JAMA Internal Medicine, June 2025, doi: 10.1001/jamainternmed.2025.0987.

[43] “STI Management Guidelines 7th Edition | DSC Clinic - Department of STI Control Singapore,” National Skin Centre. Accessed: July 19, 2025. [Online]. Available: https://www.nsc.com.sg/dsc/healthcare-professionals/publications/Pages/STI-Management-Guidelines.aspx

[44] “Sexually transmitted infections and screening for chlamydia in England: 2024 report,” GOV.UK. Accessed: July 19, 2025. [Online]. Available: https://www.gov.uk/government/statistics/sexually-transmitted-infections-stis-annual-data-tables/sexually-transmitted-infections-and-screening-for-chlamydia-in-england-2024-report

[45] D. Nikitin, L. K. Whittles, J. W. Imai-Eaton, and P. J. White, “Cost-effectiveness of 4CMenB Vaccination Against Gonorrhea: Importance of Dosing Schedule, Vaccine Sentiment, Targeting Strategy, and Duration of Protection,” J Infect Dis, vol. 231, no. 1, pp. 71–83, Apr. 2024, doi: 10.1093/infdis/jiae123.

[46] L. K. Whittles, P. J. White, and X. Didelot, “A dynamic power-law sexual network model of gonorrhoea outbreaks,” PLoS Comput Biol, vol. 15, no. 3, p. e1006748, Mar. 2019, doi: 10.1371/journal.pcbi.1006748.

