## Supplementary Information for "Long-Run Public Health Impact of Doxycycline Post-Exposure Prophylaxis and Behavioural Factors on Syphilis Transmission: A Modelling Study in Singapore and England"

**Appendix A: Background of Syphilis and Doxy-PEP**

1. Stages of syphilis

Syphilis is a chronic, systemic STI which progresses through four distinct stages - primary, secondary, latent, and tertiary - each with characteristic clinical features and varying degrees of infectivity. The primary stage typically presents as a solitary, painless ulcer (chancre) at the site of inoculation, which resolves spontaneously within a few weeks. If untreated, the infection advances to the secondary stage, which is characterized by systemic manifestations such as a generalized maculopapular rash (often involving the palms and soles), flu-like symptoms, and mucous membrane lesions. These symptoms may also resolve without treatment, leading to the latent stage. The latent stage of syphilis is asymptomatic and is divided into early latent (within one year of infection) and late latent (beyond one year). Individuals with early latent syphilis remain infectious, while those with late latent disease are generally considered non-infectious. Without treatment, syphilis can progress to the tertiary stage, which may occur years or even decades after initial infection. Tertiary syphilis can result in serious complications, including gummatous lesions, cardiovascular syphilis, and neurosyphilis, potentially leading to multi-organ damage and death [1].

2. Diagnosis of syphilis

The diagnosis of syphilis relies primarily on serologic testing, as *T. pallidum* cannot be cultured using standard laboratory methods. A two-step testing algorithm is commonly employed; in Singapore, non-treponemal and treponemal-specific tests are typically performed concurrently. Initial screening is conducted using non-treponemal tests[1] such as the Rapid Plasma Reagin (RPR) or Venereal Disease Research Laboratory (VDRL) test, which detect antibodies correlated with disease activity. Positive results are confirmed using treponemal-specific tests[2], including the Treponema pallidum Particle Agglutination (TPPA) assay, Fluorescent Treponemal Antibody Absorption (FTA-ABS), Enzyme Immunoassay (EIA), or Chemiluminescent Immunoassay (CIA). Treponemal antibodies generally persist for life due to a phenomenon known as the “serological scar.” As a result, non-treponemal test titers are used to assess disease activity. High titers (e.g., RPR ≥ 1:8) indicate active infection, while low titers (e.g., ≤ 1:4) may reflect past or treated infections. In cases with discordant results, clinical history, risk factors, and follow-up testing are necessary to guide management.

3. Treatment of syphilis

Penicillin remains the gold standard for syphilis treatment across all disease stages. Primary, secondary, and early latent syphilis are treated with a single intramuscular injection of Benzathine Penicillin G (2.4 million units). Late latent syphilis and syphilis of unknown duration require three weekly doses of Benzathine Penicillin G (2.4 million units IM). Tertiary syphilis, including gummatous and cardiovascular manifestations, is managed with the same three-dose regimen. Neurosyphilis and ocular syphilis necessitate intravenous Aqueous Penicillin G (18 - 24 million units per day), administered either every four hours or as a continuous infusion for 10–14 days. For patients allergic to penicillin, doxycycline or ceftriaxone may be considered, although penicillin desensitization is preferred, particularly in cases of neurosyphilis or pregnancy.

4. Doxy-PEP

Doxycycline is a second-generation tetracycline with moderate-spectrum activity that is well tolerated and rapidly absorbed following oral administration [2], which has been extensively used for treating acne, preventing malaria,

---

[1] Nontreponemal tests (screening tests) detect reagin antibodies produced in response to syphilis but can also be positive in other conditions.

[2] Treponemal tests (confirmation tests) detect antibodies specific to *Treponema pallidum* and remain positive for life.

and managing rosacea [3]. Doxy-PEP involves taking a 200 mg dose of immediate-release doxycycline within 72 hours after condomless sex to reduce the risk of bacterial STIs, including syphilis, as recommended by British Association for Sexual Health and HIV (BASHH) [4] and Centre for Disease Control and Prevention (CDC). A 30- to 90-day supply is typically recommended and clinicians should conduct screenings for HIV and syphilis at least every three months for individuals using doxy-PEP [5]. Additionally, CDC advises that MSM and TGW diagnosed with a bacterial STI (e.g., syphilis) within the past 12 months should be informed about doxy-PEP as a post-exposure prophylaxis option to reduce the risk of reinfection [3].

5. Acceptability of doxy-PEP

International studies have examined the acceptability and usage of doxy-PEP among MSM. In Australia, a survey found that 75.8% of participants viewed doxy-PEP as an acceptable STI prevention strategy, with 7.5% reporting prior antibiotic use for this purpose [6]. By comparison, condoms were considered acceptable by 45.1% of respondents, while STI PrEP was rated acceptable by 54.0% [6]. Similarly, a US study revealed that 84% of participants were interested in trying doxy-PEP, and 13% had used it within the past year [7], [8].

**Appendix B: Syphilis Transmission Model**

Table 1. Annual syphilis diagnoses among MSM in Singapore under different scenarios: (1) Main scenario: male syphilis incidence minus female incidence, assuming all males not having sex with females are MSM; (2) Upper bound: male syphilis incidence assuming all males are MSM; and (3) Lower bound: male syphilis incidence multiplied by the proportion of MSM in the male population, assuming equal risk of syphilis infection regardless of sexual preference.

| Year | Main scenario | Upper bound | Lower bound |
|---|---|---|---|
| 2004 | 324 | 518 | 31 |
| 2005 | 365 | 632 | 38 |
| 2006 | 488 | 840 | 51 |
| 2007 | 346 | 564 | 34 |
| 2008 | 305 | 599 | 36 |
| 2009 | 197 | 532 | 32 |
| 2010 | 159 | 407 | 24 |
| 2011 | 187 | 536 | 32 |
| 2012 | 318 | 875 | 53 |
| 2013 | 308 | 801 | 48 |
| 2014 | 525 | 935 | 56 |
| 2015 | 496 | 832 | 50 |
| 2016 | 623 | 894 | 54 |
| 2017 | 573 | 804 | 49 |
| 2018 | 605 | 863 | 52 |

Table 2. Annual syphilis diagnoses among MSM in England.

| Year | Reported cases |
|---|---|
| 2011 | 2030 |
| 2012 | 2123 |
| 2013 | 2379 |
| 2014 | 3485 |
| 2015 | 4052 |
| 2016 | 4605 |
| 2017 | 5440 |
| 2018 | 5610 |
| 2019 | 5761 |
| 2022 | 5969 |
| 2023 | 6313 |
| 2024 | 6169 |

Table 3: Fixed model parameters (Singapore): notation, definitions, source of estimates. For the survey conducted in Singapore, responses were collected using multiple recruitment strategies to ensure representation across diverse demographic groups. To reach younger participants, the survey was disseminated through popular social media platforms such as Facebook and Telegram. To increase participation among individuals with diverse sexual orientations, the survey was promoted in collaboration with Pink Dot SG - a non-profit movement supporting the LGBTQ community - via their Instagram account and at the offline Pink Dot 16 event. To engage older residents, additional responses were gathered through the survey company IPSOS. Assuming all individuals participate in anal sex also participate in oral sex (i.e., anal sex population is a subset of oral sex population) [9]. Transition rate parameters ($\theta \in \{\psi_T, \nu\}$) are presented in an annual basis, giving a mean time to transition of $365/\theta$ days.

| | Definition | Value | Source |
|---|---|---|---|
| $N(t0)$ | Initial population size of Singapore MSM (age 15-65) | 97,189 | Derived from [10][3] |
| $\alpha$ | Annual MSM population entrants (at age 15) | 2,524 | Derived from [10][4] |
| $\gamma$ | Years spent in the sexually active population | 50 | Ages 15-65 |
| $q_L$ | Proportion of the MSM population in group $L$ | 0.793 | Calibrated from survey |
| $q_H$ | Proportion of the MSM population in group $H$ | 0.207 | $1 - q_L$ |
| $c_L$ | Annual rate of partner change in group $L$ | 1.989 | Calibrated from survey |
| $c_H$ | Annual rate of partner change in group $H$ | 14.866 | Calibrated from survey |
| $\psi_T$ | Probability of leaving late-latent stage ($L \rightarrow T$) | 0.05 | Derived from [11], [12][5] |
| $\nu$ | Mortality rate at tertiary stage | 0.00967 | Derived from [13], [14][6] |

[3] Applied the 2019 estimate of the percentage of MSM in the male population [10] to the male population data from 2004.

[4] Applied the 2019 estimate of the percentage of MSM in the male population [10] to the male population entrants (age 15) from 2004 to 2018 and calculated the average.

[5] Assumed a progression period of 20 years from the late latent stage to the tertiary stage.

[6] Applied an exponential hazard rate of $-\ln(1-p)/t$ over 40 years, corresponding to an approximately 30% probability of death in the tertiary stage without treatment.

Table 4: Fixed model parameters (England): notation, definitions, source of estimates. Assuming all individuals participate in anal sex also participate in oral sex (i.e., anal sex population is a subset of oral sex population) [9]. Transition rate parameters ($\theta \in \{\psi_T, \nu\}$) are presented in an annual basis, giving a mean time to transition of $365/\theta$ days.

| | Definition | Value | Source |
|---|---|---|---|
| $N(t0)$ | Initial population size of England MSM (age 15-65) | 600,000 | [15] |
| $\alpha$ | Annual MSM population entrants (at age 15) | 12,000 | [15] |
| $\gamma$ | Years spent in the sexually active population | 50 | Ages 15-65 |
| $q_L$ | Proportion of the MSM population in group *L* | 0.85 | [15] |
| $q_H$ | Proportion of the MSM population in group *H* | 0.15 | $1 - q_L$ |
| $c_L$ | Annual rate of partner change in group *L* | 0.6 | [15] |
| $c_H$ | Annual rate of partner change in group *H* | 15.6 | [15] |
| $\psi_T$ | Probability of leaving late-latent stage ($L \rightarrow T$) | 0.05 | Derived from [11], [12][7] |
| $\nu$ | Mortality rate at tertiary stage | 0.00967 | Derived from [13], [14][8] |

[7] Assumed a progression period of 20 years from the late latent stage to the tertiary stage.

[8] Applied an exponential hazard rate of $-\ln(1-p)/t$ over 40 years, corresponding to an approximately 30% probability of death in the tertiary stage without treatment.

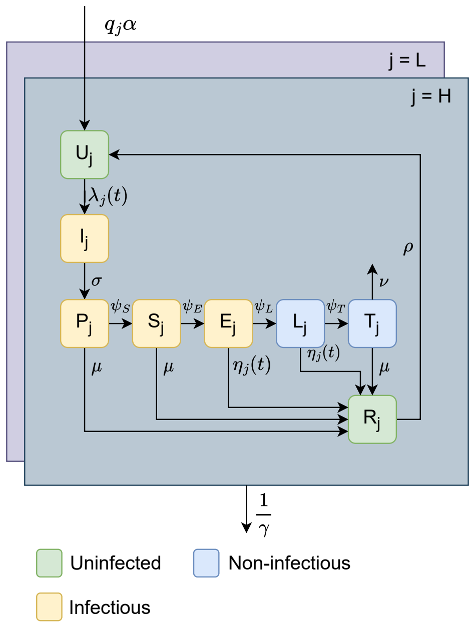


Figure 1: Model architecture for syphilis transmission. Individuals enter the sexually active population as uninfected ($U_j$). Upon infection, they progress through an incubation phase ($I_j$) before advancing to the primary ($P_j$), secondary ($S_j$), early latent ($E_j$), late latent ($L_j$), and tertiary ($T_j$) stages. The primary, secondary, and tertiary stages are symptomatic, prompting individuals to seek treatment and transition to the recovered state ($R_j$). In contrast, early latent and late latent stages are asymptomatic, with infections typically detected through screening, leading to treatment and recovery ($R_j$). Individuals in the tertiary stage face a risk of mortality, permanently exiting the compartments. Notably, only the incubation, primary, secondary, and early latent stages are infectious, while the late latent and tertiary stages are non-infectious. All treated infections are considered cured, with individuals returning to the uninfected state. Individuals may exit the sexually active population due to aging at any stage. The model includes separate compartments for individuals with low and high sexual activity levels ($j \in L, H$), represented by the purple and grey layers in the figure. Both groups share an identical compartmental structure, though, for clarity, only the transitions in and out of the high-activity group (upper layer) are depicted.

Model of syphilis transmission:

$$\frac{dU_j(t)}{dt} = q_j\alpha - \left(\lambda_j(t) + \frac{1}{\gamma}\right)U_j(t) + \rho R_j(t) \tag{1}$$

$$\frac{dI_j(t)}{dt} = \lambda_j(t)U_j(t) - \left(\sigma + \frac{1}{\gamma}\right)I_j(t) \tag{2}$$

$$\frac{dP_j(t)}{dt} = \sigma I_j(t) - \left(\mu + \psi_S + \frac{1}{\gamma}\right)P_j(t) \tag{3}$$

$$\frac{dS_j(t)}{dt} = \psi_S P_j(t) - \left(\mu + \psi_E + \frac{1}{\gamma}\right)S_j(t) \tag{4}$$

$$\frac{dE_j(t)}{dt} = \psi_E S_j(t) - \left(\eta_j(t) + \psi_L + \frac{1}{\gamma}\right)E_j(t) \tag{5}$$

$$\frac{dL_j(t)}{dt} = \psi_L E_j(t) - \left(\eta_j(t) + \psi_T + \frac{1}{\gamma}\right)L_j(t) \tag{6}$$

$$\frac{dT_j(t)}{dt} = \psi_T L_j(t) - \left(\mu + \nu + \frac{1}{\gamma}\right)L_j(t) \tag{7}$$

$$\frac{dR_j(t)}{dt} = \mu\left(P_j(t) + S_j(t) + T_j(t)\right) + \eta_j(t)\left(E_j(t) + L_j(t)\right) - \left(\rho + \frac{1}{\gamma}\right)R_j(t) \tag{8}$$

For shared variables:

$$\lambda_j(t) = c_j\beta\left(1 + \phi_\beta(t - t_0)\right)\left(\epsilon\frac{C_j(t)}{N_j(t)} + (1-\epsilon)\left(\sum_{i\in\{L,H\}} \pi_i(t)\frac{C_i(t)}{N_i(t)}\right)\right) \tag{9}$$

$$\pi_j(t) = \frac{c_j N_j(t)}{\sum_{i\in\{L,H\}} c_i\, N_i(t)} \tag{10}$$

$$C_j(t) = \sum_{i\in\{N,X,D,M\}} \left(I_j^i(t) + P_j^i(t) + S_j^i(t) + E_j^i(t)\right) \tag{11}$$

$$N_j(t) = \sum_{i\in\{N,X,D,M\}} \left(U_j^i(t) + I_j^i(t) + P_j^i(t) + S_j^i(t) + E_j^i(t) + L_j^i(t) + T_j^i(t) + R_j^i(t)\right) \tag{12}$$

$$\eta_H(t) = \eta_H(t_0)\left(1 + \phi_\eta(t - t_0)\right) \tag{13}$$

$$\eta_L(t) = \omega\eta_H(t) \tag{14}$$

We compared the observed annual number of syphilis diagnoses $Z_D(t)$ with those predicted by our model $Y_D(t)$ using Negative-Binomial likelihoods to allow for over-dispersion in the observation process relative to a Poisson distribution. If $x \sim$ NegBinom($m$, $\kappa$), with mean $m$ and shape parameter $\kappa$, then:

$$f_X(x|m,\kappa) = \frac{\Gamma(\kappa + x)}{x!\,\Gamma(\kappa)}\left(\frac{\kappa}{\kappa + m}\right)^{\kappa}\left(\frac{m}{\kappa + m}\right)^{x} \tag{9}$$

where the shape parameter characterises the level of clustering or heterogeneity in the observation process, and $\Gamma(\cdot)$ is the Gamma function. Under the negative binomial distribution $Var(x) = m + m^2/k$.

The likelihoods of $Z_D(t)$ was:

$$Z_D(t) \sim Negbinom(Y_D(t), \kappa_D) \tag{10}$$

where

$$Y_D(t) = \sum_{j\in\{L,H\}} \left(\int_t^{t+1} \rho R_j(\tau)d\tau\right) \tag{11}$$

The likelihood of the observation given the modelled trajectories produced by parameter set Θ was calculated as the product of the likelihoods of the data stream in each year t = 2004 ($t_0$), …, 2018 ($t_{max}$) for Singapore and t = 2011 ($t_0$), …, 2020, 2023, 2024 ($t_{max}$) for England.

$$L(Z|\Theta) = \prod_{t=t_0}^{t_{max}} f_D(Z_D(t)|\Theta) \quad (12)$$

Table 5. The effective sample size (ESS) and Gelman-Rubin (GR) diagnostic of posterior parameter estimates under different observation scenarios.

| | Singapore (main scenario) | | Singapore (upper bound) | | Singapore (lower bound) | | England | |
|---|---|---|---|---|---|---|---|---|
| | ESS | GR | ESS | GR | ESS | GR | ESS | GR |
| $\beta$ | 3,797 | 1.00 | 3,505 | 1.00 | 3,074 | 1.00 | 2,825 | 1.00 |
| $\phi_\beta$ | 3,881 | 1.00 | 5,056 | 1.00 | 5,562 | 1.00 | 5,543 | 1.00 |
| $\epsilon$ | 7,580 | 1.00 | 6,572 | 1.00 | 5,690 | 1.00 | 7,555 | 1.00 |
| $\mu$ | 5,159 | 1.00 | 5,202 | 1.00 | 5,751 | 1.00 | 5492 | 1.00 |
| $\rho$ | 6,922 | 1.00 | 5,908 | 1.00 | 6,726 | 1.00 | 6,113 | 1.00 |
| $\eta_H(t_0)$ | 7,886 | 1.00 | 6,165 | 1.00 | 7,799 | 1.00 | 7,726 | 1.00 |
| $\phi_\eta$ | 8,265 | 1.00 | 5,930 | 1.00 | 5,992 | 1.00 | 6,913 | 1.00 |
| $\omega$ | 4,317 | 1.00 | 4,133 | 1.00 | 5,141 | 1.00 | 5,073 | 1.00 |
| $\sigma$ | 3,975 | 1.00 | 3,384 | 1.00 | 2,947 | 1.00 | 2,633 | 1.00 |
| $\psi_S$ | 4,535 | 1.00 | 3,807 | 1.00 | 5,698 | 1.00 | 4,316 | 1.00 |
| $\psi_E$ | 5,635 | 1.00 | 4,504 | 1.00 | 4,160 | 1.00 | 4,760 | 1.00 |
| $\psi_L$ | 5,599 | 1.00 | 3,539 | 1.00 | 5,101 | 1.00 | 3,099 | 1.00 |
| $\kappa_D$ | 7,137 | 1.00 | 7,810 | 1.00 | 8,575 | 1.00 | 7,026 | 1.00 |

(A) Singapore (main scenario)

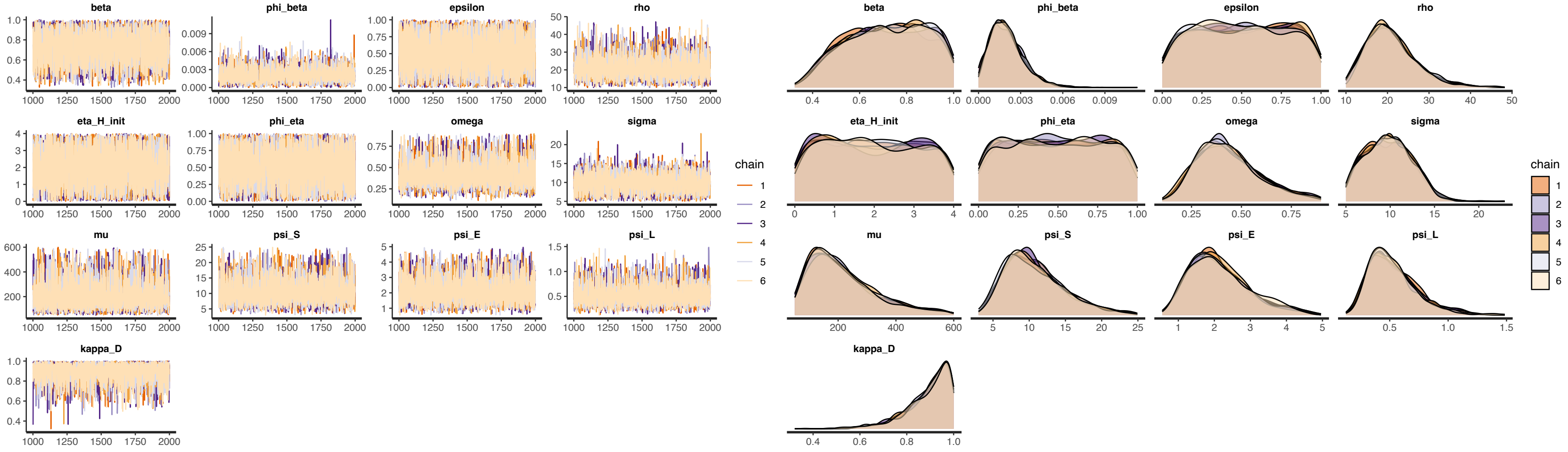


(B) Singapore (upper bound)

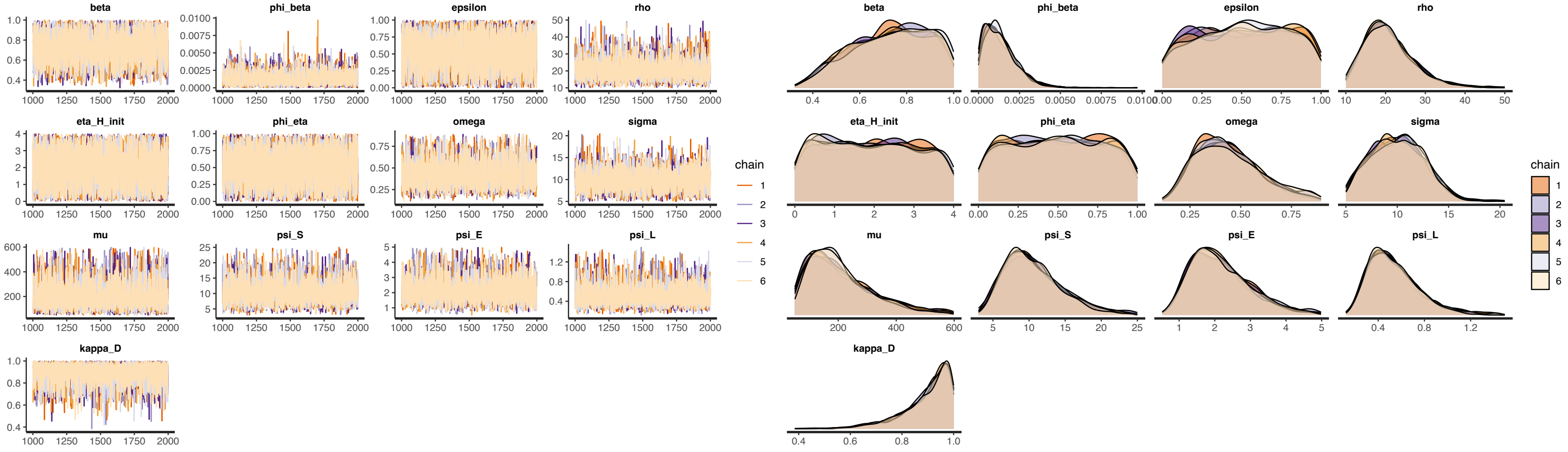

(C) Singapore (lower bound)

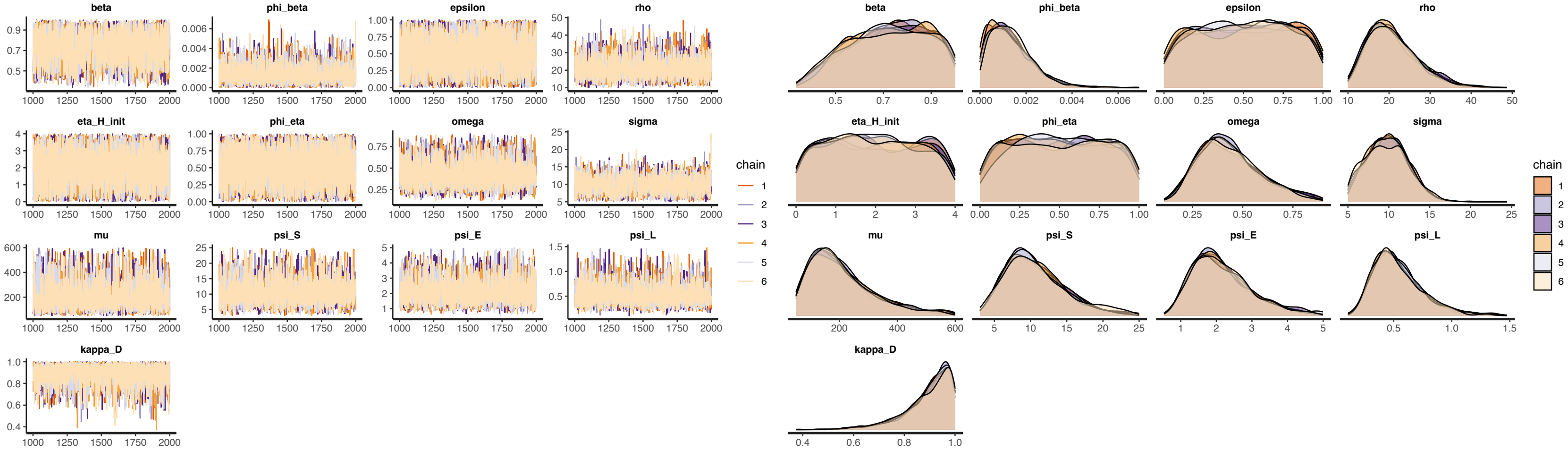


(D) England

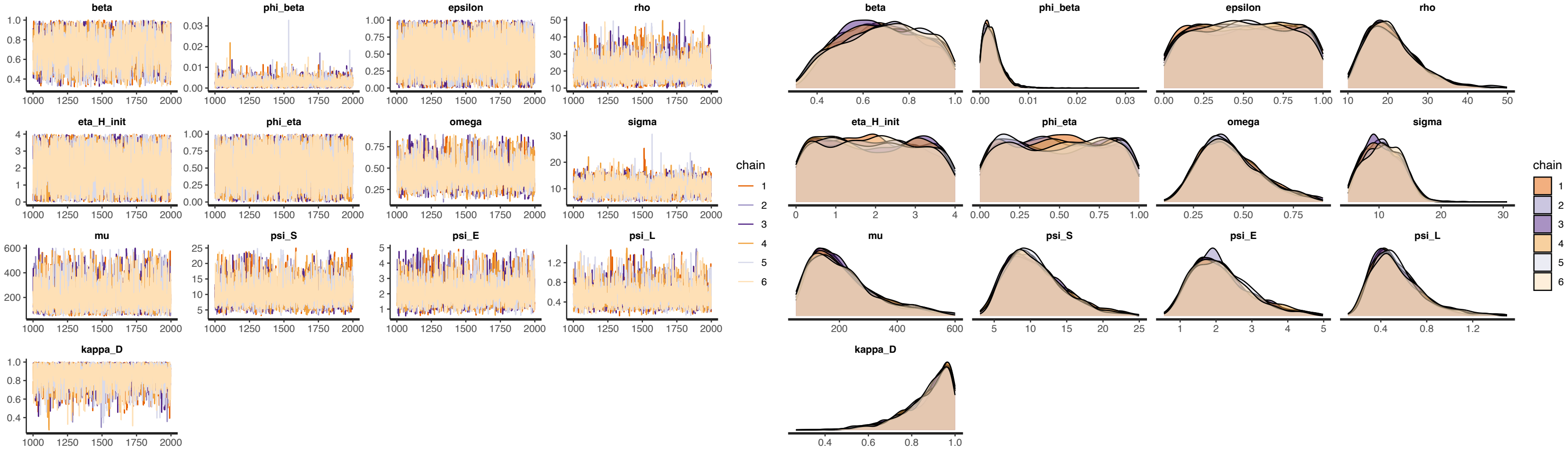

Fig. 2: Traceplots (left column) and marginal posterior densities (right column) of parameter estimates under different observation scenarios. Traceplots show MCMC samples from six chains, each with 2,000 iterations, with the first 1,000 discarded as burn-in. Coloured lines represent individual chains. For Singapore, the model was fitted to three scenarios: (A) male syphilis incidence minus female incidence, assuming all men not having sex with women are MSM; (B) total male syphilis incidence, assuming all men are MSM; and (C) male syphilis incidence scaled by the proportion of MSM in the male population, assuming equal infection risk across all men. For England, (D) the model was fitted to reported syphilis cases among MSM.

Table 6: Fitted parameters: notation, definitions, prior distributions and posterior estimates. Transition rate parameters ($\theta \in \{\sigma, \psi_S, \psi_E, \psi_L, \mu, \rho\}$) are presented in an annual basis, giving a mean time to transition of $365/\theta$ days. Posterior estimates are taken as the median, with 95% credible interval presented in parentheses.

| | Definition | Prior distribution | Parameter bound | Posterior estimate | | | |
|---|---|---|---|---|---|---|---|
| | | | | Singapore (main scenario) | Singapore (upper bound) | Singapore (lower bound) | England |
| $\beta$ | Probability of transmission per partnership | U[0, 1] | [0, 1] | 0.722 (0.406, 0.983) | 0.747 (0.413, 0.987) | 0.735 (0.416, 0.986) | 0.687 (0.375, 0.979) |
| $\phi_\beta$ | Annual increase in transmission risk behaviour | U[0, 1] | [0, 1] | 0.00185 (0.00017, 0.0048) | 0.00115 (0.00005, 0.00367) | 0.00115 (0.00007, 0.,00359) | 0.00221 (0.00012, 0.00713) |
| $\epsilon$ | Level of assortativity in sexual mixing | U[0, 1] | [0, 1] | 0.508 (0.0321, 0.976) | 0.515 (0.0248, 0.976) | 0.514 (0.0248, 0.975) | 0.510 (0.0263, 0.980) |
| $\mu$ | Rate of seeking treatment due to symptoms (*P/S/T* → *R*) | logN[5.298, 0.6] | [50, 600] | 187.716 (63.916, 512.693) | 181.411 (61.813, 510.785) | 184.490 (62.694, 505.406) | 186.659 (62.316, 505.317) |
| $\rho$ | Rate of recovery after treatment (*R* → *U*) | logN[2.996, 0.3] | [10, 50] | 20.076 (11.632, 35.735) | 19.918 (11.470, 35.934) | 19.949 (11.734, 34.937) | 19.907 (11.267, 36.754) |
| $\eta_H(t_0)$ | Initial rate of asymptomatic screening in group *H* | U[0, 4] | [0, 4] | 1.909 (0.0786, 3.886) | 1.937 (0.0842, 3.893) | 1.932 (0.0945, 3.887) | 1.931 (0.0670, 3.891) |
| $\phi_\eta$ | Annual increase in asymptomatic screening rate | U[0, 1] | [0, 1] | 0.490 (0.0207, 0.976) | 0.497 (0.0257, 0.971) | 0.498 (0.0290, 0.972) | 0.497 (0.0247, 0.975) |
| $\omega$ | Ratio of asymptomatic screening rate in group *L* vs group *H* | logN[-0.87, 0.39] | [0.1, 0.9] | 0.412 (0.190, 0.800) | 0.417 (0.192, 0.803) | 0.414 (0.193, 0.799) | 0.412 (0.195, 0.791) |
| $\sigma$ | Rate of leaving incubation period (*I* → *P*) | logN[2.708, 0.5] | [5, 40] | 9.741 (5.438, 14.806) | 9.981 (5.489, 14.953) | 9.790 (5.506, 14.962) | 10.481 (5.503, 16.360) |
| $\psi_S$ | Rate of leaving primary stage (*P* → *S*) | logN[2.303, 0.4] | [3, 25] | 10.107 (4.669, 20.809) | 9.910 (4.603, 20.424) | 10.026 (4.434, 20.874) | 10.020 (4.725, 20.481) |
| $\psi_E$ | Rate of leaving secondary stage (*S* → *E*) | logN[0.693, 0.4] | [0.5, 5] | 2.005 (0.905, 4.136) | 2.032 (0.944, 4.195) | 2.009 (0.949, 4.220) | 2.020 (0.930, 4.219) |
| $\psi_L$ | Rate of leaving early-latent stage (*E* → *L*) | logN[-0.693, 0.4] | [0.1, 1.5] | 0.490 (0.229, 1.050) | 0.499 (0.221, 1.105) | 0.497 (0.228, 1.068) | 0.495 (0.227, 1.110) |
| $\kappa_D$ | Shape parameter of communicable disease surveillance data | U[0, 1] | [0, 1] | 0.918 (0.649, 0.997) | 0.925 (0.654, 0.997) | 0.920 (0.649, 0.997) | 0.906 (0.582, 0.997) |

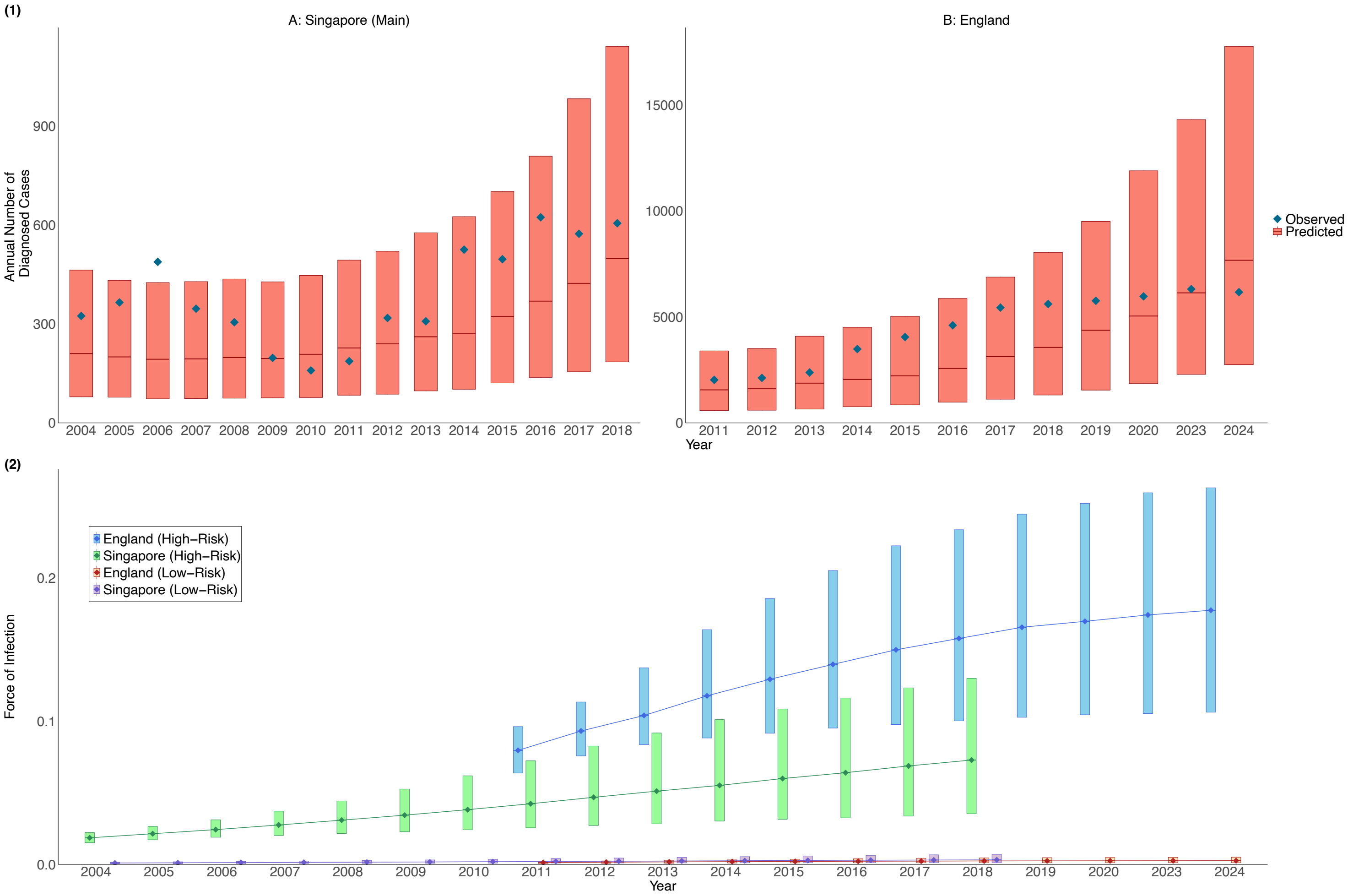


Fig. 3: Panel 1 shows simulated epidemic trajectories and observed syphilis incidence among MSM in England and Singapore (assuming all males not having sex with females are MSM). Each box represents the posterior distribution, with the median indicated by a horizontal line and the interquartile range represented by the upper and lower edges of the box. Observed annual syphilis incidence data are shown as diamonds. Panel 2 shows force of infection estimates based on Bayesian calibration for England and Singapore, stratified by high-risk and low-risk groups. For each box, the median is represented by a diamond, while the interquartile range is indicated by the upper and lower edges of the box.

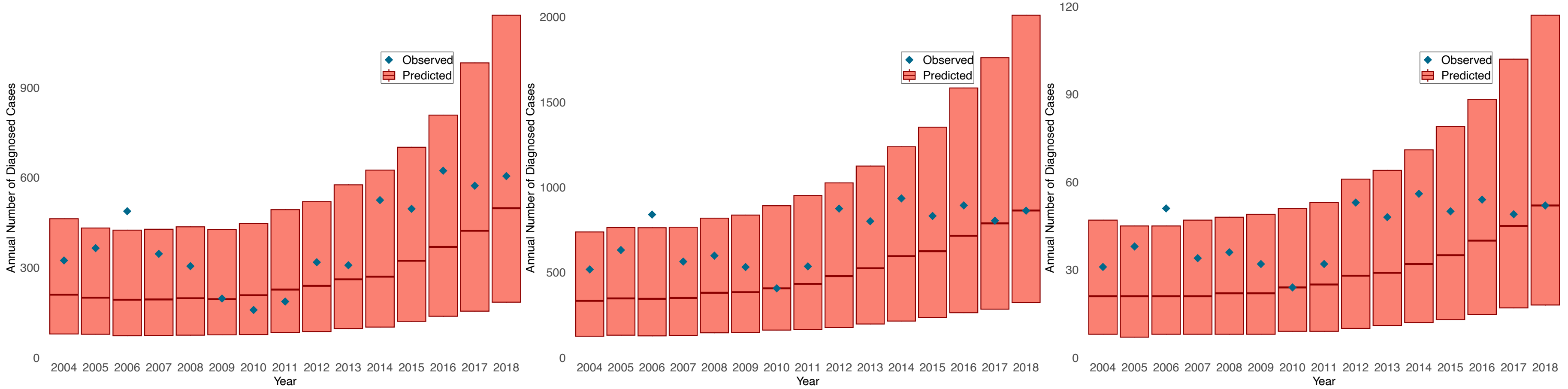


Fig. 4: Comparison of simulated epidemic trajectories and observed data in Singapore MSM under different scenarios. The median of each box is marked as a line, and the interquartile range is indicated by the upper and lower boundaries of the box respectively. Annual syphilis incidence of Singapore MSM under different observation scenarios are in squares. All three observation scenarios were created based on communicable disease surveillance Singapore data. From left to right, model fitted to three observation scenarios: (1) male syphilis incidence minus female incidence, assuming all males not having sex with females are MSM; (2) male syphilis incidence assuming all males are MSM; and (3) male syphilis incidence multiplied by the proportion of MSM in the male population, assuming equal risk of syphilis infection regardless of sexual preference.

**Appendix C: Doxy-PEP Model for Syphilis**

Simulations were conducted over time for MSM populations in Singapore and England, based on a sample of 1,000 parameter sets drawn from the joint posterior distribution. Two alternative future scenarios were considered: (A) a stabilized behaviour scenario, in which the inferred trends in time-varying behavioural parameters remain constant; and (B) a continued trend scenario, where these behavioural trends persist through to 2040.

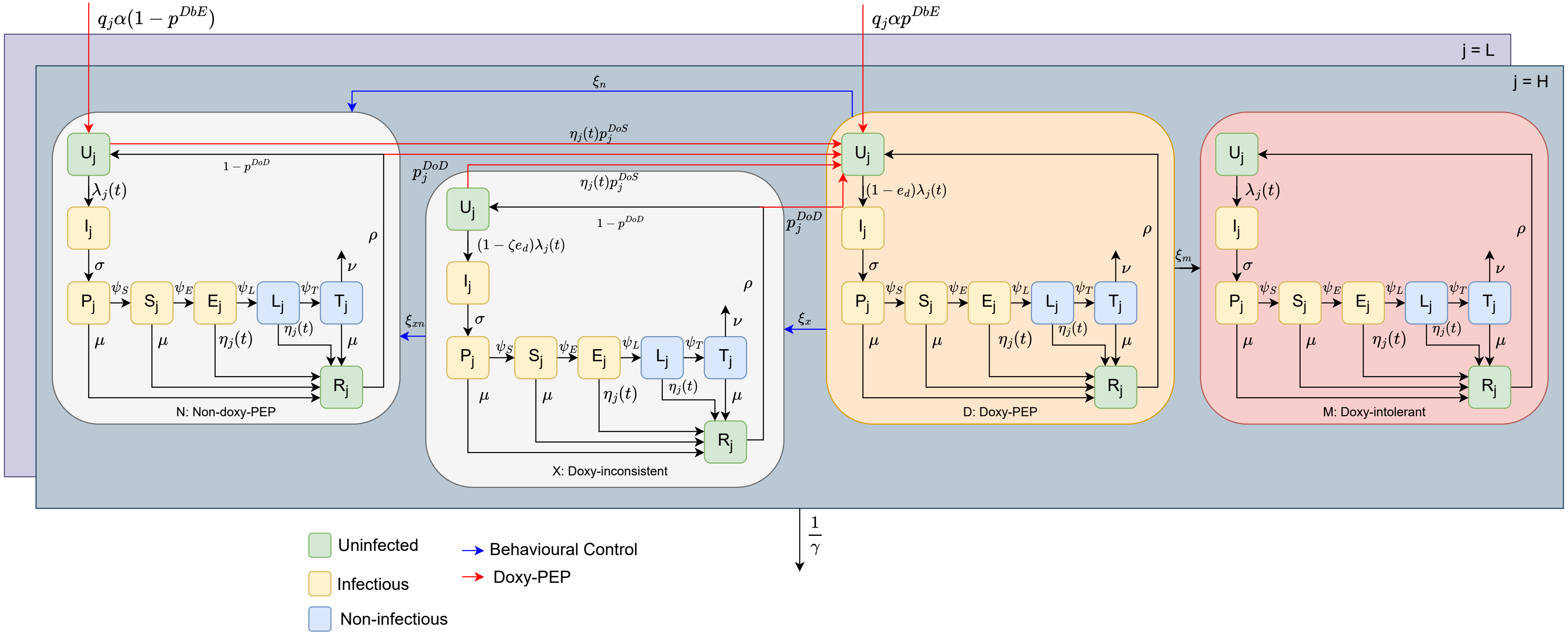


Fig. 5: Model architecture of doxy-PEP for syphilis. For each doxy-PEP stratum $i \in \{N, X, D, M\}$, the population is subdivided into compartments representing different stages of syphilis infection. The model distinguishes between individuals with low and high sexual activity levels ($j \in \{L, H\}$), illustrated as the grey (low-activity) and purple (high-activity) layers. Both groups follow the same compartmental structure but may differ in transition rates. For clarity, only the transitions in and out of the high-activity group (upper layer) are shown. For syphilis transmission (refer to stratum $N$), individuals enter the sexually active population as uninfected ($U_j$). Upon infection, driven by the time-varying force of infection $\lambda_j(t)$, they progress through an incubation phase ($I_j$) before advancing to the primary ($P_j$) at a rate $\sigma$, secondary ($S_j$) at a rate $\psi_S$, early latent ($E_j$) at a rate $\psi_E$, late latent ($L_j$) at a rate $\psi_L$, and tertiary ($T_j$) stages at a rate $\psi_T$. The primary, secondary, and tertiary stages are symptomatic, prompting individuals to seek treatment and transition to the recovered state ($R_j$) at a rate $\mu$. In contrast, early latent and late latent stages are asymptomatic, with infections typically detected through screening, leading to treatment ($R_j$) at a screening-dependent rate $\eta_j(t)$. Individuals in the tertiary stage face a risk of mortality, permanently exiting the model at a rate $\nu$. Notably, only the incubation, primary, secondary, and early latent stages are infectious, while the late latent and tertiary stages are non-infectious. Treated individuals are assumed to be cured and return to the uninfected compartment at a rate $\rho$. Individuals may exit the sexually active population due to aging at any stage at a rate $1/\gamma$. For the doxy-PEP model, individuals $\alpha$ enter the sexually active population at age 15. With probability $p^{DbE}$, they initiate doxy-PEP upon entry and enter stratum $D$; otherwise, they enter stratum $N$.

Individuals at stratum $D$ may discontinue doxy-PEP and move to stratum $N$ at a rate $\xi_n$; become suboptimally adherent and transition to stratum $X$ at a rate $\xi_x$; or develop intolerance and transition to stratum $M$ at a rate $\xi_m$. Individuals at stratum $X$ may also discontinue doxy-PEP and return to stratum $N$ at a rate $\xi_{xn}$. Individuals at strata $N$ and $X$ may initiate or reinitiate doxy-PEP following a sexual health clinic visit - either through screening or diagnosis - with probabilities $p^{DoS}$ and $p^{DoD}$, respectively, and transition into stratum $D$.

Model of doxy-PEP:

1. For non-doxy-PEP (N):

$$\frac{dU_j^N(t)}{dt} = q_j\alpha(1-p^{DbE}) - \left(\lambda_j(t) + p_j^{DoS}\eta_j(t) + \frac{1}{\gamma}\right)U_j^N(t) + \left(1-p_j^{DoD}\right)\rho R_j^N(t) + \xi_n U_j^D(t) + \xi_{xn}U_j^X(t) \quad (13)$$

$$\frac{dI_j^N(t)}{dt} = \lambda_j(t)U_j^N(t) - \left(\sigma + \frac{1}{\gamma}\right)I_j^N(t) + \xi_n I_j^D(t) + \xi_{xn}I_j^X(t) \quad (14)$$

$$\frac{dP_j^N(t)}{dt} = \sigma I_j^N(t) - \left(\mu + \psi_S + \frac{1}{\gamma}\right)P_j^N(t) + \xi_n P_j^D(t) + \xi_{xn}P_j^X(t) \quad (15)$$

$$\frac{dS_j^N(t)}{dt} = \psi_S P_j^N(t) - \left(\mu + \psi_E + \frac{1}{\gamma}\right)S_j^N(t) + \xi_n S_j^D(t) + \xi_{xn}S_j^X(t) \quad (16)$$

$$\frac{dE_j^N(t)}{dt} = \psi_E S_j^N(t) - \left(\eta_j(t) + \psi_L + \frac{1}{\gamma}\right)E_j^N(t) + \xi_n E_j^D(t) + \xi_{xn}E_j^X(t) \quad (17)$$

$$\frac{dL_j^N(t)}{dt} = \psi_L E_j^N(t) - \left(\eta_j(t) + \psi_T + \frac{1}{\gamma}\right)L_j^N(t) + \xi_n L_j^D(t) + \xi_{xn}L_j^X(t) \quad (18)$$

$$\frac{dT_j^N(t)}{dt} = \psi_T L_j^N(t) - \left(\mu + \nu + \frac{1}{\gamma}\right)T_j^N(t) + \xi_n T_j^D(t) + \xi_{xn}T_j^X(t) \quad (19)$$

$$\frac{dR_j^N(t)}{dt} = \mu\left(P_j^N(t) + S_j^N(t) + T_j^N(t)\right) + \eta_j(t)\left(E_j^N(t) + L_j^N(t)\right) - \left(\rho + \frac{1}{\gamma}\right)R_j^N(t) + \xi_n R_j^D(t) + \xi_{xn}R_j^X(t) \quad (20)$$

2. For doxy-inconsistent (X):

$$\frac{dU_j^X(t)}{dt} = \xi_x U_j^D(t) + \left(1-p_j^{DoD}\right)\rho R_j^X(t) - \left((1-\zeta e_d)\lambda_j(t) + \frac{1}{\gamma} + \xi_{xn} + p_j^{DoS}\eta_j(t)\right)U_j^X(t) \quad (21)$$

$$\frac{dI_j^X(t)}{dt} = \xi_x I_j^D(t) + (1-\zeta e_d)\lambda_j(t)U_j^X(t) - \left(\sigma + \frac{1}{\gamma} + \xi_{xn}\right)I_j^X(t) \quad (22)$$

$$\frac{dP_j^X(t)}{dt} = \xi_x P_j^D(t) + \sigma I_j^X(t) - \left(\mu + \psi_S + \frac{1}{\gamma} + \xi_{xn}\right)P_j^X(t) \quad (23)$$

$$\frac{dS_j^X(t)}{dt} = \xi_x S_j^D(t) + \psi_S P_j^X(t) - \left(\mu + \psi_E + \frac{1}{\gamma} + \xi_{xn}\right)S_j^X(t) \quad (24)$$

$$\frac{dE_j^X(t)}{dt} = \xi_x E_j^D(t) + \psi_E S_j^X(t) - \left(\eta_j(t) + \psi_L + \frac{1}{\gamma} + \xi_{xn}\right)E_j^X(t) \quad (25)$$

$$\frac{dL_j^X(t)}{dt} = \xi_x L_j^D(t) + \psi_L E_j^X(t) - \left(\eta_j(t) + \psi_T + \frac{1}{\gamma} + \xi_{xn}\right)L_j^X(t) \quad (26)$$

$$\frac{dT_j^X(t)}{dt} = \xi_x T_j^D(t) + \psi_T L_j^X(t) - \left(\mu + \nu + \frac{1}{\gamma} + \xi_{xn}\right)T_j^X(t) \quad (27)$$

$$\frac{dR_j^X(t)}{dt} = \xi_x R_j^D(t) + \mu\left(P_j^X(t) + S_j^X(t) + T_j^X(t)\right) + \eta_j(t)\left(E_j^X(t) + L_j^X(t)\right) - \left(\rho + \frac{1}{\gamma} + \xi_{xn}\right)R_j^X(t) \quad (28)$$

3. For doxy-PEP (D):

$$\frac{dU_j^D(t)}{dt} = q_j\alpha p^{DbE} + p_j^{DoS}\eta_j(t)(U_j^N(t) + U_j^X(t)) + p_j^{DoD}\rho(R_j^N(t) + R_j^X(t)) + \rho R_j^D(t) - \left((1-e_d)\lambda_j(t) + \frac{1}{\gamma} + \xi_x + \xi_n + \xi_m\right)U_j^D(t) \quad (29)$$

$$\frac{dI_j^D(t)}{dt} = (1-e_d)\lambda_j(t)U_j^D(t) - \left(\sigma + \frac{1}{\gamma} + \xi_x + \xi_n + \xi_m\right)I_j^D(t) \quad (30)$$

$$\frac{dP_j^D(t)}{dt} = \sigma I_j^D(t) - \left(\mu + \psi_S + \frac{1}{\gamma} + \xi_x + \xi_n + \xi_m\right) P_j^D(t) \tag{31}$$

$$\frac{dS_j^D(t)}{dt} = \psi_S P_j^D(t) - \left(\mu + \psi_E + \frac{1}{\gamma} + \xi_x + \xi_n + \xi_m\right) S_j^D(t) \tag{32}$$

$$\frac{dE_j^D(t)}{dt} = \psi_E S_j^D(t) - \left(\eta_j(t) + \psi_L + \frac{1}{\gamma} + \xi_x + \xi_n + \xi_m\right) E_j^D(t) \tag{33}$$

$$\frac{dL_j^D(t)}{dt} = \psi_L E_j^D(t) - \left(\eta_j(t) + \psi_T + \frac{1}{\gamma} + \xi_x + \xi_n + \xi_m\right) L_j^D(t) \tag{34}$$

$$\frac{dT_j^D(t)}{dt} = \psi_T L_j^D(t) - \left(\mu + \nu + \frac{1}{\gamma} + \xi_x + \xi_n + \xi_m\right) T_j^D(t) \tag{35}$$

$$\frac{dR_j^D(t)}{dt} = \mu\left(P_j^D(t) + S_j^D(t) + T_j^D(t)\right) + \eta_j(t)\left(E_j^D(t) + L_j^D(t)\right) - \left(\rho + \frac{1}{\gamma} + \xi_x + \xi_n + \xi_m\right) R_j^D(t) \tag{36}$$

4. For doxy-intolerant (M):

$$\frac{dU_j^M(t)}{dt} = \xi_m U_j^D(t) + \rho R_j^M(t) - \left(\lambda_j(t) + \frac{1}{\gamma}\right) U_j^M(t) \tag{37}$$

$$\frac{dI_j^M(t)}{dt} = \xi_m I_j^D(t) + \lambda_j(t) U_j^M(t) - \left(\sigma + \frac{1}{\gamma}\right) I_j^M(t) \tag{38}$$

$$\frac{dP_j^M(t)}{dt} = \xi_m P_j^D(t) + \sigma I_j^M(t) - \left(\mu + \psi_S + \frac{1}{\gamma}\right) P_j^M(t) \tag{39}$$

$$\frac{dS_j^M(t)}{dt} = \xi_m S_j^D(t) + \psi_S P_j^M(t) - \left(\mu + \psi_E + \frac{1}{\gamma}\right) S_j^M(t) \tag{40}$$

$$\frac{dE_j^M(t)}{dt} = \xi_m E_j^D(t) + \psi_E S_j^M(t) - \left(\eta_j(t) + \psi_L + \frac{1}{\gamma}\right) E_j^M(t) \tag{41}$$

$$\frac{dL_j^M(t)}{dt} = \xi_m L_j^D(t) + \psi_L E_j^M(t) - \left(\eta_j(t) + \psi_T + \frac{1}{\gamma}\right) L_j^M(t) \tag{42}$$

$$\frac{dT_j^M(t)}{dt} = \xi_m T_j^D(t) + \psi_T L_j^M(t) - \left(\mu + \nu + \frac{1}{\gamma}\right) T_j^M(t) \tag{43}$$

$$\frac{dR_j^M(t)}{dt} = \xi_m R_j^D(t) + \mu\left(P_j^M(t) + S_j^M(t) + T_j^M(t)\right) + \eta_j(t)\left(E_j^M(t) + L_j^M(t)\right) - \left(\rho + \frac{1}{\gamma}\right) R_j^M(t) \tag{44}$$

For each doxy-PEP stratum $i \in \{N, X, D, M\}$ and calendar year $t$, the total number of recovered cases $Y_R^i(t)$, the number of primary-stage cases $Y_P^i(t)$, the number of secondary-stage cases $Y_S^i(t)$, the number of early-latent-stage cases $Y_E^i(t)$, the number of late-latent-stage cases $Y_L^i(t)$, the number of tertiary-stage cases $Y_T^i(t)$, the number of unaffected patients screened for syphilis $Y_U^i(t)$, are as follows:

$$Y_R^i(t) = \sum_{j\in\{L,H\}} \left(\int_t^{t+1} \rho R_j^i(\tau) d\tau\right) \tag{53}$$

$$Y_I^i(t) = \sum_{j\in\{L,H\}} \left(\int_t^{t+1} \sigma I_j^i(\tau) d\tau\right) \tag{53}$$

$$Y_P^i(t) = \sum_{j\in\{L,H\}} \left(\int_t^{t+1} (\mu + \psi_S) P_j^i(\tau) d\tau\right) \tag{54}$$

$$Y_S^i(t) = \sum_{j\in\{L,H\}} \left(\int_t^{t+1} (\mu + \psi_E) S_j^i(\tau) d\tau\right) \tag{55}$$

$$Y_E^i(t) = \sum_{j\in\{L,H\}} \left(\int_t^{t+1} (\eta_j(\tau) + \psi_L) E_j^i(\tau) d\tau\right) \tag{56}$$

$$Y_L^i(t) = \sum_{j\in\{L,H\}} \left( \int_t^{t+1} (\eta_j(\tau) + \psi_T) L_j^i(\tau) d\tau \right) \quad (57)$$

$$Y_T^i(t) = \sum_{j\in\{L,H\}} \left( \int_t^{t+1} (\mu + \nu) T_j^i(\tau) d\tau \right) \quad (58)$$

$$Y_U^i(t) = \sum_{j\in\{L,H\}} \left( \int_t^{t+1} \eta_j(\tau) U_j^i(\tau) d\tau \right) \quad (59)$$

While specific data on doxy-PEP adherence are limited, insights can be drawn from studies on HIV pre-exposure prophylaxis (PrEP), which face similar adherence and discontinuation challenges. Although HIV PrEP and doxy-PEP differ in regimen (daily vs. event-driven or post-exposure), behavioural patterns related to adherence and reinitiation can still provide qualitative insight. Assuming exponential behaviour, a constant annual transition rate can be derived from the proportion of individuals who restart over a given period, using the formula:

$$Aunual\ hazard\ rate = -\frac{\ln(1-p)}{t}, \quad (61)$$

where $p$ is the proportion of individuals who transit to other compartments within the time interval and $t$ is the duration of the time interval in years. For modelling purposes, it is assumed that all doxycycline is obtained through STI clinics and that reinitiation of doxy-PEP after discontinuation occurs only upon visiting an STI clinic.

Table 7: Doxy-PEP parameters used in scenario analysis. We examined three uptake levels: low (10%), moderate (33%), and high (100%), along with three adherence behaviour behavioural patterns, namely, low ($u$ = 2), normal ($u$ = 1), and high ($u$ = 0.5). A high adherence behavioural pattern corresponds to low rates of both discontinuation and suboptimal adherence, whereas a low adherence behavioural pattern reflects high rates of both. The different adherence levels are defined relative to HIV PrEP, with 'normal' corresponding to typical adherence observed in HIV PrEP studies.

| | Definition | Value | Source |
|---|---|---|---|
| $e_d$ | Efficacy of doxycycline against infections | 83% | [16] |
| $\zeta$ | Scaling factor accounting for doxycycline inefficacy due to inconsistent and irregular usage | 10.0%, 33.0%, 66.0% | - |
| $\xi_m$ | Intolerance rate of doxycycline | 0.0135 | [16][9] |
| $\xi_n$ | Discontinuation rate of doxy-PEP | $0.362u$ | [17][10] |
| $\xi_x$ | Suboptimal adherence rate of doxy-PEP | $0.420u$ | [17][11] |
| $\xi_{xn}$ | Discontinuation rate among suboptimally adherent doxy-PEP | $0.936u$ | -[12] |
| $p^{DbE}$ | Probability of uptake of doxycycline before entry into the sexually-active population | 10.0%, 33.0%, 66.0% | - |
| $p_j^{DoD}$ | Probability of uptake of doxycycline on diagnosis in group $j$ | 10.0%, 33.0%, 66.0% | - |
| $p_j^{DoS}$ | Probability of uptake of doxycycline on screening with negative results in group $j$ | 10.0%, 33.0%, 66.0% | - |

[9] In an 18-month randomized controlled trial, 2% of participants discontinued doxy-PEP due to gastrointestinal adverse events (doxycycline intolerance).

[10] 30.4% of gay or bisexual MSM and transgender women discontinued HIV PrEP within one year.

[11] 34.3% of gay or bisexual MSM and transgender women exhibited suboptimal adherence for HIV PrEP within one year.

[12] Assumed a probability of dropout twice as high for fully adherent doxy-PEP (i.e., 60.8% of MSM discontinued within one year).

**Appendix D: Results**

1. Performance metrics

To evaluate the effectiveness of each doxy-PEP strategy, we compute the total number of syphilis cases at year $t$ across all doxy-PEP strata $i \in \{N, X, D, M\}$, denoted as:

$$Y_C(t) = \sum_i [Y_I^i(t)] \tag{62}$$

We compare this to the corresponding baseline without intervention, $\hat{Y}_C(t)$, to calculate the total number of averted syphilis cases over $M$ years (starting from year $t_0$):

$$\sum_{t=1}^{M} [\hat{Y}_C(t_0 + t) - Y_C(t_0 + t)] \tag{63}$$

In addition, we calculate the total number of doxy-PEP prescriptions administered over $M$ years (starting from year $t_0$):

$$\sum_{t=1}^{M} [\alpha p^{DbE} + p_j^{DoS}(Y_U^N(t_0 + t) + Y_U^X(t_0 + t)) + p_j^{DoD}(Y_R^N(t_0 + t) + Y_R^X(t_0 + t))] \tag{64}$$

Furthermore, we calculate the number of syphilis cases averted per doxy-PEP prescription over a period of $M$ years, beginning from year $t_0$:

$$\sum_{t=1}^{M} \frac{\hat{Y}_C(t_0 + t) - Y_C(t_0 + t)}{\alpha p^{DbE} + p_j^{DoS}(Y_U^N(t_0 + t) + Y_U^X(t_0 + t)) + p_j^{DoD}(Y_R^N(t_0 + t) + Y_R^X(t_0 + t))} \tag{65}$$

Next, to better understand the transmission dynamics and inform early detection strategies, we compute the total number of syphilis cases at year $t$ by stage - primary, secondary, and other - aggregated across all doxy-PEP strata $i \in \{N, X, D, M\}$, denoted respectively as:

$$Y_P(t) = \sum_i Y_P^i(t) \tag{66}$$

$$Y_S(t) = \sum_i Y_S^i(t) \tag{67}$$

$$Y_O(t) = \sum_i [Y_E^i(t) + Y_L^i(t) + Y_T^i(t)] \tag{68}$$

We compare these values to their corresponding baselines without intervention, denoted as $\hat{Y}_P(t)$, $\hat{Y}_S(t)$, and $\hat{Y}_O(t)$, to compute the total number of averted syphilis cases over $M$ years (starting from year $t_0$) by stage - primary, secondary, and other:

$$\sum_{t=1}^{M} [\hat{Y}_P(t_0 + t) - Y_P(t_0 + t)] \tag{69}$$

$$\sum_{t=1}^{M} [\hat{Y}_S(t_0 + t) - Y_S(t_0 + t)] \tag{70}$$

$$\sum_{t=1}^{M}[\hat{Y}_O(t_0+t)-Y_O(t_0+t)] \quad (71)$$

2. Results of Singapore (main scenario) and England

This subsection presents results under the following assumptions: (1) protection is reduced to 33.0% of the baseline efficacy for suboptimal adherence stratum (i.e., $\zeta$ = 33.0%) and (2) stabilization of inferred time-varying behavioural trends. The outcomes include the total number of averted syphilis cases, total number of doxy-PEP prescriptions, number of averted cases per prescription, stage-specific averted cases, annual syphilis incidence, and the annual number of susceptible MSM in the high-risk group. These results are stratified by varying levels of uptake, asymptomatic screening rate, and adherence behavioural pattern.

Table 8: Singapore: estimated impact and efficiency of doxy-PEP strategies among MSM with a normal adherence behavioural pattern (i.e., $u = 1$), where protection is reduced to 33.0% of the baseline efficacy for suboptimal adherence strata (i.e., $\zeta = 33.0\%$), assuming stabilization of inferred time-varying behavioural trends in the main scenario (i.e., assuming all males not engaging in sex with females are classified as MSM). Results are presented as median (95% credible interval), assuming a high asymptomatic screening rate (taken from our calibration, aligned with CDC-recommended levels [18]).

| | Total number of averted cases, thousands: Uptake rate | | | Number of doxy-PEP prescriptions, thousands: Uptake rate | | | Number of averted cases per prescription: Uptake rate | | |
|---|---|---|---|---|---|---|---|---|---|
| | 10.0% | 33.0% | 66.0% | 10.0% | 33.0% | 66.0% | 10.0% | 33.0% | 66.0% |
| DbE | 2.5 (0.6, 6.1) | 7.3 (1.6, 19.2) | 12.5 (2.8, 35.8) | 3.8 (3.8, 3.8) | 12.5 (12.5, 12.5) | 25.0 (25, 25) | 0.66 (0.15, 1.60) | 0.59 (0.13, 1.53) | 0.50 (0.11, 1.43) |
| DoD(H) | 4.4 (0.4, 27.8) | 10.0 (1.1, 53.2) | 14.0 (1.9, 67.1) | 1.7 (0.6, 4.7) | 4.0 (1.6, 9.8) | 5.9 (2.8, 13) | 2.64 (0.70, 6.29) | 2.50 (0.68, 5.94) | 2.35 (0.65, 5.62) |
| DoD | 4.4 (0.4, 27.9) | 10.1 (1.2, 53.2) | 14.0 (2, 67.3) | 2.0 (0.7, 6.3) | 4.8 (2, 12.8) | 7.1 (3.3, 16.9) | 2.20 (0.56, 5.12) | 2.08 (0.54, 4.88) | 1.96 (0.52, 4.68) |
| DoA(H) | 24.4 (6.6, 93.1) | 24.7 (6.8, 93.1) | 24.8 (6.9, 93.1) | 244.9 (15.6, 396.2) | 358.7 (46.3, 659.4) | 454.4 (82.1, 1012.7) | 0.12 (0.03, 1.53) | 0.08 (0.02, 0.63) | 0.06 (0.01, 0.38) |
| DoA | 24.4 (6.6, 93.1) | 24.7 (6.8, 93.1) | 24.8 (6.9, 93.1) | 853.8 (42.3, 1568.7) | 1390.7 (130.5, 2390.9) | 1732.9 (241.1, 3376.4) | 0.03 (0.01, 0.58) | 0.02 (0.00, 0.23) | 0.02 (0.00, 0.13) |
| DaR | 24.4 (6.6, 93.1) | 24.7 (6.8, 93.1) | 24.8 (6.9, 93.1) | 244.9 (15.7, 396.2) | 358.7 (46.4, 659.4) | 454.4 (82.3, 1012.8) | 0.12 (0.03, 1.51) | 0.08 (0.02, 0.63) | 0.06 (0.01, 0.37) |

Table 9: Singapore: estimated impact of doxy-PEP strategies (total number of averted cases by stage) among MSM with a normal adherence behavioural pattern (i.e., $u = 1$), where protection is reduced to 33.0% of the baseline efficacy for suboptimal adherence strata (i.e., $\zeta = 33.0\%$), assuming stabilization of inferred time-varying behavioural trends in the main scenario (i.e., assuming all males not engaging in sex with females are classified as MSM). Results are presented as median (95% credible interval), assuming a high asymptomatic screening rate (taken from our calibration, aligned with CDC-recommended levels [18]).

| | Primary stage, thousands: Uptake rate | | | Secondary stage, thousands: Uptake rate | | | Other stages: Uptake rate | | | Diagnosed, thousands: Uptake rate | | |
|---|---|---|---|---|---|---|---|---|---|---|---|---|
| | 10.0% | 33.0% | 66.0% | 10.0% | 33.0% | 66.0% | 10.0% | 33.0% | 66.0% | 10.0% | 33.0% | 66.0% |
| DbE | 2.5 | 7.4 | 11.5 | 0.1 | 0.3 | 0.6 | 1 | 3 | 6 | 2.5 | 7.3 | 12.5 |
| | (0.6, 6.1) | (1.6, 17.2) | (2.8, 35.9) | (0, 0.6) | (0.1, 0.8) | (0.1, 3.2) | (0, 14) | (0, 42) | (0, 76) | (0.6, 6.1) | (1.6, 19) | (2.8, 35.8) |
| DoD(H) | 4.5 | 10.1 | 14.0 | 0.2 | 0.5 | 0.7 | 2 | 5 | 7 | 4.4 | 10.0 | 13.9 |
| | (0.4, 27.9) | (1.1, 53.6) | (2.0, 67.5) | (0, 2.2) | (0, 4.0) | (0.1, 5.0) | (0, 42) | (0, 83) | (0, 106) | (0.4, 27.7) | (1.1, 53.1) | (1.9, 67) |
| DoD | 4.5 | 10.1 | 14.0 | 0.2 | 0.5 | 0.7 | 2 | 5 | 7 | 4.4 | 10.0 | 13.9 |
| | (0.4, 27.9) | (1.2, 53.8) | (2.0, 67.6) | (0, 2.2) | (0, 4.0) | (0.1, 5.0) | (0, 42) | (0, 83) | (0, 106) | (0.4, 27.7) | (1.1, 53.3) | (1.9, 67.1) |
| DoA(H) | 24.5 | 24.8 | 23.8 | 1.1 | 1.2 | 1.2 | 1 | 12 | 12 | 24.3 | 24.7 | 24.7 |
| | (6.6, 93.8) | (6.8, 94.2) | (6.9, 94.2) | (0.2, 6.7) | (0.2, 6.7) | (0.2, 6.7) | (12, 165) | (1, 167) | (1, 167) | (6.6, 93.2) | (6.8, 93.6) | (6.9, 93.6) |
| DoA | 24.5 | 24.8 | 24.8 | 1.1 | 1.2 | 1.2 | 1 | 12 | 12 | 24.3 | 24.7 | 24.7 |
| | (6.6, 93.8) | (6.9, 94.2) | (6.9, 94.2) | (0.2, 6.7) | (0.2, 6.7) | (0.2, 6.7) | (12, 165) | (1, 167) | (1, 167) | (6.6, 93.2) | (6.8, 93.6) | (6.9, 93.6) |
| DaR | 24.5 | 24.8 | 23.8 | 1.1 | 1.1 | 1.2 | 1 | 12 | 12 | 24.3 | 24.7 | 24.7 |
| | (6.6, 93.8) | (6.8, 94.2) | (6.9, 94.2) | (0.2, 6.7) | (0.2, 6.6) | (0.2, 6.7) | (12, 165) | (1, 167) | (1, 167) | (6.6, 93.2) | (6.8, 93.6) | (6.9, 93.6) |

Table 10: England: estimated impact and efficiency of doxy-PEP strategies among MSM with a normal adherence behavioural pattern (i.e., $u = 1$), where protection is reduced to 33.0% of the baseline efficacy for suboptimal adherence strata (i.e., $\zeta = 33.0\%$), assuming stabilization of inferred time-varying behavioural trends. Results are presented as median (95% credible interval), assuming a high asymptomatic screening rate (taken from our calibration, aligned with CDC-recommended levels [18]).

| | Total number of averted cases, thousands: Uptake rate | | | Number of doxy-PEP prescriptions, thousands: Uptake rate | | | Number of averted cases per prescription: Uptake rate | | |
|---|---|---|---|---|---|---|---|---|---|
| | 10.0% | 33.0% | 66.0% | 10.0% | 33.0% | 66.0% | 10.0% | 33.0% | 66.0% |
| DbE | 13.8 (6.6, 23) | 44.5 (20.6, 74.6) | 84.7 (37.6, 146.7) | 18.0 (18.0, 18.0) | 59.4 (59.4, 59.4) | 118.8 (118.8, 118.8) | 0.77 (0.37, 1.28) | 0.75 (0.35, 1.26) | 0.71 (0.31, 1.24) |
| DoD(H) | 87.4 (21.9, 293.2) | 165 (49.3, 503.7) | 206.2 (67.8, 595.3) | 17.9 (8.1, 42.4) | 35.6 (17.8, 77.2) | 46.3 (24.2, 100.9) | 4.82 (2.24, 7.99) | 4.60 (2.12, 7.79) | 4.38 (2.00, 7.56) |
| DoD | 87.4 (21.9, 293.3) | 165 (49.3, 503.7) | 206.2 (67.8, 595.3) | 19.7 (8.1, 48.8) | 38.4 (19.9, 87.7) | 51.1 (27.7, 112) | 4.43 (2.04, 7.28) | 4.19 (1.92, 7.01) | 3.99 (1.81, 6.83) |
| DoA(H) | 273.9 (105.5, 709.2) | 279.8 (109.1, 724) | 280.9 (109.8, 742.5) | 874.7 (82, 1452.1) | 1308 (215.8, 2336.9) | 1636.4 (361.3, 3497.5) | 0.34 (0.10, 3.21) | 0.21 (0.07, 1.38) | 0.16 (0.05, 0.94) |
| DoA | 273.9 (105.5, 709.3) | 279.8 (109.2, 724) | 280.9 (109.8, 742.5) | 4059.1 (246.3, 7909.2) | 6919.2 (724.1, 11785.3) | 8643.8 (1311.2, 16245.6) | 0.07 (0.02, 1.17) | 0.04 (0.01, 0.46) | 0.03 (0.01, 0.26) |
| DaR | 273.9 (105.5, 709.2) | 279.8 (109.1, 724) | 280.9 (109.8, 742.5) | 874.7 (82.4, 1452.2) | 1308.1 (216.5, 2337.1) | 1636.9 (361.8, 3497.9) | 0.34 (0.10, 3.20) | 0.21 (0.07, 1.38) | 0.16 (0.05, 0.94) |

Table 11: England: estimated impact of doxy-PEP strategies (total number of averted cases by stage) among MSM with a normal adherence behavioural pattern (i.e., $u$ = 1), where protection is reduced to 33.0% of the baseline efficacy for suboptimal adherence strata (i.e., $\zeta$ = 33.0%), assuming stabilization of inferred time-varying behavioural trends. Results are presented as median (95% credible interval), assuming a high asymptomatic screening rate (taken from our calibration, aligned with CDC-recommended levels [18]).

| | Primary stage, thousands: Uptake rate | | | Secondary stage, thousands: Uptake rate | | | Other stages: Uptake rate | | | Diagnosed, thousands: Uptake rate | | |
|---|---|---|---|---|---|---|---|---|---|---|---|---|
| | 10.0% | 33.0% | 66.0% | 10.0% | 33.0% | 66.0% | 10.0% | 33.0% | 66.0% | 10.0% | 33.0% | 66.0% |
| DbE | 13.8<br>(6.6, 23.0) | 44.4<br>(20.5, 74.6) | 84.6<br>(37.2, 145.7) | 0.7<br>(0.2, 2.4) | 2.3<br>(0.5, 7.8) | 4.2<br>(1, 15.1) | 8<br>(1, 81) | 25<br>(2, 260) | 47<br>(4, 502) | 13.7<br>(6.6, 22.9) | 44.0<br>(20.3, 74.2) | 83.9<br>(36.9, 145.4) |
| DoD(H) | 87.4<br>(21.8, 293.9) | 164.8<br>(48.6, 507.0) | 205.1<br>(67.4, 591.5) | 4.3<br>(0.7, 23.0) | 8.4<br>(1.3, 40.9) | 10.5<br>(1.7, 50.4) | 47<br>(3, 643) | 90<br>(7, 1161) | 112<br>(9, 1414) | 86.9<br>(21.6, 291.9) | 164.2<br>(48.4, 505.8) | 203.6<br>(67, 587.9) |
| DoD | 87.4<br>(21.8, 294.0) | 164.8<br>(48.6, 507.0) | 205.2<br>(67.4, 591.5) | 4.3<br>(0.7, 23.0) | 8.4<br>(1.3, 41.0) | 10.5<br>(1.7, 50.4) | 47<br>(3, 643) | 90<br>(7, 1162) | 112<br>(9, 1414) | 86.9<br>(21.6, 292) | 164.3<br>(48.4, 505.8) | 203.7<br>(67, 587.9) |
| DoA(H) | 272.7<br>(104.4, 713.4) | 276.7<br>(109.5, 724.2) | 277.2<br>(110.1, 744.6) | 14.0<br>(2.8, 62.7) | 14.2<br>(2.9, 64.0) | 14.2<br>(2.9, 64.0) | 152<br>(13, 1766) | 155<br>(14, 1815) | 156<br>(14, 1828) | 271.7<br>(104.2, 711.6) | 275.9<br>(108.9, 722.3) | 276.4<br>(109.9, 740.3) |
| DoA | 272.7<br>(104.5, 713.4) | 276.7<br>(109.5, 724.2) | 277.2<br>(110.1, 744.6) | 14.0<br>(2.8, 62.7) | 14.2<br>(2.9, 64.0) | 14.2<br>(2.9, 64.0) | 152<br>(13, 1766) | 155<br>(14, 1815) | 156<br>(14, 1828) | 271.7<br>(104.3, 711.6) | 276.0<br>(108.9, 722.3) | 276.4<br>(109.9, 740.3) |
| DaR | 272.7<br>(104.4, 713.4) | 276.7<br>(109.5, 724.2) | 277.2<br>(110.1, 744.6) | 14.0<br>(2.8, 62.7) | 14.2<br>(2.9, 64.0) | 14.2<br>(2.9, 64.0) | 152<br>(14, 1766) | 155<br>(14, 1815) | 156<br>(14, 1828) | 271.7<br>(104.2, 711.6) | 275.9<br>(108.9, 722.3) | 276.4<br>(109.9, 740.3) |

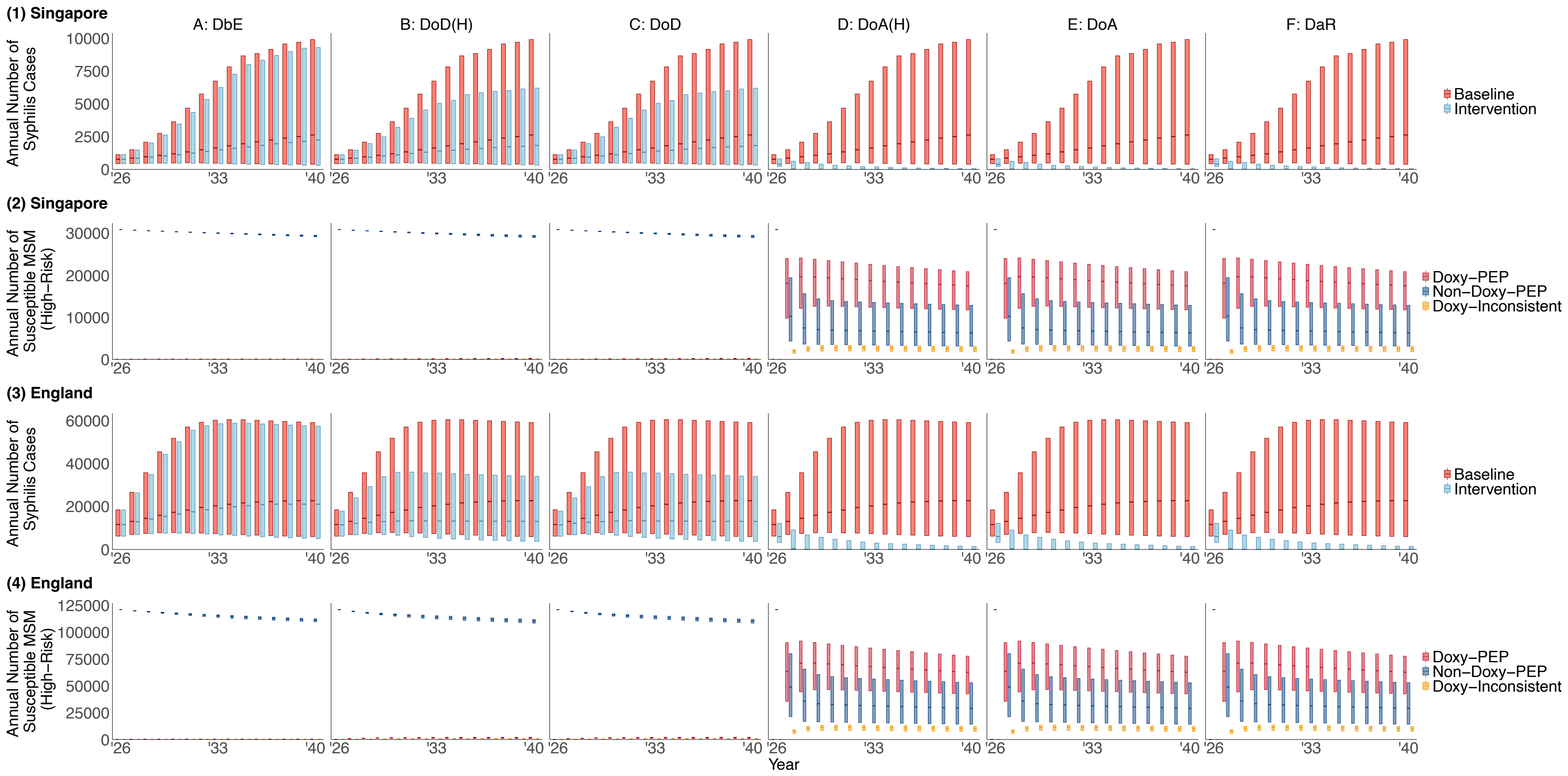


Fig. 6: From left to right, the doxy-PEP strategies are: DbE, DoD(H), DoD, DoA(H), DoA, and DaR. The Singapore main scenario assumes stabilization of inferred time-varying behavioural trends, with all males not engaging in sex with females classified as MSM. Panels (1) and (3) show the annual number of syphilis cases under intervention (uptake rate of 10.0%) and baseline scenarios for Singapore and England, respectively, assuming suboptimal adherence that reduces protection to 33.0% of baseline efficacy (i.e., $\zeta$ = 33.0%). Each boxplot shows the median (central line) and the 95% credible interval (box bounds). Panels (2) and (4) present the annual number of susceptible MSM at high-risk group for Singapore and England, respectively. Each boxplot shows the median (central line) and the interquartile range (box bounds).

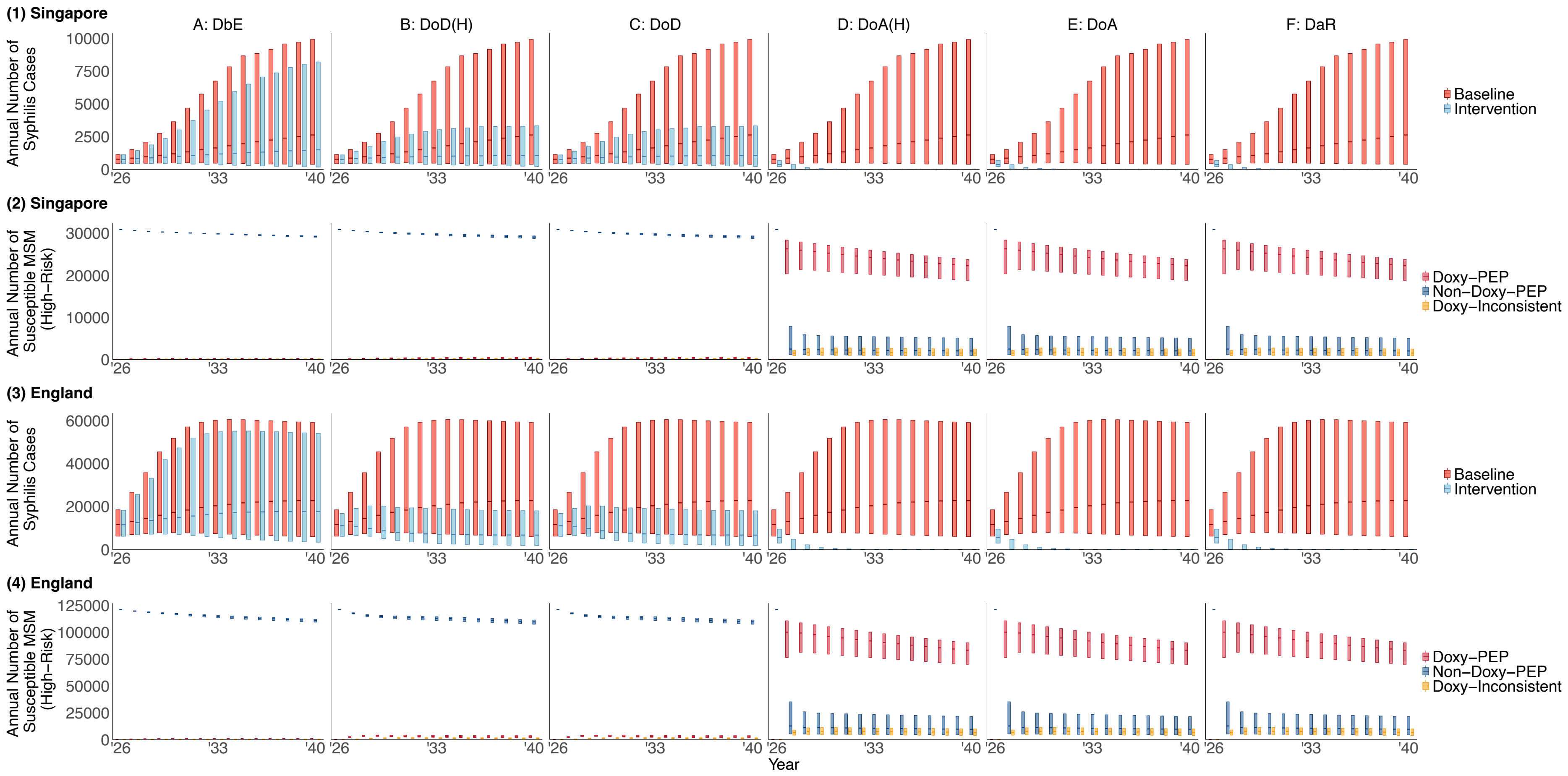


Fig. 7: From left to right, the doxy-PEP strategies are: DbE, DoD(H), DoD, DoA(H), DoA, and DaR. The Singapore main scenario assumes stabilization of inferred time-varying behavioural trends, with all males not engaging in sex with females classified as MSM. Panels (1) and (3) show the annual number of syphilis cases under intervention (uptake rate of 33.0%) and baseline scenarios for Singapore and England, respectively, assuming suboptimal adherence that reduces protection to 33.0% of baseline efficacy (i.e., $\zeta$ = 33.0%). Each boxplot shows the median (central line) and the 95% credible interval (box bounds). Panels (2) and (4) present the annual number of susceptible MSM at high-risk group for Singapore and England, respectively. Each boxplot shows the median (central line) and the interquartile range (box bounds).

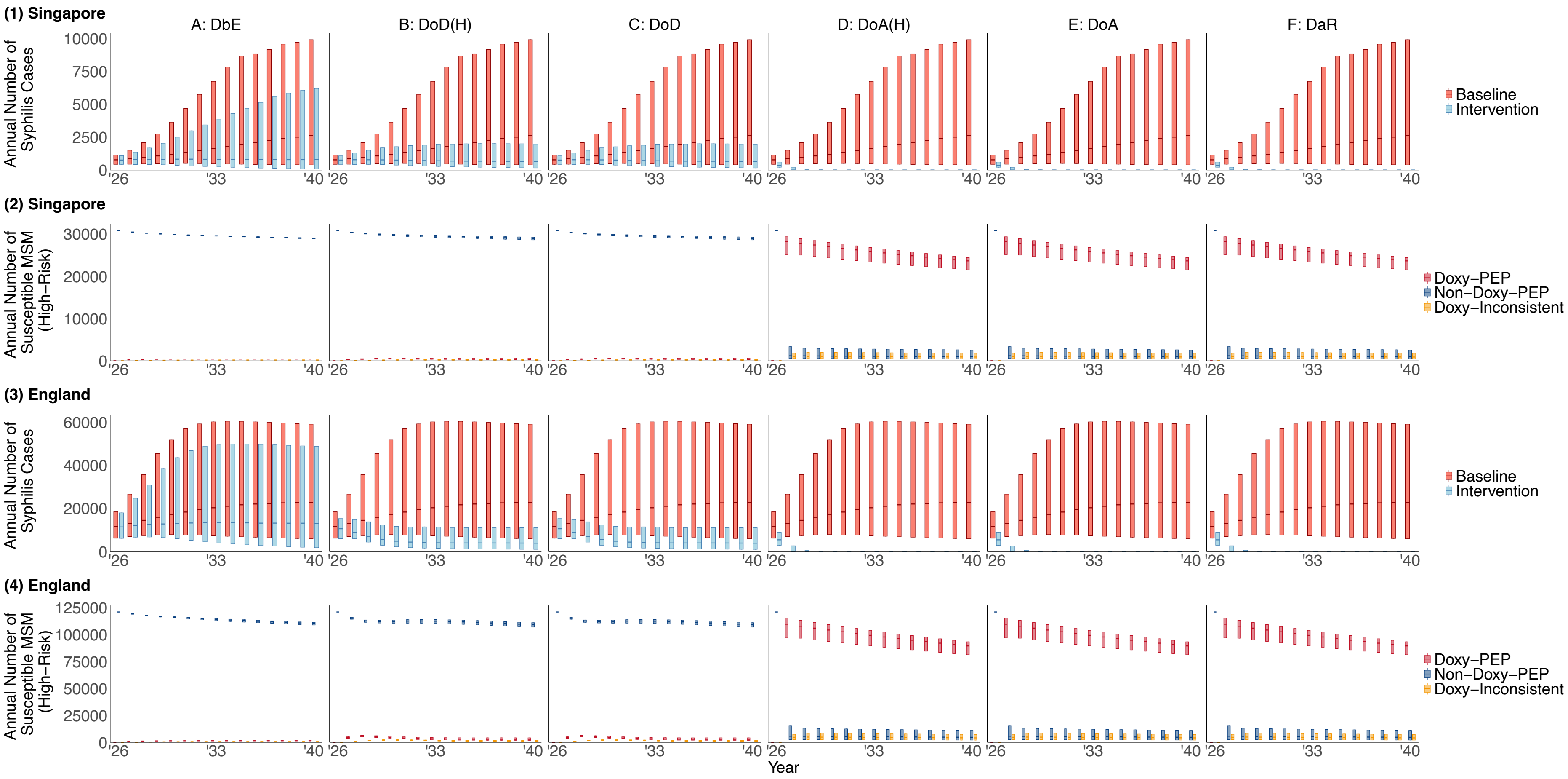


Fig. 8: From left to right, the doxy-PEP strategies are: DbE, DoD(H), DoD, DoA(H), DoA, and DaR. The Singapore main scenario assumes stabilization of inferred time-varying behavioural trends, with all males not engaging in sex with females classified as MSM. Panels (1) and (3) show the annual number of syphilis cases under intervention (uptake rate of 66.0%) and baseline scenarios for Singapore and England, respectively, assuming suboptimal adherence that reduces protection to 33.0% of baseline efficacy (i.e., $\zeta$ = 33.0%). Each boxplot shows the median (central line) and the 95% credible interval (box bounds). Panels (2) and (4) present the annual number of susceptible MSM at high-risk group for Singapore and England, respectively. Each boxplot shows the median (central line) and the interquartile range (box bounds).

Next, we compare the results of the strategy under high vs. low asymptomatic screening rate to investigate the effectiveness of doxy-PEP strategies if asymptomatic screening rates drop.

Table 12: Singapore: estimated impact of doxy-PEP strategies among MSM with a normal adherence behavioural pattern (i.e., $u$ = 1), where protection is reduced to 33.0% of the baseline efficacy for suboptimal adherence strata (i.e., $\zeta$ = 33.0%), assuming stabilization of inferred time-varying behavioural trends in the main scenario (i.e., assuming all males not engaging in sex with females are classified as MSM). Results are presented as median (95% credible interval), assuming a low asymptomatic screening rate as informed by [19].

| | Total number of averted cases, thousands: Uptake rate | | | Number of doxy-PEP prescriptions, thousands: Uptake rate | | | Number of averted cases per prescription: Uptake rate | | |
|---|---|---|---|---|---|---|---|---|---|
| | 10.0% | 33.0% | 66.0% | 10.0% | 33.0% | 66.0% | 10.0% | 33.0% | 66.0% |
| DbE | 3.3<br>(0.6, 7) | 10.0<br>(1.9, 22.8) | 17.4<br>(3.3, 43.8) | 3.8<br>(3.8, 3.8) | 12.5<br>(12.5, 12.5) | 25.0<br>(25.0, 25.0) | 0.88<br>(0.17, 1.85) | 0.80<br>(0.15, 1.82) | 0.70<br>(0.13, 1.75) |
| DoD(H) | 7.6<br>(0.5, 52.7) | 16.2<br>(1.4, 96.5) | 22.0<br>(2.4, 118.3) | 2.3<br>(0.7, 7.7) | 5.2<br>(1.9, 15.1) | 7.5<br>(3.1, 20) | 3.20<br>(0.79, 6.89) | 3.04<br>(0.76, 6.63) | 2.87<br>(0.73, 6.35) |
| DoD | 7.6<br>(0.5, 52.8) | 16.2<br>(1.4, 96.6) | 22<br>(2.4, 118.3) | 2.8<br>(0.8, 10.0) | 6.5<br>(2.3, 19.5) | 9.2<br>(3.8, 25.6) | 2.58<br>(0.62, 5.62) | 2.43<br>(0.60, 5.36) | 2.31<br>(0.57, 5.13) |
| DoA(H) | 23.6<br>(4.3, 88.2) | 32.6<br>(6.8, 143.0) | 34.2<br>(7.3, 151.4) | 8.5<br>(7.9, 12.5) | 24.6<br>(24.3, 27.3) | 45.6<br>(45.4, 46.4) | 2.77<br>(0.54, 7.16) | 1.32<br>(0.28, 5.33) | 0.75<br>(0.16, 3.27) |
| DoA | 23.8<br>(4.3, 88.3) | 32.7<br>(6.8, 143.4) | 34.3<br>(7.3, 151.4) | 21.4<br>(14.2, 32.5) | 64.5<br>(42.9, 96.6) | 122.4<br>(81.2, 182.2) | 1.14<br>(0.20, 3.60) | 0.51<br>(0.10, 2.17) | 0.28<br>(0.06, 1.22) |
| DaR | 23.6<br>(4.3, 88.2) | 32.6<br>(6.8, 143.0) | 34.2<br>(7.3, 151.4) | 8.7<br>(8, 13.9) | 24.8<br>(24.4, 28.4) | 45.8<br>(45.5, 46.9) | 2.70<br>(0.53, 6.37) | 1.31<br>(0.28, 5.04) | 0.75<br>(0.16, 3.27) |

Table 13: England: estimated impact of doxy-PEP strategies among MSM with a normal adherence behavioural pattern (i.e., $u$ = 1), where protection is reduced to 33.0% of the baseline efficacy for suboptimal adherence strata (i.e., $\zeta$ = 33.0%), assuming stabilization of inferred time-varying behavioural trends. Results are presented as median (95% credible interval), assuming a low asymptomatic screening rate as informed by [15].

| | Total number of averted cases, thousands: Uptake rate | | | Number of doxy-PEP prescriptions, thousands: Uptake rate | | | Number of averted cases per prescription: Uptake rate | | |
|---|---|---|---|---|---|---|---|---|---|
| | 10.0% | 33.0% | 66.0% | 10.0% | 33.0% | 66.0% | 10.0% | 33.0% | 66.0% |
| DbE | 14.8 (7.2, 23.5) | 48.0 (22.9, 76.8) | 92.5 (42.4, 150.9) | 18.0 (18.0, 18.0) | 59.4 (59.4, 59.4) | 118.8 (118.8, 118.8) | 0.82 (0.40, 1.31) | 0.81 (0.39, 1.29) | 0.78 (0.36, 1.27) |
| DoD(H) | 113.5 (26.9, 370.7) | 214.5 (58.9, 632.9) | 265.9 (79.4, 759.3) | 22.6 (9.5, 55.4) | 45.0 (21.9, 100.8) | 58.5 (30.2, 123.9) | 5.05 (2.43, 8.02) | 4.82 (2.30, 7.76) | 4.59 (2.18, 7.52) |
| DoD | 113.5 (26.9, 370.8) | 214.5 (58.9, 632.9) | 265.9 (79.4, 759.3) | 25.1 (10.8, 59.5) | 49.3 (24.5, 111.2) | 64.5 (33.8, 140.7) | 4.56 (2.17, 7.34) | 4.37 (2.06, 7.11) | 4.18 (1.93, 6.87) |
| DoA(H) | 210.2 (74.7, 503) | 323.7 (111.8, 821.5) | 346.8 (121.6, 941.1) | 44.9 (36.8, 73.8) | 110.4 (106.1, 139) | 196.9 (194, 207.2) | 4.61 (2.00, 7.75) | 2.93 (1.04, 6.32) | 1.76 (0.62, 4.45) |
| DoA | 210.4 (74.7, 503.4) | 323.9 (111.9, 821.6) | 347.0 (121.6, 941.6) | 125.4 (81.8, 191.7) | 359.5 (233.4, 563.7) | 673.8 (436.1, 1056.4) | 1.72 (0.60, 3.74) | 0.91 (0.30, 2.33) | 0.52 (0.17, 1.46) |
| DaR | 210.2 (74.7, 503) | 323.7 (111.8, 821.5) | 346.9 (121.6, 941.1) | 46.2 (37.4, 77.9) | 111.7 (106.8, 145.7) | 198.1 (194.6, 211.2) | 4.49 (1.99, 7.44) | 2.89 (1.04, 6.10) | 1.74 (0.62, 4.42) |

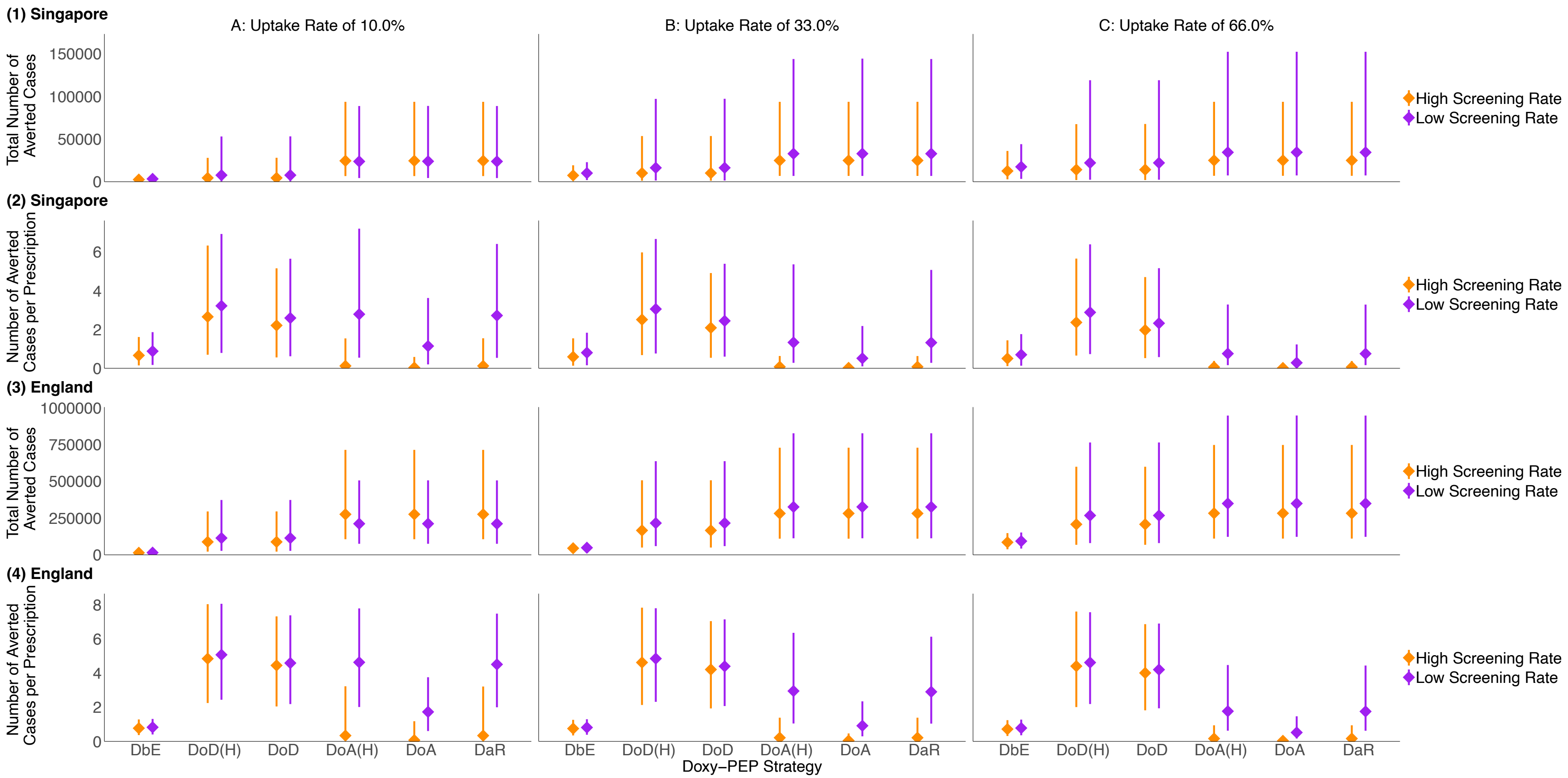


Fig. 9: From left to right, the assumed uptake rates are 0.10, 0.33, and 0.66. Each point-range plot displays the median estimate (central diamond) and the 95% credible interval (lines), under the assumption of suboptimal adherence that reduces protection to 33.0% of the baseline efficacy (i.e., $\zeta$ = 33.0%). The main scenario assumes stabilization of inferred time-varying behavioural trends, with all males not engaging in sex with females classified as MSM. Panels (1) and (3) show the total number of averted diagnosed cases comparing low vs. high asymptomatic screening rates across doxy-PEP strategies for Singapore and England, respectively. Panels (2) and (4) show the number of averted diagnosed cases per prescription, comparing low vs. high asymptomatic screening rates across doxy-PEP strategies for Singapore and England, respectively.

Next, we compare the results of the strategy under different adherence behavioural patterns to investigate the effectiveness of doxy-PEP strategies.

Table 14: Singapore: estimated impact of doxy-PEP strategies among MSM with a high adherence behavioural pattern (i.e., $u = 0.5$), where protection is reduced to 33.0% of the baseline efficacy for suboptimal adherence stratum (i.e., $\zeta = 33.0\%$), assuming stabilization of inferred time-varying behavioural trends in the main scenario (i.e., assuming all males not engaging in sex with females are classified as MSM). Results are presented as median (95% credible interval), assuming a high asymptomatic screening rate (taken from our calibration, aligned with CDC-recommended levels [18]).

| | Total number of averted cases, thousands: Uptake rate | | | Number of doxy-PEP prescriptions, thousands: Uptake rate | | | Number of averted cases per prescription: Uptake rate | | |
|---|---|---|---|---|---|---|---|---|---|
| | 10.0% | 33.0% | 66.0% | 10.0% | 33.0% | 66.0% | 10.0% | 33.0% | 66.0% |
| DbE | 4.0 (0.9, 10.2) | 10.9 (2.4, 31) | 16.7 (3.7, 54.5) | 3.8 (3.8, 3.8) | 12.5 (12.5, 12.5) | 25.0 (25.0, 25.0) | 1.06 (0.23, 2.70) | 0.88 (0.19, 2.48) | 0.67 (0.15, 2.18) |
| DoD(H) | 6.4 (0.6, 38) | 13.1 (1.7, 64.1) | 16.7 (2.7, 75.8) | 1.5 (0.5, 4) | 3.2 (1.5, 7.2) | 4.3 (2.3, 8.9) | 4.27 (1.13, 10.37) | 4.00 (1.09, 9.90) | 3.77 (1.04, 9.53) |
| DoD | 6.4 (0.6, 38) | 13.1 (1.7, 64.2) | 16.7 (2.7, 76) | 1.8 (0.7, 5.4) | 3.9 (1.8, 9.4) | 5.2 (2.7, 11.8) | 3.56 (0.90, 8.45) | 3.34 (0.87, 8.15) | 3.15 (0.83, 7.88) |
| DoA(H) | 24.5 (6.8, 93.1) | 24.8 (6.9, 93.1) | 24.8 (6.9, 93.1) | 162.7 (15, 261.8) | 231.4 (42.2, 510.1) | 313.5 (70, 859.9) | 0.17 (0.04, 1.72) | 0.11 (0.02, 0.72) | 0.08 (0.01, 0.52) |
| DoA | 24.5 (6.8, 93.1) | 24.8 (6.9, 93.1) | 24.8 (6.9, 93.1) | 616.3 (41.3, 1009.6) | 900.9 (122.7, 1706.1) | 1136.4 (216.3, 2661.5) | 0.05 (0.01, 0.64) | 0.03 (0.01, 0.26) | 0.02 (0.00, 0.16) |
| DaR | 24.5 (6.8, 93.1) | 24.8 (6.9, 93.1) | 24.8 (6.9, 93.1) | 162.7 (15.1, 261.8) | 231.4 (42.3, 510.1) | 313.5 (70.2, 860) | 0.17 (0.04, 1.72) | 0.11 (0.02, 0.72) | 0.08 (0.01, 0.52) |

Table 15: Singapore: estimated impact of doxy-PEP strategies among MSM with a low adherence behavioural pattern (i.e., $u = 2$), where protection is reduced to 33.0% of the baseline efficacy for suboptimal adherence stratum (i.e., $\zeta = 33.0\%$), assuming stabilization of inferred time-varying behavioural trends in the main scenario (i.e., assuming all males not engaging in sex with females are classified as MSM). Results are presented as median (95% credible interval), assuming a high asymptomatic screening rate (taken from our calibration, aligned with CDC-recommended levels [18]).

| | Total number of averted cases, thousands: Uptake rate | | | Number of doxy-PEP prescriptions, thousands: Uptake rate | | | Number of averted cases per prescription: Uptake rate | | |
|---|---|---|---|---|---|---|---|---|---|
| | 10.0% | 33.0% | 66.0% | 10.0% | 33.0% | 66.0% | 10.0% | 33.0% | 66.0% |
| DbE | 1.4 (0.3, 3.3) | 4.3 (1, 10.7) | 8.0 (1.8, 20.7) | 3.8 (3.8, 3.8) | 12.5 (12.5, 12.5) | 25.0 (25.0, 25.0) | 0.37 (0.09, 0.88) | 0.35 (0.08, 0.86) | 0.32 (0.07, 0.83) |
| DoD(H) | 2.7 (0.2, 18.1) | 6.9 (0.7, 40.1) | 10.6 (1.3, 54.9) | 1.8 (0.6, 5.5) | 4.8 (1.7, 12.7) | 7.8 (3.2, 18.7) | 1.50 (0.39, 3.53) | 1.43 (0.38, 3.35) | 1.37 (0.37, 3.20) |
| DoD | 2.7 (0.2, 18.1) | 6.9 (0.7, 40.3) | 10.7 (1.3, 55.1) | 2.2 (0.7, 7.4) | 5.8 (2.1, 17.1) | 9.3 (3.8, 24.3) | 1.24 (0.31, 2.87) | 1.18 (0.31, 2.76) | 1.13 (0.30, 2.64) |
| DoA(H) | 24.0 (6.4, 90.9) | 24.6 (6.8, 93.1) | 24.8 (6.9, 93.1) | 350.9 (16.2, 630.2) | 571.1 (49.3, 944.6) | 709.9 (91.7, 1311.4) | 0.09 (0.02, 1.20) | 0.05 (0.01, 0.53) | 0.04 (0.01, 0.32) |
| DoA | 24.0 (6.4, 91.1) | 24.7 (6.8, 93.1) | 24.8 (6.9, 93.1) | 1117.3 (43.9, 2457.2) | 2108.5 (136, 3644.6) | 2733.1 (259.5, 4746.9) | 0.03 (0.00, 0.47) | 0.01 (0.00, 0.20) | 0.01 (0.00, 0.11) |
| DaR | 24.0 (6.4, 90.9) | 24.6 (6.8, 93.1) | 24.8 (6.9, 93.1) | 350.9 (16.5, 630.2) | 571.2 (49.5, 944.7) | 709.9 (91.9, 1311.4) | 0.09 (0.02, 1.18) | 0.05 (0.01, 0.52) | 0.04 (0.01, 0.32) |

Table 16: England: estimated impact of doxy-PEP strategies among MSM with a high adherence behavioural pattern (i.e., $u = 0.5$), where protection is reduced to 33.0% of the baseline efficacy for suboptimal adherence stratum (i.e., $\zeta = 33.0\%$), assuming stabilization of inferred time-varying behavioural trends. Results are presented as median (95% credible interval), assuming a high asymptomatic screening rate (taken from our calibration, aligned with CDC-recommended levels [18]).

| | Total number of averted cases, thousands: Uptake rate | | | Number of doxy-PEP prescriptions, thousands: Uptake rate | | | Number of averted cases per prescription: Uptake rate | | |
|---|---|---|---|---|---|---|---|---|---|
| | 10.0% | 33.0% | 66.0% | 10.0% | 33.0% | 66.0% | 10.0% | 33.0% | 66.0% |
| DbE | 23.3<br>(10.8, 39.1) | 72.6<br>(31.9, 126.3) | 130.8<br>(53.2, 241.5) | 18.0<br>(18.0, 18.0) | 59.4<br>(59.4, 59.4) | 118.8<br>(118.8, 118.8) | 1.29<br>(0.60, 2.17) | 1.22<br>(0.54, 2.13) | 1.10<br>(0.45, 2.03) |
| DoD(H) | 121.2<br>(32.6, 395.4) | 199.6<br>(63.3, 573.8) | 232.6<br>(80.7, 662.2) | 14.7<br>(7.0, 33.3) | 25.1<br>(13.3, 55.1) | 30.9<br>(17.6, 67.6) | 8.13<br>(3.70, 13.92) | 7.77<br>(3.45, 13.61) | 7.39<br>(3.23, 13.22) |
| DoD | 121.2<br>(32.6, 395.4) | 199.6<br>(63.3, 573.8) | 232.6<br>(80.7, 662.3) | 16.0<br>(7.7, 38) | 27.7<br>(15.3, 61.2) | 34.0<br>(19.2, 74.3) | 7.44<br>(3.35, 12.58) | 7.06<br>(3.13, 12.20) | 6.75<br>(2.92, 11.90) |
| DoA(H) | 276.1<br>(106.3, 724) | 280.0<br>(109.6, 738.5) | 280.9<br>(109.8, 746.3) | 594.3<br>(78.3, 944.5) | 835.9<br>(187.9, 1761.7) | 1101.5<br>(301.7, 2906.1) | 0.49<br>(0.16, 3.86) | 0.32<br>(0.09, 1.79) | 0.24<br>(0.06, 1.33) |
| DoA | 276.2<br>(106.3, 724) | 280.0<br>(109.6, 738.5) | 280.9<br>(109.8, 746.3) | 3018.4<br>(232.4, 5064.7) | 4514.5<br>(669.8, 8208.7) | 5570.3<br>(1155.5, 12465.5) | 0.10<br>(0.03, 1.32) | 0.06<br>(0.02, 0.50) | 0.05<br>(0.01, 0.30) |
| DaR | 276.1<br>(106.3, 724) | 280.0<br>(109.6, 738.5) | 280.9<br>(109.8, 746.3) | 594.4<br>(78.6, 944.6) | 836.1<br>(188.3, 1761.9) | 1101.9<br>(302.8, 2906.4) | 0.49<br>(0.16, 3.86) | 0.32<br>(0.09, 1.78) | 0.24<br>(0.06, 1.32) |

Table 17: England: estimated impact of doxy-PEP strategies among MSM with a low adherence behavioural pattern (i.e., $u$ = 2), where protection is reduced to 33.0% of the baseline efficacy for suboptimal adherence stratum (i.e., $\zeta$ = 33.0%), assuming stabilization of inferred time-varying behavioural trends. Results are presented as median (95% credible interval), assuming a high asymptomatic screening rate (taken from our calibration, aligned with CDC-recommended levels [18]).

| | Total number of averted cases, thousands: Uptake rate | | | Number of doxy-PEP prescriptions, thousands: Uptake rate | | | Number of averted cases per prescription: Uptake rate | | |
|---|---|---|---|---|---|---|---|---|---|
| | 10.0% | 33.0% | 66.0% | 10.0% | 33.0% | 66.0% | 10.0% | 33.0% | 66.0% |
| DbE | 7.6 (3.7, 12.5) | 24.6 (11.8, 40.9) | 48.2 (22.5, 80.7) | 18.0 (18.0, 18.0) | 59.4 (59.4, 59.4) | 118.8 (118.8, 118.8) | 0.42 (0.20, 0.69) | 0.41 (0.20, 0.69) | 0.41 (0.19, 0.68) |
| DoD(H) | 56.4 (13.2, 197.3) | 124.8 (34.3, 398.3) | 170.4 (51.4, 513.1) | 20.6 (9.0, 50.9) | 47.7 (22.7, 109.3) | 68.4 (34.4, 148) | 2.67 (1.25, 4.37) | 2.56 (1.20, 4.26) | 2.47 (1.15, 4.15) |
| DoD | 56.4 (13.2, 197.3) | 124.8 (34.4, 398.4) | 170.4 (51.4, 513.1) | 22.9 (10.1, 57.4) | 52.1 (25.0, 124) | 73.8 (38.3, 168.4) | 2.44 (1.14, 3.96) | 2.34 (1.09, 3.86) | 2.24 (1.04, 3.73) |
| DoA(H) | 271.9 (100.5, 705.7) | 278.3 (107.5, 724) | 280.3 (109.7, 724) | 1215.4 (87.3, 2315.1) | 2068.7 (256.9, 3426.5) | 2586.3 (421.0, 4648.1) | 0.24 (0.07, 2.37) | 0.14 (0.04, 1.20) | 0.11 (0.03, 0.71) |
| DoA | 271.9 (100.5, 705.7) | 278.3 (107.5, 724) | 280.3 (109.7, 724) | 5128.6 (258.5, 12257.9) | 10263.2 (764.3, 18276.3) | 13575.6 (1437.4, 23404.6) | 0.06 (0.01, 0.81) | 0.03 (0.01, 0.41) | 0.02 (0.01, 0.24) |
| DaR | 271.9 (100.5, 705.7) | 278.3 (107.5, 724) | 280.3 (109.7, 724) | 1215.4 (89.7, 2315.1) | 2068.8 (257.7, 3426.7) | 2586.6 (422.0, 4648.5) | 0.24 (0.07, 2.33) | 0.14 (0.04, 1.20) | 0.11 (0.03, 0.71) |

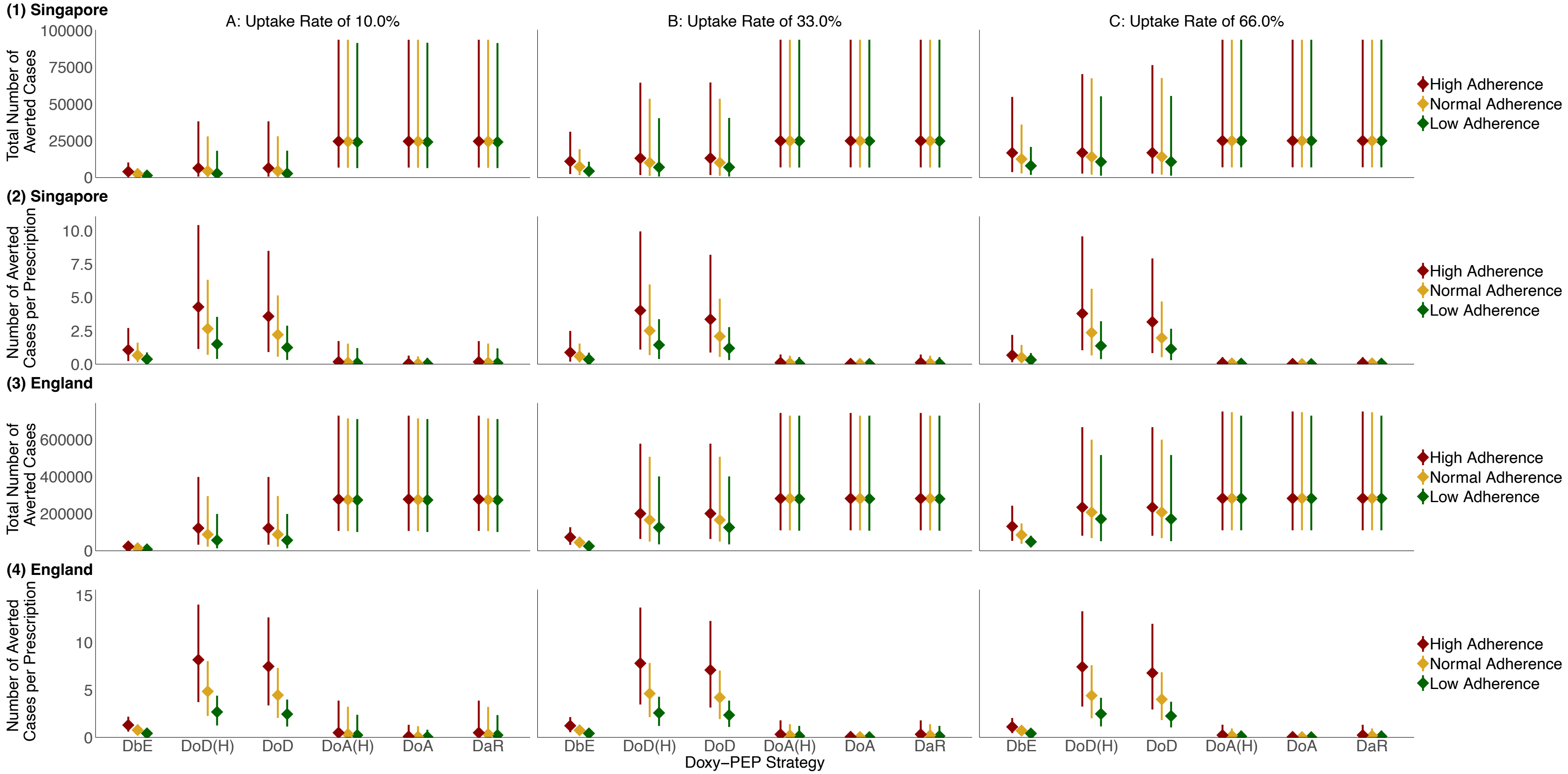


Fig. 10: From left to right, the assumed uptake rates are 0.10, 0.33, and 0.66. Each point-range plot displays the median estimate (central diamond) and the 95% credible interval (lines), under the assumption of suboptimal adherence that reduces protection to 33.0% of the baseline efficacy (i.e., $\zeta$ = 33.0%). The Singapore main scenario assumes stabilization of inferred time-varying behavioural trends, with all males not engaging in sex with females classified as MSM. Panels (1) and (3) show the total number of averted diagnosed cases comparing low vs. normal vs. high adherence behavioural patterns across doxy-PEP strategies for Singapore and England, respectively. Panels (2) and (4) show the number of averted diagnosed cases per prescription, comparing low vs. normal vs. high adherence behavioural patterns across doxy-PEP strategies for Singapore and England, respectively.

3. Results of Singapore (main scenario) and England

This subsection presents results under the following assumption: (1) stabilization of inferred time-varying behavioural trends. The outcomes include the total number of averted syphilis cases, total number of doxy-PEP prescriptions, number of averted cases per prescription. These results are stratified by varying levels of uptake and $\zeta$.

Table 18: Singapore: estimated impact and efficiency of doxy-PEP strategies among MSM with a normal adherence behavioural pattern (i.e., $u = 1$), where protection is reduced to 33.0% of the baseline efficacy for suboptimal adherence stratum (i.e., $\zeta = 10.0\%$), assuming the inferred time-varying behavioural trends persist through 2040 in the main scenario (i.e., assuming all males not engaging in sex with females are classified as MSM). Results are presented as median (95% credible interval), assuming a high asymptomatic screening rate (taken from our calibration, aligned with CDC-recommended levels [18]).

| | Total number of averted cases, thousands: Uptake ratio | | | Number of doxy-PEP prescription, thousands: Uptake ratio | | | Number of averted cases per prescription: Uptake ratio | | |
|---|---|---|---|---|---|---|---|---|---|
| | 10.0% | 33.0% | 66.0% | 10.0% | 33.0% | 66.0% | 10.0% | 33.0% | 66.0% |
| DbE | 2.3 (0.5, 5.6) | 6.8 (1.5, 17.7) | 11.8 (2.7, 33.4) | 3.8 (3.8, 3.8) | 12.5 (12.5, 12.5) | 25.0 (25.0, 25.0) | 0.61 (0.14, 1.48) | 0.55 (0.12, 1.42) | 0.47 (0.10, 1.33) |
| DoD(H) | 4.1 (0.4, 26.4) | 9.6 (1.1, 51.6) | 13.6 (1.8, 65.5) | 1.7 (0.6, 4.8) | 4.1 (1.6, 10.3) | 6.1 (2.9, 13.7) | 2.44 (0.64, 5.81) | 2.31 (0.63, 5.49) | 2.18 (0.60, 5.21) |
| DoD | 4.1 (0.4, 26.5) | 9.6 (1.1, 51.6) | 13.6 (1.8, 65.7) | 2.1 (0.7, 6.5) | 4.9 (2.0, 13.3) | 7.4 (3.4, 17.8) | 2.03 (0.51, 4.73) | 1.92 (0.50, 4.52) | 1.82 (0.48, 4.32) |
| DoA(H) | 24.2 (6.6, 93.1) | 24.7 (6.8, 93.1) | 24.8 (6.9, 93.1) | 244.9 (15.6, 396.2) | 358.7 (46.3, 659.4) | 454.4 (82.1, 1012.7) | 0.12 (0.03, 1.50) | 0.08 (0.02, 0.63) | 0.06 (0.01, 0.37) |
| DoA | 24.2 (6.6, 93.1) | 24.7 (6.8, 93.1) | 24.8 (6.9, 93.1) | 853.8 (42.4, 1568.7) | 1390.7 (130.5, 2390.9) | 1732.9 (241.1, 3376.4) | 0.03 (0.01, 0.57) | 0.02 (0.00, 0.23) | 0.02 (0.00, 0.13) |
| DaR | 24.2 (6.6, 93.1) | 24.7 (6.8, 93.1) | 24.8 (6.9, 93.1) | 244.9 (15.8, 396.2) | 358.7 (46.4, 659.4) | 454.4 (82.3, 1012.8) | 0.12 (0.03, 1.50) | 0.08 (0.02, 0.61) | 0.06 (0.01, 0.37) |

Table 19: Singapore: estimated impact and efficiency of doxy-PEP strategies among MSM with a normal adherence behavioural pattern (i.e., $u = 1$), where protection is reduced to 33.0% of the baseline efficacy for suboptimal adherence stratum (i.e., $\zeta = 66.0\%$), assuming the inferred time-varying behavioural trends persist through 2040 in the main scenario (i.e., assuming all males not engaging in sex with females are classified as MSM). Results are presented as median (95% credible interval), assuming a high asymptomatic screening rate (taken from our calibration, aligned with CDC-recommended levels [18]).

| | Total number of averted cases, thousands: Uptake ratio | | | Number of doxy-PEP prescription, thousands: Uptake ratio | | | Number of averted cases per prescription: Uptake ratio | | |
|---|---|---|---|---|---|---|---|---|---|
| | 10.0% | 33.0% | 66.0% | 10.0% | 33.0% | 66.0% | 10.0% | 33.0% | 66.0% |
| DbE | 2.8 (0.6, 6.8) | 8.0 (1.8, 21.2) | 13.4 (3.0, 39.2) | 3.8 (3.8, 3.8) | 12.5 (12.5, 12.5) | 25.0 (25.0, 25.0) | 0.73 (0.16, 1.78) | 0.64 (0.14, 1.70) | 0.53 (0.12, 1.57) |
| DoD(H) | 4.8 (0.4, 29.8) | 10.7 (1.3, 55.4) | 14.6 (2.1, 68.9) | 1.6 (0.5, 4.6) | 3.8 (1.6, 9.3) | 5.5 (2.7, 12.1) | 2.92 (0.77, 6.98) | 2.75 (0.75, 6.58) | 2.59 (0.72, 6.22) |
| DoD | 4.8 (0.4, 29.9) | 10.7 (1.3, 55.5) | 14.6 (2.1, 69) | 2.0 (0.7, 6.1) | 4.6 (1.9, 12) | 6.7 (3.2, 15.8) | 2.44 (0.62, 5.69) | 2.30 (0.60, 5.41) | 2.16 (0.58, 5.18) |
| DoA(H) | 24.5 (6.7, 93.1) | 24.7 (6.8, 93.1) | 24.8 (6.9, 93.1) | 244.9 (15.5, 396.2) | 358.7 (46.2, 659.4) | 454.4 (82.1, 1012.7) | 0.12 (0.03, 1.56) | 0.08 (0.02, 0.63) | 0.06 (0.01, 0.38) |
| DoA | 24.5 (6.7, 93.1) | 24.8 (6.8, 93.1) | 24.8 (6.9, 93.1) | 853.8 (42.2, 1568.7) | 1390.7 (130.5, 2390.9) | 1732.9 (241.1, 3376.4) | 0.03 (0.01, 0.60) | 0.02 (0.00, 0.23) | 0.02 (0.00, 0.13) |
| DaR | 24.5 (6.7, 93.1) | 24.7 (6.8, 93.1) | 24.8 (6.9, 93.1) | 244.9 (15.7, 396.2) | 358.7 (46.4, 659.4) | 454.4 (82.3, 1012.8) | 0.12 (0.03, 1.51) | 0.08 (0.02, 0.63) | 0.06 (0.01, 0.38) |

Table 20: England: estimated impact and efficiency of doxy-PEP strategies among MSM with a normal adherence behavioural pattern (i.e., $u = 1$), where protection is reduced to 33.0% of the baseline efficacy for suboptimal adherence stratum (i.e., $\zeta = 10.0\%$), assuming the inferred time-varying behavioural trends persist through 2040 in the main scenario (i.e., assuming all males not engaging in sex with females are classified as MSM). Results are presented as median (95% credible interval), assuming a high asymptomatic screening rate (taken from our calibration, aligned with CDC-recommended levels [18]).

| | Total number of averted cases, thousands: Uptake ratio | | | Number of doxy-PEP prescription, thousands: Uptake ratio | | | Number of averted cases per prescription: Uptake ratio | | |
|---|---|---|---|---|---|---|---|---|---|
| | 10.0% | 33.0% | 66.0% | 10.0% | 33.0% | 66.0% | 10.0% | 33.0% | 66.0% |
| DbE | 12.7<br>(6.1, 21.2) | 41.1<br>(19.1, 68.8) | 78.8<br>(35.3, 135.3) | 18.0<br>(18.0, 18.0) | 59.4<br>(59.4, 59.4) | 118.8<br>(118.8, 118.8) | 0.71<br>(0.34, 1.18) | 0.69<br>(0.32, 1.16) | 0.66<br>(0.30, 1.14) |
| DoD(H) | 82.7<br>(20.4, 276.7) | 159.6<br>(47.2, 491.7) | 202.0<br>(65.5, 581.6) | 18.3<br>(8.2, 43.6) | 37.3<br>(18.5, 81.3) | 49.1<br>(25.5, 106.2) | 4.45<br>(2.07, 7.37) | 4.24<br>(1.96, 7.18) | 4.05<br>(1.86, 6.98) |
| DoD | 82.7<br>(20.4, 276.8) | 159.6<br>(47.2, 491.7) | 202.0<br>(65.5, 581.6) | 20.2<br>(9.1, 49.8) | 40.2<br>(20.6, 92.4) | 53.9<br>(29.2, 117.9) | 4.08<br>(1.88, 6.71) | 3.87<br>(1.78, 6.45) | 3.69<br>(1.68, 6.30) |
| DoA(H) | 273.9<br>(105.3, 709.2) | 279.7<br>(108.9, 724) | 280.3<br>(109.7, 741.1) | 874.7<br>(82.1, 1452.1) | 1308.0<br>(219.0, 2336.9) | 1636.4<br>(361.8, 3497.5) | 0.34<br>(0.10, 3.16) | 0.21<br>(0.07, 1.38) | 0.16<br>(0.05, 0.94) |
| DoA | 273.9<br>(105.4, 709.2) | 279.7<br>(108.9, 724) | 280.3<br>(109.7, 741.1) | 4059.1<br>(248, 7909.2) | 6919.2<br>(724.2, 11785.3) | 8643.8<br>(1311.3, 16245.6) | 0.07<br>(0.02, 1.13) | 0.04<br>(0.01, 0.46) | 0.03<br>(0.01, 0.26) |
| DaR | 273.9<br>(105.3, 709.2) | 279.7<br>(108.9, 724) | 280.3<br>(109.7, 741.1) | 874.7<br>(82.5, 1452.2) | 1308.1<br>(219.8, 2337.1) | 1636.9<br>(362.4, 3497.9) | 0.34<br>(0.10, 3.14) | 0.21<br>(0.07, 1.38) | 0.16<br>(0.05, 0.94) |

Table 21: England: estimated impact and efficiency of doxy-PEP strategies among MSM with a normal adherence behavioural pattern (i.e., $u = 1$), where protection is reduced to 33.0% of the baseline efficacy for suboptimal adherence stratum (i.e., $\zeta = 66.0\%$), assuming the inferred time-varying behavioural trends persist through 2040 in the main scenario (i.e., assuming all males not engaging in sex with females are classified as MSM). Results are presented as median (95% credible interval), assuming a high asymptomatic screening rate (taken from our calibration, aligned with CDC-recommended levels [18]).

| | Total number of averted cases, thousands: Uptake ratio | | | Number of doxy-PEP prescription, thousands: Uptake ratio | | | Number of averted cases per prescription: Uptake ratio | | |
|---|---|---|---|---|---|---|---|---|---|
| | 10.0% | 33.0% | 66.0% | 10.0% | 33.0% | 66.0% | 10.0% | 33.0% | 66.0% |
| DbE | 15.4 (7.4, 25.7) | 49.4 (22.8, 83.3) | 93.4 (40.6, 162.8) | 18.0 (18.0, 18.0) | 59.4 (59.4, 59.4) | 118.8 (118.8, 118.8) | 0.86 (0.41, 1.43) | 0.83 (0.38, 1.40) | 0.79 (0.34, 1.37) |
| DoD(H) | 93.9 (23.9, 316.9) | 172.9 (52.1, 519) | 211.3 (70.7, 613.2) | 17.3 (7.9, 40.7) | 33.4 (16.9, 72.4) | 42.9 (22.7, 94) | 5.36 (2.49, 8.91) | 5.11 (2.35, 8.68) | 4.86 (2.21, 8.40) |
| DoD | 93.9 (23.9, 317) | 172.9 (52.1, 519.1) | 211.4 (70.7, 613.2) | 18.9 (8.7, 46.9) | 36.1 (18.8, 82.1) | 47.4 (25.8, 104) | 4.93 (2.26, 8.13) | 4.65 (2.12, 7.80) | 4.42 (2.00, 7.62) |
| DoA(H) | 274.6 (105.6, 717.5) | 279.8 (109.5, 724) | 280.9 (109.8, 744) | 874.7 (81.8, 1452.1) | 1308.0 (212.7, 2336.9) | 1636.4 (360.7, 3497.5) | 0.34 (0.10, 3.27) | 0.21 (0.07, 1.38) | 0.16 (0.05, 0.94) |
| DoA | 274.6 (105.6, 717.6) | 279.9 (109.5, 724) | 280.9 (109.8, 744) | 4059.1 (242.6, 7909.2) | 6919.2 (723.9, 11785.3) | 8643.8 (1311.1, 16245.6) | 0.07 (0.02, 1.23) | 0.04 (0.01, 0.47) | 0.03 (0.01, 0.26) |
| DaR | 274.6 (105.6, 717.5) | 279.8 (109.5, 724) | 280.9 (109.8, 744) | 874.7 (82.2, 1452.2) | 1308.1 (213.3, 2337.1) | 1636.9 (361.2, 3497.9) | 0.34 (0.10, 3.26) | 0.21 (0.07, 1.38) | 0.16 (0.05, 0.94) |

4. Results of Singapore (main scenario) and England

This subsection presents results under the following assumptions: (1) protection is reduced to 33.0% of the baseline efficacy for suboptimal adherence stratum (i.e., $\zeta$ = 33.0%) and (2) the inferred time-varying behavioural trends persist through 2040. The outcomes include the total number of averted syphilis cases, total number of doxy-PEP prescriptions, number of averted cases per prescription. These results are stratified by varying levels of uptake and the inferred time-varying behavioural trends.

Table 22: Singapore: estimated impact and efficiency of doxy-PEP strategies among MSM with a normal adherence behavioural pattern (i.e., $u = 1$), where protection is reduced to 33.0% of the baseline efficacy for suboptimal adherence stratum (i.e., $\zeta = 33.0\%$), assuming the inferred time-varying behavioural trends persist through 2040 in the main scenario (i.e., assuming all males not engaging in sex with females are classified as MSM). Results are presented as median (95% credible interval), assuming a high asymptomatic screening rate (taken from our calibration, aligned with CDC-recommended levels [18]).

| | Total number of averted cases, thousands: Uptake ratio | | | Number of doxy-PEP prescription, thousands: Uptake ratio | | | Number of averted cases per prescription: Uptake ratio | | |
|---|---|---|---|---|---|---|---|---|---|
| | 10.0% | 33.0% | 66.0% | 10.0% | 33.0% | 66.0% | 10.0% | 33.0% | 66.0% |
| DbE | 4.1 (0.7, 7.6) | 12.8 (1.9, 25.1) | 23.4 (3.2, 49.5) | 3.8 (3.8, 3.8) | 12.5 (12.5, 12.5) | 25.0 (25.0, 25.0) | 1.09 (0.17, 2.02) | 1.03 (0.15, 2.01) | 0.94 (0.13, 1.98) |
| DoD(H) | 12.0 (0.6, 63.9) | 24.5 (1.5, 119.4) | 32.2 (2.5, 146.6) | 3.0 (0.6, 9.2) | 6.6 (1.7, 17.8) | 9.0 (2.9, 22.9) | 3.78 (0.82, 7.79) | 3.61 (0.79, 7.55) | 3.44 (0.76, 7.28) |
| DoD | 12.0 (0.6, 64.3) | 24.5 (1.5, 119.6) | 32.2 (2.5, 146.8) | 3.7 (0.7, 12.3) | 7.8 (2.1, 23.1) | 10.8 (3.5, 29.6) | 3.14 (0.66, 6.20) | 3.03 (0.64, 6.03) | 2.90 (0.61, 5.83) |
| DoA(H) | 48.9 (7.3, 189.5) | 49.6 (7.5, 189.7) | 49.6 (7.6, 189.8) | 268.5 (20, 410) | 373.1 (58, 678.9) | 467.7 (100.1, 1047.2) | 0.22 (0.03, 2.79) | 0.14 (0.02, 1.11) | 0.11 (0.01, 0.76) |
| DoA | 48.9 (7.3, 189.5) | 49.6 (7.5, 189.7) | 49.6 (7.6, 189.8) | 981.4 (55.6, 1651.5) | 1482.7 (168.4, 2461.1) | 1806.0 (301.4, 3478.9) | 0.06 (0.01, 1.07) | 0.04 (0.00, 0.41) | 0.03 (0.00, 0.25) |
| DaR | 48.9 (7.3, 189.5) | 49.6 (7.5, 189.7) | 49.6 (7.6, 189.8) | 268.5 (20.2, 410) | 373.2 (58.1, 678.9) | 467.8 (100.2, 1047.2) | 0.22 (0.03, 2.78) | 0.14 (0.02, 1.11) | 0.11 (0.01, 0.73) |

Table 23: England: estimated impact and efficiency of doxy-PEP strategies among MSM with a normal adherence behavioural pattern (i.e., $u = 1$), where protection is reduced to 33.0% of the baseline efficacy for suboptimal adherence stratum (i.e., $\zeta = 33.0\%$), assuming the inferred time-varying behavioural trends persist through 2040. Results are presented as median (95% credible interval), assuming a high asymptomatic screening rate (taken from our calibration, aligned with CDC-recommended levels [18]).

| | Total number of averted cases, thousands: Uptake ratio | | | Number of doxy-PEP prescription, thousands: Uptake ratio | | | Number of averted cases per prescription: Uptake ratio | | |
|---|---|---|---|---|---|---|---|---|---|
| | 10.0% | 33.0% | 66.0% | 10.0% | 33.0% | 66.0% | 10.0% | 33.0% | 66.0% |
| DbE | 16.1 | 52.4 | 102.2 | 18.0 | 59.4 | 118.8 | 0.90 | 0.88 | 0.86 |
| | (7.2, 25.5) | (22.4, 84) | (41.4, 166.4) | (18.0, 18.0) | (59.4, 59.4) | (118.8, 118.8) | (0.40, 1.42) | (0.38, 1.41) | (0.35, 1.40) |
| DoD(H) | 142.6 | 264.3 | 324.8 | 26.5 | 49.9 | 63.9 | 5.34 | 5.15 | 4.97 |
| | (24.6, 493.9) | (54.3, 876.8) | (73.2, 1025.8) | (8.5, 69.0) | (18.6, 122.9) | (25.7, 151.9) | (2.46, 8.56) | (2.32, 8.33) | (2.16, 8.12) |
| DoD | 142.7 | 264.3 | 324.9 | 29.2 | 55.0 | 69.9 | 4.88 | 4.68 | 4.50 |
| | (24.6, 494) | (54.3, 876.9) | (73.2, 1025.9) | (9.4, 78.5) | (20.7, 142) | (28.5, 174.4) | (2.22, 7.81) | (2.09, 7.57) | (1.97, 7.38) |
| DoA(H) | 425.4 | 431.4 | 433.4 | 983.4 | 1374.1 | 1692.4 | 0.47 | 0.31 | 0.24 |
| | (112.1, 1226.2) | (115.6, 1266.1) | (116.8, 1273.1) | (110.5, 1514.9) | (295.0, 2414.7) | (466.3, 3630.5) | (0.10, 4.08) | (0.07, 1.84) | (0.05, 1.37) |
| DoA | 425.4 | 431.4 | 433.5 | 4799.0 | 7498.4 | 9105.1 | 0.10 | 0.06 | 0.05 |
| | (112.1, 1226.2) | (115.7, 1266.1) | (116.8, 1273.1) | (339.0, 8436.6) | (983, 12208.7) | (1737.6, 16811.1) | (0.02, 1.42) | (0.01, 0.58) | (0.01, 0.33) |
| DaR | 425.4 | 431.4 | 433.4 | 983.4 | 1374.3 | 1692.3 | 0.47 | 0.31 | 0.24 |
| | (112.1, 1226.2) | (115.6, 1266.1) | (116.8, 1273.1) | (111.2, 1515.0) | (296.1, 2414.9) | (466.8, 3630.9) | (0.10, 4.04) | (0.07, 1.84) | (0.05, 1.37) |

5. Results of Singapore (upper bound scenario)

This subsection presents results under the following assumption: (1) protection is reduced to 33.0% of the baseline efficacy for suboptimal adherence stratum (i.e., $\zeta$ = 33.0%). The outcomes include the total number of averted syphilis cases, total number of doxy-PEP prescriptions, number of averted cases per prescription. These results are stratified by varying levels of uptake and the inferred time-varying behavioural trends.

Table 24: Singapore: estimated impact and efficiency of doxy-PEP strategies among MSM with a normal adherence behavioural pattern (i.e., $u = 1$), where protection is reduced to 33.0% of the baseline efficacy for suboptimal adherence stratum (i.e., $\zeta = 33.0\%$), assuming stabilization of inferred time-varying behavioural trends in the upper bound scenario (i.e., assuming all males are classified as MSM). Results are presented as median (95% credible interval), assuming a high asymptomatic screening rate (taken from our calibration, aligned with CDC-recommended levels [18]).

| | Total number of averted cases, thousands: Uptake ratio | | | Number of doxy-PEP prescription, thousands: Uptake ratio | | | Number of averted cases per prescription: Uptake ratio | | |
|---|---|---|---|---|---|---|---|---|---|
| | 10.0% | 33.0% | 66.0% | 10.0% | 33.0% | 66.0% | 10.0% | 33.0% | 66.0% |
| DbE | 2.7 (1.0, 6.3) | 8.0 (2.9, 19.4) | 13.6 (4.9, 36.7) | 3.8 (3.8, 3.8) | 12.5 (12.5, 12.5) | 25.0 (25.0, 25.0) | 0.71 (0.26, 1.65) | 0.64 (0.23, 1.55) | 0.55 (0.20, 1.47) |
| DoD(H) | 6.2 (1.2, 32.3) | 13.3 (3.3, 59.8) | 17.7 (5.2, 71.7) | 2.0 (1.0, 5.2) | 4.5 (2.6, 10.6) | 6.5 (4.1, 13.9) | 3.18 (1.19, 6.58) | 2.99 (1.14, 6.28) | 2.82 (1.08, 6.00) |
| DoD | 6.2 (1.2, 32.5) | 13.3 (3.3, 59.8) | 17.7 (5.2, 71.7) | 2.3 (1.2, 6.8) | 5.4 (3.1, 13.6) | 7.8 (4.9, 18) | 2.60 (0.98, 5.47) | 2.46 (0.94, 5.20) | 2.31 (0.90, 4.96) |
| DoA(H) | 28.4 (12.4, 95.5) | 28.6 (12.7, 96.3) | 28.7 (12.7, 96.5) | 246.5 (21.5, 389.7) | 360.6 (61.8, 641.8) | 457.4 (105.9, 978.2) | 0.14 (0.05, 1.09) | 0.09 (0.03, 0.47) | 0.07 (0.02, 0.35) |
| DoA | 28.4 (12.4, 95.6) | 28.7 (12.7, 96.3) | 28.8 (12.7, 96.5) | 850.1 (55.5, 1570.9) | 1388.4 (167.4, 2390.4) | 1734.7 (294.3, 3382.3) | 0.04 (0.01, 0.45) | 0.02 (0.01, 0.17) | 0.02 (0.01, 0.11) |
| DaR | 28.4 (12.4, 95.5) | 28.6 (12.7, 96.3) | 28.7 (12.7, 96.5) | 246.6 (21.6, 389.7) | 360.7 (62.0, 641.8) | 457.5 (106.1, 978.3) | 0.14 (0.05, 1.09) | 0.09 (0.03, 0.47) | 0.07 (0.02, 0.35) |

Table 25: Singapore: estimated impact and efficiency of doxy-PEP strategies among MSM with a normal adherence behavioural pattern (i.e., $u = 1$), where protection is reduced to 33.0% of the baseline efficacy for suboptimal adherence stratum (i.e., $\zeta = 33.0\%$), assuming the inferred time-varying behavioural trends persist through 2040 in the upper bound scenario (i.e., assuming all males are classified as MSM). Results are presented as median (95% credible interval), assuming a high asymptomatic screening rate (taken from our calibration, aligned with CDC-recommended levels [18]).

| | Total number of averted cases, thousands: Uptake ratio | | | Number of doxy-PEP prescription thousands: Uptake ratio | | | Number of averted cases per prescription: Uptake ratio | | |
|---|---|---|---|---|---|---|---|---|---|
| | 10.0% | 33.0% | 66.0% | 10.0% | 33.0% | 66.0% | 10.0% | 33.0% | 66.0% |
| DbE | 3.7 (1.0, 7.5) | 11.0 (3.1, 24.3) | 19.7 (5.3, 47.8) | 3.8 (3.8, 3.8) | 12.5 (12.5, 12.5) | 25.0 (25.0, 25.0) | 0.96 (0.28, 1.97) | 0.88 (0.25, 1.94) | 0.79 (0.21, 1.91) |
| DoD(H) | 10.2 (1.3, 62.5) | 20.8 (3.5, 110.4) | 27.4 (5.5, 131.5) | 2.7 (1.0, 8.8) | 5.8 (2.6, 16.6) | 8.2 (4.1, 21.1) | 3.80 (1.30, 7.80) | 3.61 (1.25, 7.54) | 3.42 (1.18, 7.32) |
| DoD | 10.2 (1.3, 62.5) | 20.9 (3.5, 110.4) | 27.4 (5.5, 132.1) | 3.2 (1.2, 11.2) | 7.0 (3.3, 21.7) | 9.8 (5.0, 27.8) | 3.16 (1.04, 6.30) | 3.00 (0.99, 6.07) | 2.83 (0.95, 5.88) |
| DoA(H) | 41.0 (12.8, 170.8) | 41.2 (13.2, 173.4) | 41.2 (13.2, 173.4) | 270.6 (27.6, 403.7) | 375.3 (76.9, 660.7) | 470.9 (126.9, 1011.3) | 0.18 (0.05, 1.65) | 0.12 (0.03, 0.76) | 0.09 (0.02, 0.56) |
| DoA | 41.0 (12.8, 171.2) | 41.2 (13.2, 173.4) | 41.2 (13.3, 173.4) | 975.9 (74.7, 1652.5) | 1479.2 (215.4, 2462.5) | 1804.3 (370.2, 3486.6) | 0.05 (0.01, 0.61) | 0.03 (0.01, 0.25) | 0.02 (0.01, 0.17) |
| DaR | 41.0 (12.8, 170.8) | 41.2 (13.2, 173.4) | 41.2 (13.2, 173.4) | 270.6 (27.6, 403.7) | 375.4 (77.1, 660.7) | 470.9 (127.1, 1011.3) | 0.18 (0.05, 1.65) | 0.12 (0.03, 0.76) | 0.09 (0.02, 0.56) |

### 6. Results of Singapore (lower bound scenario)

This subsection presents results under the following assumption: (1) protection is reduced to 33.0% of the baseline efficacy for suboptimal adherence stratum (i.e., $\zeta$ = 33.0%). The outcomes include the total number of averted syphilis cases, total number of doxy-PEP prescriptions, number of averted cases per prescription. These results are stratified by varying levels of uptake and the inferred time-varying behavioural trends.

Table 26: Singapore: estimated impact and efficiency of doxy-PEP strategies among MSM with a normal adherence behavioural pattern (i.e., $u$ = 1), where protection is reduced to 33.0% of the baseline efficacy for suboptimal adherence stratum (i.e., $\zeta$ = 33.0%), assuming stabilization of inferred time-varying behavioural trends in the lower bound scenario (i.e., assuming equal risk of syphilis infection regardless of sexual preference). Results are presented as median (95% credible interval), assuming a high asymptomatic screening rate (taken from our calibration, aligned with CDC-recommended levels [18]).

| | Total number of averted cases, thousands: Uptake ratio | | | Number of doxy-PEP prescription, thousands: Uptake ratio | | | Number of averted cases per prescription: Uptake ratio | | |
|---|---|---|---|---|---|---|---|---|---|
| | 10.0% | 33.0% | 66.0% | 10.0% | 33.0% | 66.0% | 10.0% | 33.0% | 66.0% |
| DbE | 0.3<br>(0.1, 3.6) | 0.9<br>(0.2, 10.4) | 1.4<br>(0.3, 16.4) | 3.8<br>(3.8, 3.8) | 12.5<br>(12.5, 12.5) | 25.0<br>(25.0, 25.0) | 0.09<br>(0.02, 0.96) | 0.07<br>(0.02, 0.83) | 0.06<br>(0.01, 0.66) |
| DoD(H) | 0.1<br>(0, 4.4) | 0.2<br>(0, 9.9) | 0.4<br>(0, 13.8) | 0.2<br>(0.1, 1.6) | 0.6<br>(0.2, 4) | 1.1<br>(0.4, 5.8) | 0.36<br>(0.09, 2.87) | 0.36<br>(0.09, 2.78) | 0.35<br>(0.09, 2.68) |
| DoD | 0.1<br>(0, 4.4) | 0.2<br>(0, 9.9) | 0.4<br>(0, 13.8) | 0.2<br>(0.1, 2.1) | 0.7<br>(0.3, 5) | 1.4<br>(0.5, 7.5) | 0.30<br>(0.07, 2.31) | 0.30<br>(0.07, 2.19) | 0.29<br>(0.07, 2.09) |
| DoA(H) | 2.4<br>(0.8, 25.3) | 2.5<br>(0.8, 25.4) | 2.5<br>(0.8, 25.4) | 251.1<br>(34.4, 383.1) | 365.8<br>(93.5, 628.7) | 467<br>(149.3, 955.7) | 0.01<br>(0.00, 0.21) | 0.01<br>(0.00, 0.10) | 0.01<br>(0.00, 0.08) |
| DoA | 2.4<br>(0.8, 25.3) | 2.5<br>(0.8, 25.4) | 2.5<br>(0.8, 25.4) | 898.3<br>(87.7, 1573.9) | 1437.4<br>(252.5, 2396.5) | 1795.8<br>(438.7, 3384.5) | 0.00<br>(0.00, 0.07) | 0.00<br>(0.00, 0.03) | 0.00<br>(0.00, 0.02) |
| DaR | 2.4<br>(0.8, 25.3) | 2.5<br>(0.8, 25.4) | 2.5<br>(0.8, 25.4) | 251.1<br>(34.5, 383.1) | 365.8<br>(93.6, 628.7) | 467.0<br>(149.3, 955.7) | 0.01<br>(0.00, 0.21) | 0.01<br>(0.00, 0.10) | 0.01<br>(0.00, 0.08) |

Table 27: Singapore: estimated impact and efficiency of doxy-PEP strategies among MSM with a normal adherence behavioural pattern (i.e., $u$ = 1), where protection is reduced to 33.0% of the baseline efficacy for suboptimal adherence stratum (i.e., $\zeta$ = 33.0%), assuming the inferred time-varying behavioural trends persist through 2040 in the lower bound scenario (i.e., assuming equal risk of syphilis infection regardless of sexual preference). Results are presented as median (95% credible interval), assuming a high asymptomatic screening rate (taken from our calibration, aligned with CDC-recommended levels [18]).

| | Total number of averted cases, thousands: Uptake ratio | | | Number of doxy-PEP prescription, thousands: Uptake ratio | | | Number of averted cases per prescription: Uptake ratio | | |
|---|---|---|---|---|---|---|---|---|---|
| | 10.0% | 33.0% | 66.0% | 10.0% | 33.0% | 66.0% | 10.0% | 33.0% | 66.0% |
| DbE | 0.7<br>(0.1, 6.6) | 2.0<br>(0.2, 21) | 3.0<br>(0.4, 39.6) | 3.8<br>(3.8, 3.8) | 12.5<br>(12.5, 12.5) | 25.0<br>(25.0, 25.0) | 0.19<br>(0.02, 1.74) | 0.16<br>(0.02, 1.68) | 0.12<br>(0.01, 1.59) |
| DoD(H) | 0.2<br>(0, 28) | 0.7<br>(0, 51.6) | 1.1<br>(0, 63.1) | 0.4<br>(0.1, 5) | 1.1<br>(0.2, 10.2) | 2.0<br>(0.4, 13.4) | 0.65<br>(0.09, 5.37) | 0.63<br>(0.09, 5.21) | 0.62<br>(0.09, 5.05) |
| DoD | 0.2<br>(0, 28) | 0.7<br>(0, 51.6) | 1.1<br>(0, 63.1) | 0.5<br>(0.1, 6.3) | 1.4<br>(0.3, 12.9) | 2.3<br>(0.5, 17.7) | 0.51<br>(0.07, 4.32) | 0.50<br>(0.07, 4.25) | 0.49<br>(0.07, 4.10) |
| DoA(H) | 4.7<br>(0.8, 84.9) | 4.8<br>(0.8, 88.2) | 4.8<br>(0.8, 88.3) | 274.2<br>(40.3, 397) | 379.4<br>(106.2, 647.1) | 479.8<br>(164.7, 987.7) | 0.02<br>(0.00, 0.61) | 0.01<br>(0.00, 0.34) | 0.01<br>(0.00, 0.24) |
| DoA | 4.7<br>(0.8, 84.9) | 4.8<br>(0.8, 88.2) | 4.8<br>(0.8, 88.3) | 1025.8<br>(117.1, 1655.9) | 1527.8<br>(325.8, 2466.8) | 1866.1<br>(536.4, 3487.9) | 0.01<br>(0.00, 0.21) | 0.00<br>(0.00, 0.10) | 0.00<br>(0.00, 0.07) |
| DaR | 4.7<br>(0.8, 84.9) | 4.8<br>(0.8, 88.2) | 4.8<br>(0.8, 88.3) | 274.2<br>(40.3, 397) | 3794.5<br>(106.2, 647.1) | 479.9<br>(164.7, 987.7) | 0.02<br>(0.00, 0.61) | 0.01<br>(0.00, 0.34) | 0.01<br>(0.00, 0.24) |

**Reference:**


[1] R. W. Peeling, D. Mabey, M. L. Kamb, X.-S. Chen, J. D. Radolf, and A. S. Benzaken, "Syphilis," *Nat Rev Dis Primers*, vol. 3, no. 1, p. 17073, Oct. 2017, doi: 10.1038/nrdp.2017.73.

[2] H. Peyriere, A. Makinson, H. Marchandin, and J. Reynes, "Doxycycline in the management of sexually transmitted infections," *J Antimicrob Chemother*, vol. 73, no. 3, pp. 553–563, Mar. 2018, doi: 10.1093/jac/dkx420.

[3] L. H. Bachmann, "CDC Clinical Guidelines on the Use of Doxycycline Postexposure Prophylaxis for Bacterial Sexually Transmitted Infection Prevention, United States, 2024," *MMWR Recomm Rep*, vol. 73, 2024, doi: 10.15585/mmwr.rr7302a1.

[4] J. Saunders, J. Deering, C. Dewsnap, R. Drayton, and J. Gilmore, "UK National Guideline for the Use of Doxycycline Post-Exposure Prophylaxis (DoxyPEP) for the Prevention of Syphilis".

[5] D. E. DiMarco *et al.*, *Doxycycline Post-Exposure Prophylaxis to Prevent Bacterial Sexually Transmitted Infections*. in New York State Department of Health AIDS Institute Clinical Guidelines. Baltimore (MD): Johns Hopkins University, 2024. Accessed: Feb. 13, 2025. [Online]. Available: http://www.ncbi.nlm.nih.gov/books/NBK597440/

[6] M. Holt *et al.*, "Acceptability of Doxycycline Prophylaxis, Prior Antibiotic Use, and Knowledge of Antimicrobial Resistance Among Australian Gay and Bisexual Men and Nonbinary People," *Sex Transm Dis*, vol. 52, no. 2, pp. 73–80, Feb. 2025, doi: 10.1097/OLQ.0000000000002079.

[7] M. A. Spinelli, H. M. Scott, E. Vittinghoff, A. Y. Liu, K. Coleman, and S. P. Buchbinder, "High Interest in Doxycycline for Sexually Transmitted Infection Postexposure Prophylaxis in a Multicity Survey of Men Who Have Sex With Men Using a Social Networking Application," *Sex Transm Dis*, vol. 46, no. 4, pp. e32–e34, Apr. 2019, doi: 10.1097/OLQ.0000000000000942.

[8] M. W. Traeger, D. S. Krakower, K. H. Mayer, S. M. Jenness, and J. L. Marcus, "Use of Doxycycline and Other Antibiotics as Bacterial Sexually Transmitted Infection Prophylaxis in a US Sample of Primarily Gay and Bisexual Men," *Sexually Transmitted Diseases*, vol. 51, no. 12, p. 763, Dec. 2024, doi: 10.1097/OLQ.0000000000002061.

[9] J. G. Rosenberger *et al.*, "Sexual Behaviors and Situational Characteristics of Most Recent Male-Partnered Sexual Event among Gay and Bisexually Identified Men in the United States," *The Journal of Sexual Medicine*, vol. 8, no. 11, pp. 3040–3050, 2011, doi: 10.1111/j.1743-6109.2011.02438.x.

[10] S. E. D. Quaye *et al.*, "Application of the network scale-up method to estimate the sizes of key populations for HIV in Singapore using online surveys," *J Int AIDS Soc*, vol. 26, no. 3, p. e25973, Mar. 2023, doi: 10.1002/jia2.25973.

[11] M. E. Tudor, A. M. Al Aboud, S. W. Leslie, and W. Gossman, "Syphilis," in *StatPearls*, Treasure Island (FL): StatPearls Publishing, 2025. Accessed: Mar. 10, 2025. [Online]. Available: http://www.ncbi.nlm.nih.gov/books/NBK534780/

[12] M. E. Kent and F. Romanelli, "Reexamining Syphilis: An Update on Epidemiology, Clinical Manifestations, and Management," *Ann Pharmacother*, vol. 42, no. 2, pp. 226–236, Feb. 2008, doi: 10.1345/aph.1K086.

[13] E. G. Clark and N. Danbolt, "The Oslo study of the natural history of untreated syphilis: An epidemiologic investigation based on a restudy of the Boeck-Bruusgaard material a review and appraisal," *Journal of Chronic Diseases*, vol. 2, no. 3, pp. 311–344, Sep. 1955, doi: 10.1016/0021-9681(55)90139-9.

[14] M. J. Tobin, "Fiftieth Anniversary of Uncovering the Tuskegee Syphilis Study: The Story and Timeless Lessons," *Am J Respir Crit Care Med*, vol. 205, no. 10, pp. 1145–1158, doi: 10.1164/rccm.202201-0136SO.

[15] L. K. Whittles, X. Didelot, and P. J. White, "Public health impact and cost-effectiveness of gonorrhoea vaccination: an integrated transmission-dynamic health-economic modelling analysis," *The Lancet Infectious Diseases*, vol. 22, no. 7, pp. 1030–1041, Jul. 2022, doi: 10.1016/S1473-3099(21)00744-1.

[16] J.-M. Molina *et al.*, "Doxycycline prophylaxis and meningococcal group B vaccine to prevent bacterial sexually transmitted infections in France (ANRS 174 DOXYVAC): a multicentre, open-label, randomised trial with a 2 × 2 factorial design," *The Lancet Infectious Diseases*, vol. 24, no. 10, pp. 1093–1104, Oct. 2024, doi: 10.1016/S1473-3099(24)00236-6.

[17] J. Zhang *et al.*, "Discontinuation, suboptimal adherence, and reinitiation of oral HIV pre-exposure prophylaxis: a global systematic review and meta-analysis," *The Lancet HIV*, vol. 9, no. 4, pp. e254–e268, Apr. 2022, doi: 10.1016/S2352-3018(22)00030-3.
[18] "STI Screening Recommendations," Centers for Disease Control and Prevention. Accessed: Jun. 08, 2025. [Online]. Available: https://www.cdc.gov/std/treatment-guidelines/screening-recommendations.htm
[19] L. Geng *et al.*, "Potential public health impacts of gonorrhea vaccination programmes under declining incidences: A modeling study," *PLoS Med*, vol. 22, no. 2, p. e1004521, Feb. 2025, doi: 10.1371/journal.pmed.1004521.
